# Wastewater concentrations of rotavirus RNA are associated with infection and vaccination metrics in the USA

**DOI:** 10.1101/2025.07.21.25331943

**Authors:** Elana M. G. Chan, Alessandro Zulli, Alexandria B. Boehm

## Abstract

Surveillance of rotavirus infections remains critical because vaccines are underutilized in the USA. Using wastewater solids measurements of rotavirus RNA collected over one year from 185 wastewater treatment plants (WWTPs) in the USA, we inferred spatiotemporal occurrence patterns of rotavirus infections and compared occurrence patterns to clinical metrics of infections and markers of vaccination coverage. We also estimated infection prevalence from wastewater measurements using available data on rotavirus RNA shedding in feces. Nationally, wastewater measurements of rotavirus RNA were correlated with clinical metrics of infection and exhibited elevated winter-spring concentrations beginning in the South. WWTP service areas characterized by markers of high vaccination coverage generally experienced a shorter duration of elevated rotavirus concentrations compared to areas characterized by markers of low vaccination coverage. Rotavirus infection prevalence estimates were highly uncertain and sensitive to shedding parameters. Wastewater monitoring of vaccine-preventable diseases is valuable for informing where vaccination campaigns should be targeted.

## Introduction

Rotavirus infection remains the leading cause of diarrheal deaths worldwide, particularly affecting individuals under 5 and over 70 years, despite available vaccines.^1^ Globally, rotavirus deaths declined between 1990 and 2019 but there was a significant uptrend in Canada and the United States—despite North America having some of the highest levels of vaccination coverage in the world.^1,2^ Still, vaccines have alleviated the burden of rotavirus disease. About 2.7 million cases were reported each year in the United States, but vaccines have since annually averted 280,000 clinic visits; 62,000 emergency department visits; and 45,000 hospitalizations.^3^ Continued surveillance of infections is necessary to understand the epidemiology of rotavirus in the post-vaccination era and to identify populations inadequately covered by vaccines. However, rotavirus disease is not nationally notifiable in the United States,^4^ so comprehensive clinical surveillance data remains limited.

Two live-attenuated, oral rotavirus vaccines are currently licensed in the United States. RotaTeq (Merck and Company, Whitehouse Station, New Jersey) was licensed in 2006 and is a three-dose series.^5^ Rotarix (GlaxoSmithKline Biologicals, Rixensart, Belgium) was licensed in 2008 and is a two-dose series.^5^ Both vaccines are administered before 8 months of age and highly effective at preventing severe infection during the first two years of life.^5,6^ Whether vaccines confer lifelong immunity is unknown, but waning effectiveness has been reported in some children.^6,7^ Older children and adults born prior to licensure of vaccines may experience repeat infections throughout their lives, although with reduced disease severity, as immunity from natural rotavirus infection is incomplete.^8–10^ Nonetheless, studies have shown that rotavirus vaccines provide some indirect protection to unvaccinated populations.^11,12^

Despite the important direct and indirect benefits of rotavirus vaccines, vaccines are underutilized in the United States. Coverage for recommended childhood vaccines decreased in the United States during the COVID-19 pandemic.^13^ Estimated rotavirus vaccination coverage was 75.1% in the 2020–2021 birth cohort, which falls below the Healthy People 2020 target of 80% and lags behind other vaccines for the same birth cohort (e.g., poliovirus: 91.1%; measles, mumps, and rubella: 90.3%; varicella: 89.9%).^13,14^ Several geographic, sociodemographic, and healthcare characteristics are documented to influence rotavirus vaccination coverage rates.^14–16^ Still, vaccines have impacted rotavirus seasonality in the United States according to available clinical testing data. Prior to vaccines, rotavirus followed an annual, winter–spring occurrence pattern.^4,17^ Following vaccine introduction, seasonality shifted to a biennial occurrence pattern, with low incidence during even years and elevated incidence during odd years.^4,17^

Symptomatically and asymptomatically infected individuals shed rotavirus RNA in feces and other excreta;^18^ studies have demonstrated that community-level monitoring of rotavirus infections can be conducted via wastewater.^19–24^ Monitoring rotavirus in wastewater is complementary to clinical surveillance because clinical testing has declined since vaccine introduction, is biased towards those with severe disease and access to healthcare, and is reported on a voluntary basis by healthcare providers.^4^ In contrast, a wastewater sample represents an unbiased composite biological sample of an entire contributing population and measurements can be available within 24 hours. Wastewater measurements are inclusive of individuals with self-limiting symptoms who are less likely to seek clinical care, such as adults, and may better represent population-level disease occurrence.^8,9^

In this study, we aimed to characterize spatiotemporal occurrence patterns in rotavirus disease across the United States using wastewater measurements of rotavirus RNA. Samples were collected from 185 wastewater treatment plants (WWTPs) multiple times a week over approximately one year. We compared occurrence patterns of rotavirus disease inferred from wastewater to clinical metrics of rotavirus infections, estimates of rotavirus vaccination coverage, and population characteristics associated with rotavirus vaccination coverage. As a secondary aim, we estimated rotavirus infection prevalence using wastewater rotavirus RNA concentrations and available data on shedding of rotavirus RNA in human feces. Although previous studies have linked rotavirus RNA concentrations in wastewater to clinical disease occurrence (as measured by clinical test positivity rates and reported case numbers),^19–24^ those studies were conducted over small geographic scales and none of them compared rotavirus RNA concentrations to markers of vaccination coverage to assess the utility of wastewater monitoring for identifying vaccination coverage disparities.

## Results

Wastewater samples were typically collected three times per week from 185 WWTPs across 40 states and the District of Columbia between 3 September 2023 and 12 December 2024. Across a total of 33,471 wastewater samples, measured rotavirus RNA concentrations ranged from non-detect to 8.4 × 10^7^ gene copies per gram (gc/g), with a median concentration of 5.1 × 10^4^ gc/g and interquartile range (IQR) of 8.5 × 10^3^–2.4 × 10^5^ gc/g. We detected pepper mild mottle virus (PMMoV) RNA in all wastewater samples, suggesting adequate viral recovery throughout the analysis period (**Figure S1**).

### Occurrence of rotavirus wastewater events across the United States

We examined patterns in rotavirus RNA concentrations in wastewater at the WWTP, state, and national scales to identify periods when concentrations were elevated, suggesting higher than background levels of rotavirus infections; these periods are referred to as rotavirus “wastewater events”. Some locations had more than one wastewater event, so the longest wastewater event is the one described below.

Nationally aggregated, PMMoV-normalized wastewater measurements of rotavirus RNA were variable over the study period, with relatively high values observed during the single national wastewater event occurring between 1 January 2024 and 23 June 2024 (duration: 173 days) (**Figure 1a**). An example rotavirus wastewater event at the WWTP scale is shown in **Figure 1b** for San Jose, California (duration: 316 days). PMMoV-normalized rotavirus RNA wastewater measurements and events for all other WWTPs and states are included in the Supporting Information (SI) (**Figures S2–S3**). At the WWTP scale, wastewater event durations ranged from 20–444 days (median: 182 days); two WWTPs did not experience any wastewater event. At the state scale, the maximum wastewater event duration occurred in Utah (306 days) and the minimum duration occurred in Vermont (14 days); the median duration was 150 days (**Figure S4**).

**Figure 1.**
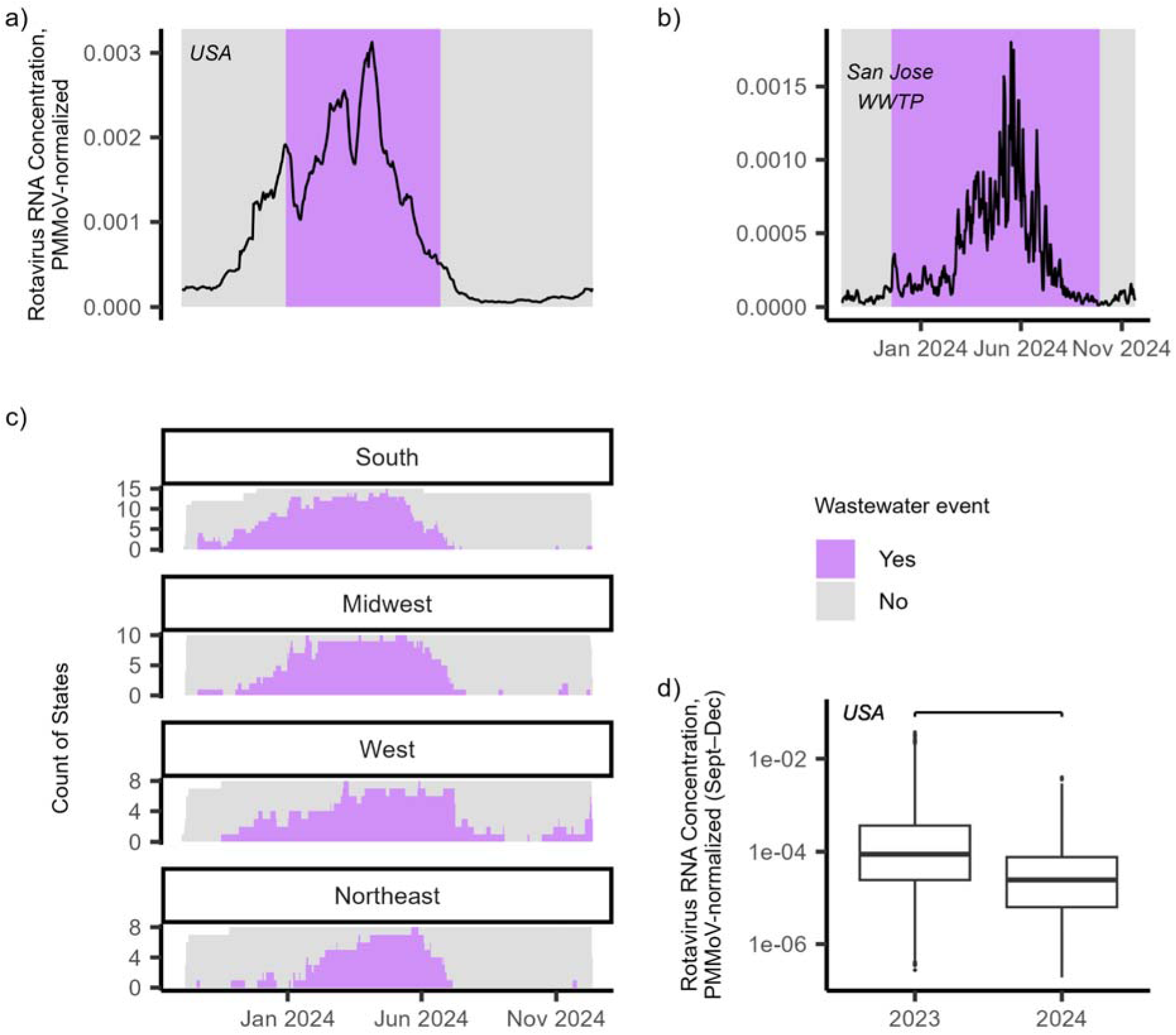
Wastewater rotavirus RNA concentrations and events. (**a**) Nationally aggregated, smoothed, PMMoV-normalized wastewater rotavirus RNA concentrations with the national rotavirus wastewater event shaded. (**b**) Smoothed, PMMoV-normalized wastewater rotavirus RNA concentrations with the rotavirus wastewater event shaded for an individual WWTP (San Jose, California). (**c**) Count of states in a wastewater event each day, grouped by census region. (**d**) Smoothed, PMMoV-normalized wastewater rotavirus RNA concentrations across all samples collected between September–December in 2023 versus 2024. The center line represents the median, the box limits represent the 25th and 75th percentiles, the whiskers represent 1.5 x IQR from the box limits, and points represent values beyond the whiskers. Concentrations are displayed on a log_10_ scale, and the horizontal bar denotes a significant difference. Abbreviations: IQR = interquartile range, PMMoV = pepper mild mottle virus, WWTP = wastewater treatment plant.

Figure 1c displays the number of states experiencing a wastewater event each day, stratified by census region. Wastewater events appeared to onset first in the South followed by the Midwest, West, and Northeast. At least half of states in the region were in onset for 21 consecutive days beginning 10 December 2023 in the South, 1 January 2024 in the Midwest, 25 January 2024 in the West, and 7 February 2024 in the Northeast. Wastewater events appeared to offset first in the South followed by the Northeast, Midwest, and West. Less than half of states in the region were in onset for 21 consecutive days following onset beginning 24 May 2024 in the South, 27 June 2024 in the Northeast, 1 July 2024 in the Midwest, and 10 July 2024 in the West. Across all wastewater samples collected between September–December, PMMoV-normalized wastewater measurements of rotavirus RNA were higher in 2023 compared to 2024 (Wilcoxon test W = 39,152,179; p < 0.0001; n = 8,929 in 2023; n = 6,278 in 2024) (Figure 2d).

**Figure 2.**
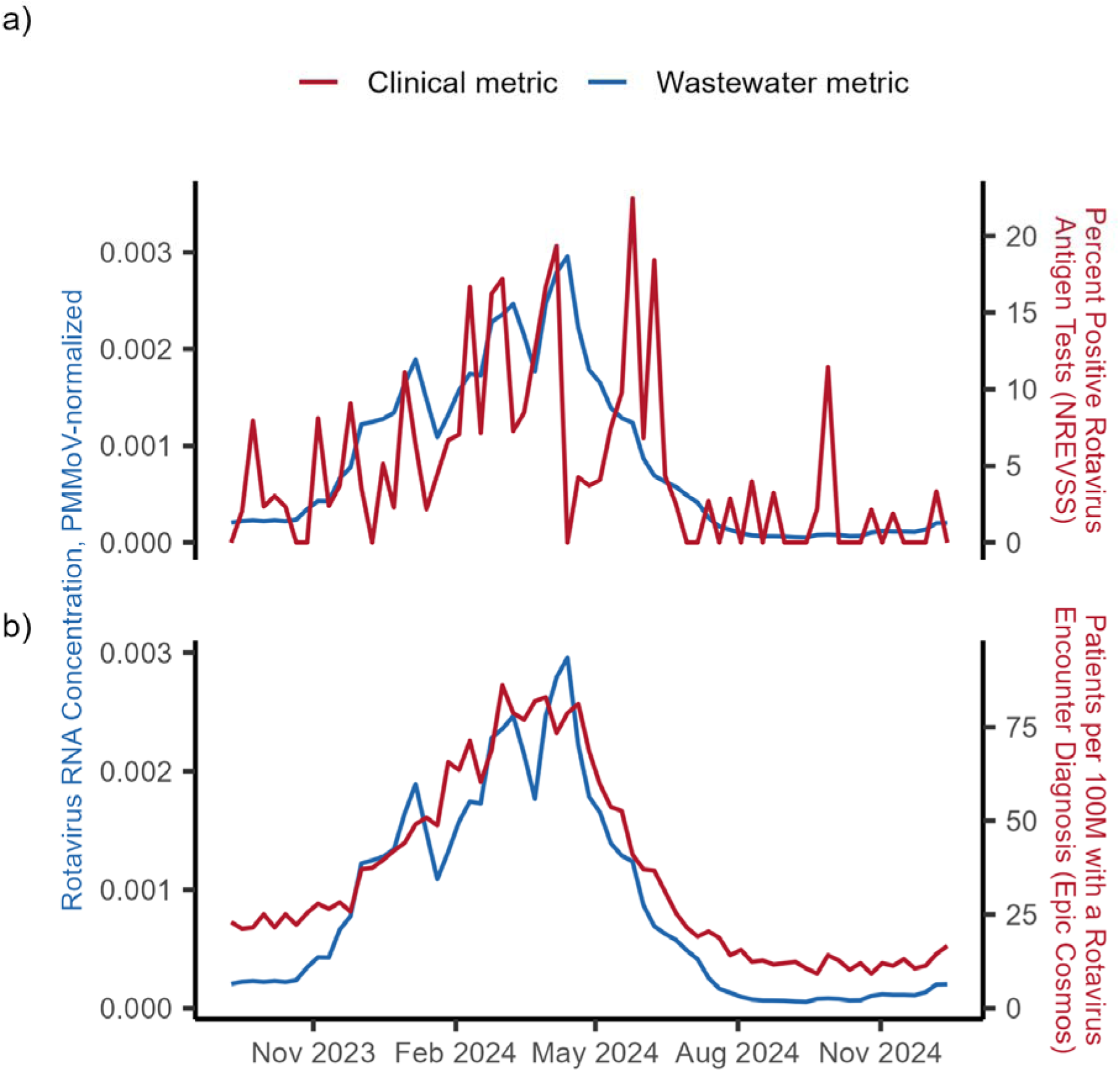
Rotavirus RNA wastewater concentrations and occurrence of rotavirus infections across the United States. Weekly smoothed, PMMoV-normalized rotavirus RNA concentration in wastewater (left y-axis; blue) and weekly clinical surveillance metric of rotavirus infections (right y-axis; red) for the United States. (**A**) Percent of rotavirus antigen tests reported as positive each week from the National Respiratory and Enteric Virus Surveillance System (NREVSS) is used as the clinical metric. (**B**) Patients per 100 million with a rotaviral enteritis encounter diagnosis each week from Epic Cosmos is used as the clinical metric.

### Wastewater measurements of rotavirus RNA compared to clinical metrics of rotavirus infection

Weekly PMMoV-normalized rotavirus RNA wastewater measurements were correlated with weekly rotavirus clinical metrics at the national scale (Figure 2). The correlation coefficient was larger between wastewater measurements and the proportion of patients with a rotaviral enteritis encounter diagnosis from Epic Cosmos (Kendall’s tau = 0.84 [95% CI: 0.80, 0.88], p < 0.0001; n = 67) versus rotavirus test positivity rate from the National Respiratory and Enteric Virus Surveillance System (NREVSS) (Kendall’s tau = 0.49 [95% CI: 0.36, 0.63], p < 0.0001; n = 67).

### Rotavirus wastewater event durations compared to estimates of rotavirus vaccination coverage

Rotavirus wastewater event durations significantly differed across tertiles of vaccination coverage at the sewershed scale using vaccination estimates from Epic Cosmos (χ^2^ = 7.04, p = 0.030); the effect size was small (η^2^ = 0.028) with a significant difference between the middle and highest tertile (Figure 3). At the state scale, rotavirus wastewater event durations did not significantly differ across tertiles of vaccination coverage using rotavirus vaccination estimates from both the US Centers for Disease Control (CDC) National Center for Immunization and Respiratory Diseases (NCIRD) (χ^2^ = 0.67, p = 0.72) and Epic Cosmos (χ^2^ = 2.14, p = 0.34) (**Figure S5**). The state-level rotavirus vaccination estimates from NCIRD and Epic Cosmos were not correlated (**Figure S6**).

**Figure 3.**
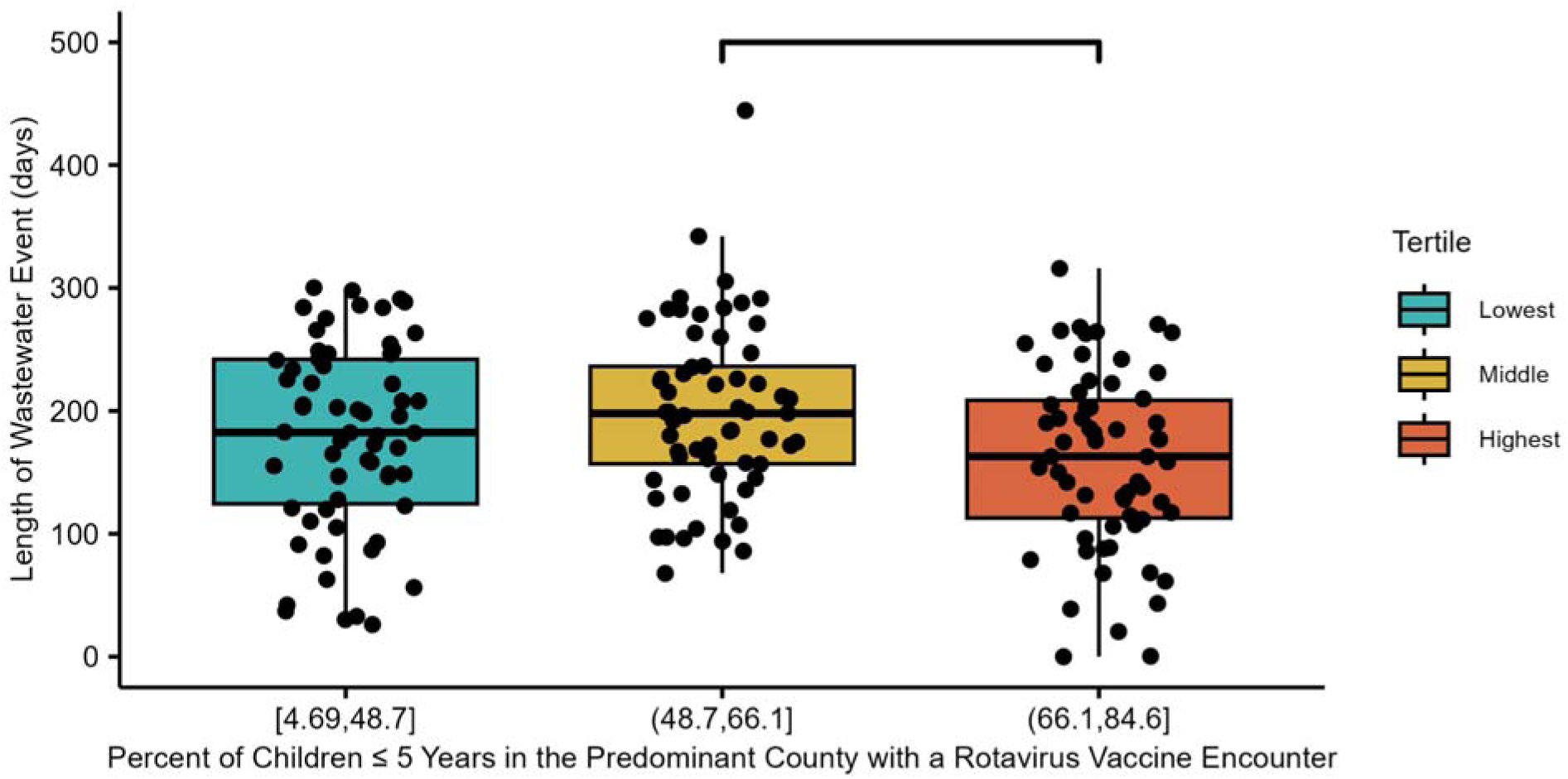
Rotavirus wastewater event durations among sewersheds by tertile of rotavirus vaccination coverage. Distribution of rotavirus wastewater event durations among sewersheds by tertile of rotavirus vaccination coverage in the predominant county of the sewershed. The center line represents the median, the box limits represent the 25th and 75th percentiles, and the whiskers represent 1.5 x IQR from the box limits. Jittered points represent individual data points. Rotavirus vaccination coverage estimates from Epic Cosmos ranged from 4.69–84.6% among sewersheds. Horizontal bar denotes significant pairwise differences. Abbreviations: IQR = interquartile range.

### Rotavirus wastewater event durations compared to population characteristics associated with rotavirus vaccination coverage

Ghaswalla et al.^14^ identified several population characteristics—pertaining to social, geographical, family, and healthcare factors—associated with vaccination coverage in the United States. The distribution of wastewater event durations among sewersheds by population characteristic tertile or census region is shown in Figure 4; refer to the SI for tertile cutoffs (**Figures S7–S10**). Horizontal bars denote significant pairwise differences following Kruskal Wallis tests with p < 0.05. Refer to **Table 1** for chi square test statistics, p values, and eta-squared effect size estimates for each Kruskal-Wallis test. We did not observe a significant difference across tertiles for Hispanic or Latino children or pediatrician-to-child ratio. We observed a significant difference across tertiles for publicly insured children; however, the effect size was small (η^2^ = 0.024) and no significant pairwise differences were identified. We observed a significant difference across groups for all other characteristics with a small effect size for foreign-born children (η^2^ = 0.044), children below poverty (η^2^ = 0.044), average family size (η^2^ = 0.034), and urbanicity (η^2^ = 0.053); a moderate effect size for Black or African American children (η^2^ = 0.073), uninsured children (η^2^ = 0.074), and children born to younger mothers (η^2^ = 0.098); and a large effect size for census region (η^2^ =0.14).

**Figure 4.**
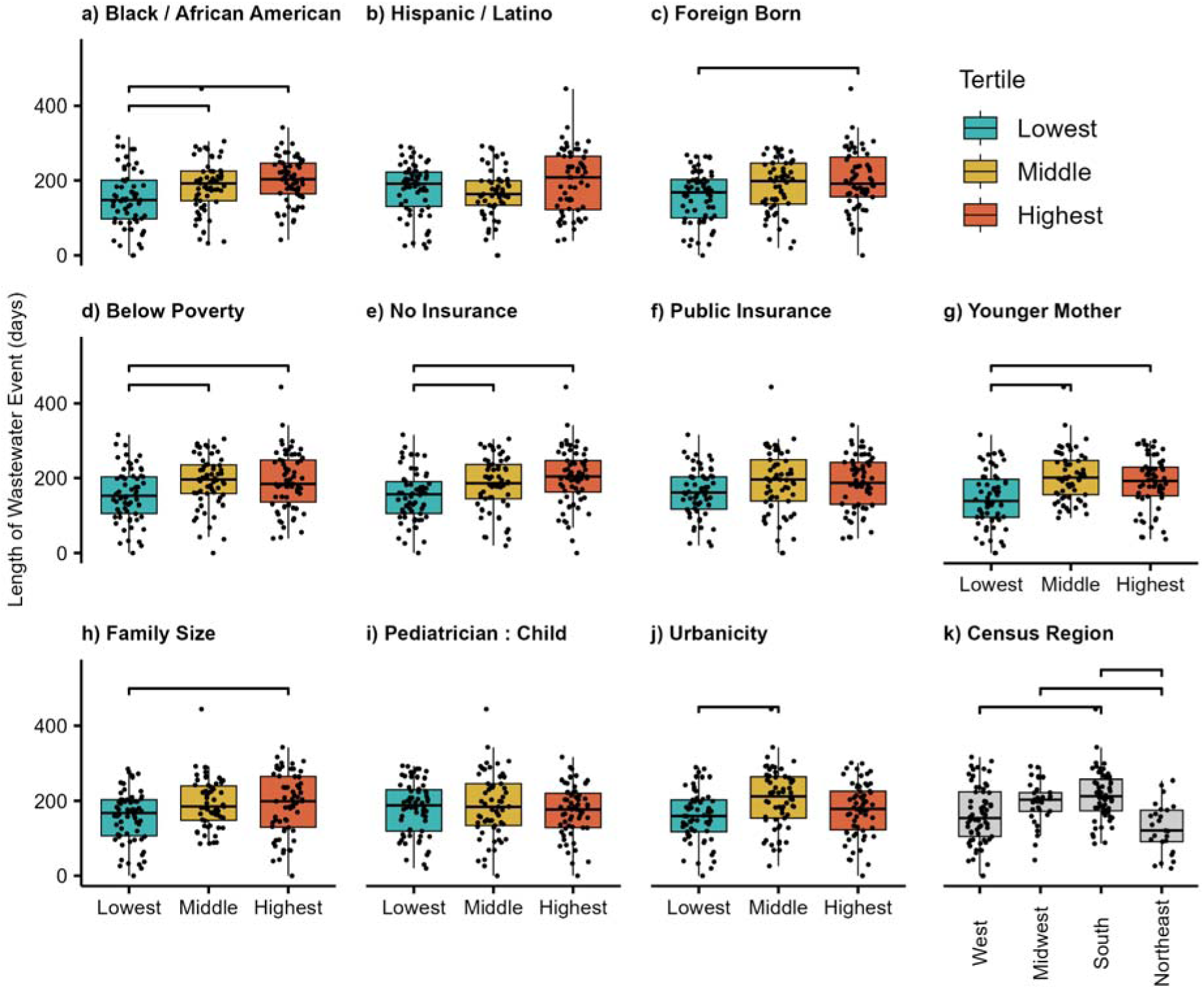
Rotavirus wastewater event durations by population characteristics. Distribution of rotavirus wastewater event durations among sewersheds by population characteristic tertiles (**a–j**) or census region (**k**). The center line represents the median, the box limits represent the 25th and 75th percentiles, and the whiskers represent 1.5 x IQR from the box limits. Jittered points represent individual data points. Horizontal bars denote significant pairwise differences. Refer to the Supporting Information for tertile cutoffs of population characteristics in a–j. Abbreviations: IQR = interquartile range.

**Table 1.** Association between rotavirus wastewater event duration and population characteristics at the sewershed scale.

| **Sewershed Characteristic** | **Kruskal-Wallis $\chi^2$ (p value)<sup>a</sup>** | **Eta-squared (effect size)** | **Conover-Inman Post-Hoc Test: Significant Pairwise Differences<sup>a</sup>** |
| --- | --- | --- | --- |
| Percent of children under 5 years, Black or African American alone | 15.21 (0.00050) | 0.073 (moderate) | lowest-middle<br>lowest-highest |
| Percent of children under 5 years, Hispanic or Latino | 4.65 (0.098) | not conducted | not conducted |
| Percent of children under 5 years, foreign born | 9.95 (0.0069) | 0.044 (small) | lowest-highest |
| Percent of children under 5 years, income in the past 12 months below the federal poverty level | 10.06 (0.0065) | 0.044 (small) | lowest-middle<br>lowest-highest |
| Percent of children under 6 years, no health insurance | 15.44 (0.00044) | 0.074 (moderate) | lowest-middle<br>lowest-highest |
| Percent of children under 6 years, public health insurance | 6.29 (0.043) | 0.024 (small) | none |
| Percent of women who gave birth in the past 12 months, ages 15-24 years | 19.91 (< 0.0001) | 0.098 (moderate) | lowest-middle<br>lowest-highest |
| Average family size | 8.11 (0.017) | 0.034 (small) | lowest-highest |
| Pediatricians per 100,000 children < 18 years | 0.77 (0.68) | not conducted | not conducted |
| Urbanicity | 11.69 (0.0029) | 0.053 (small) | lowest-middle |
| Census region | 28.97 (< 0.0001) | 0.14 (large) | Northeast-Midwest<br>Northeast-South<br>South-West |

<sup>a</sup> Sewersheds were grouped based on whether they fell into the lowest, middle, or highest tertile for each characteristic in order to conduct Kruskal-Wallis and post hoc tests.

### Estimated population shedding rotavirus RNA from measurements of rotavirus RNA in wastewater solids

We adopted a virus-independent mass balance model established by Wolfe et al.^25^ to estimate the population fraction shedding rotavirus RNA into wastewater each day (F_shed_). The model uses PMMoV-normalized measurements of rotavirus RNA in wastewater solids (C_rota_ww_/C_PMMoV_ww_) and fecal shedding concentrations for PMMoV RNA (C_PMMoV_feces_) and rotavirus RNA (C_rota_feces_) as inputs. Arts et al.^26^ and Zheng et al.^27^ provide large datasets on fecal shedding which were used to characterize the distributions of C_PMMoV_feces_ and C_rota_feces_, respectively. The distributions of C_PMMoV_feces_ and C_rota_feces_ were reasonably modeled as log_10_-normal distributions upon examination of quantile-quantile plots. The mean and standard deviation of log_10_-transformed concentrations of PMMoV RNA measured in feces in gc/g from Arts et al.^26^ is 7.94 and 1.57 log_10_ gc/g, respectively. The mean and standard deviation of log_10_-transformed concentrations of rotavirus RNA measured in feces in gc/g from Zheng et al.^27^ is 7.01 and 2.16 log_10_ gc/g, respectively.

Figure S11 shows F_shed_ expressed as a percentage modeled from C_rota_ww_/C_PMMoV_ww_. Daily estimates of the percentage of people shedding rotavirus RNA for an individual WWTP (San Jose, California) and across all WWTPs for the United States are shown in Figure 5. At the San Jose, California WWTP, C_rota_ww_/C_PMMoV_ww_ ranged from 0.000011 to 0.0018, corresponding to a percentage of people shedding rotavirus RNA of 0.0094% (25th percentile = 0.00015%, 75th percentile = 0.61%) and 1.5% (25th percentile = 0.026%, 75th percentile = 110%). Nationally aggregated C_rota_ww_/C_PMMoV_ww_ across all WWTPs in the study ranged from 0.000053 to 0.0031, corresponding to a percentage of people shedding rotavirus RNA of 0.040% (25th percentile = 0.00059%, 75th percentile = 2.5%) and 3.0% (25th percentile = 0.046%, 75th percentile = 184%).

**Figure 5.**
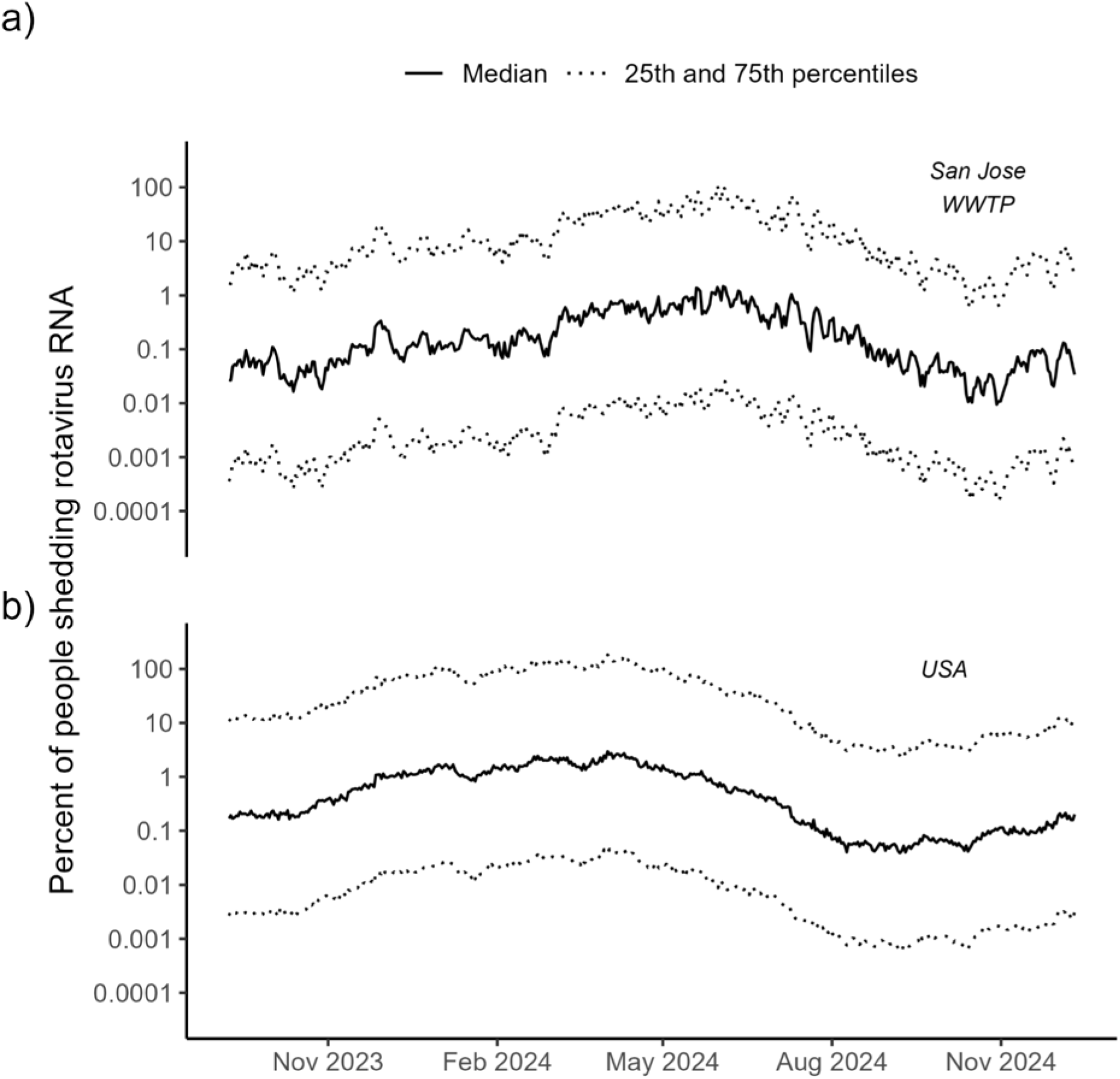
Daily estimates of the percentage of the population shedding rotavirus RNA. (**a**) Estimates for an individual WWTP (San Jose, California). (**b**) National estimates across all WWTPs. Solid line represents the median estimate; dotted lines represent the 25th and 75th percentile estimates. Abbreviation: WWTP = wastewater treatment plant.

The sensitivity analysis suggests the model is highly sensitive to all input parameters. We calculated F_shed_ for each parameter at its 75th percentile (F_shed,75_) and 25th percentile (F_shed,25_) holding all other parameters at their 50th percentile. The F_shed,75_:F_shed,25_ ratio was 0.0013 for C_rota_feces_, 987 for C_PMMoV_feces_, and 100 for C_rota_ww_/C_PMMoV_ww_. The fraction of the population shedding rotavirus RNA decreases as the concentration of rotavirus in feces increases, increases as the concentration of PMMoV in feces increases, and increases as the PMMoV-normalized rotavirus RNA concentration in wastewater increases.

## Discussion

Rotavirus disease is not nationally notifiable in the United States, yet surveillance of rotavirus infections is critical for identifying gaps in rotavirus vaccination coverage and understanding rotavirus epidemiology in the post-vaccination era. We used wastewater RNA concentrations of rotavirus collected over more than one year across the United States to infer rotavirus disease occurrence in the United States and compare occurrence patterns to clinical metrics of rotavirus infection and markers of rotavirus vaccination coverage. Rotavirus RNA was detected in wastewater samples throughout the study period in varying concentrations.

Prior to our study, concentrations of rotavirus RNA in wastewater solids have only been reported in California.^19^ Rotavirus has been detected in wastewater sludge in several countries,^28^ but our study is the first to report concentrations in wastewater solids across the United States. Boehm et al.^19^ retrospectively measured rotavirus RNA concentrations in wastewater solids at two WWTPs in California between February 2021–April 2023 and observed median concentrations on the order of 10^3^ gc/g. Herein, we observed median concentrations of rotavirus RNA in wastewater solids on the order of 10^4^ gc/g based on samples from 185 WWTPs across 40 states and the District of Columbia between September 2023– December 2024. Studies that measured rotavirus RNA concentrations in raw sewage observed seasonal occurrence patterns, as we observed in our study, with concentrations ranging on the order of 10^2^–10^8^ gc/L and 1–10^4^ fluorescent foci/L.^29–34^

Nationally, a rotavirus wastewater event occurred 1 January to 23 June which aligned with the winter–spring occurrence pattern of rotavirus infections according to clinical testing data.^4,17^ Wastewater measurements of rotavirus RNA were higher in fall 2023 (leading up to an even-numbered season) than in fall 2024 (leading up to an odd-numbered season) across the United States. We did not have longitudinal wastewater measurements from two consecutive winter– spring seasons, but our observations suggest a deviation from the typical biennial occurrence pattern of rotavirus. According to clinical testing data, odd-numbered seasons are generally associated with elevated incidence compared to even-numbered seasons in the post-vaccination era.^4,17^ The COVID-19 pandemic impacted the biennial pattern of rotavirus occurrence.^35^ Mathematical modeling suggests rotavirus transmission will not return to pre-pandemic equilibrium before 2030 which may explain why wastewater rotavirus RNA measurements seemingly did not align with the biennial occurrence pattern.^35^ Continued wastewater monitoring of rotavirus RNA can be a useful tool for empirically understanding how temporal patterns in rotavirus occurrence continue to develop.

The spatial progression of rotavirus occurrence in wastewater lacked national coherency. Elevated occurrence of rotavirus in wastewater began first in the South followed by the Midwest, West, and Northeast. Prior to vaccines, annual rotavirus occurrence also lacked national coherency, beginning in the Southwest and ending in the Northeast.^36,37^ In the post-vaccination era, Burnett et al.^38^ suggest areas with high birth rate and low vaccination coverage have the most rapid accumulation of susceptible individuals and consequently drive annual rotavirus occurrence. Indeed, rotavirus occurrence began in Oklahoma, Arkansas, and the western Gulf coast—all areas in the South with high birth rate and low vaccination coverage—according to clinical testing data across five seasons in the post-vaccination era.^38^ Wastewater monitoring of rotavirus RNA over consecutive seasons can be used alongside clinical testing data to characterize patterns in the spatial progression of rotavirus in the post-vaccination era.

Weekly wastewater measurements of rotavirus RNA were positively correlated with clinical metrics of rotavirus infections obtained from both NREVSS and Epic Cosmos at the national scale. Although the two clinical metrics themselves were positively correlated at the national scale (**Figure S12**), the two datasets are distinct. Epic Cosmos is a research dataset that aggregates electronic health record data from 295 million patients.^39,40^ In contrast, NREVSS relies on voluntary reporting of rotavirus tests; fewer than 100 rotavirus test results were submitted to NREVSS each week during our analysis period. As shown in Figure 3, test positivity from NREVSS was visually prone to more week-to-week variability compared to patient diagnoses from Epic Cosmos. Moreover, data can be accessed at subnational scales from Epic Cosmos unlike from NREVSS.^40^ For example, we observed wastewater measurements of rotavirus RNA were still positively correlated with clinical metrics of rotavirus infections at the state scale using data from Epic Cosmos (**Figure S13**, **Table S1**). Nonetheless, electronic health record datasets like Epic Cosmos still suffer biases not present in wastewater monitoring data, such as bias towards individuals with symptomatic disease and access to healthcare. Rotavirus encounter diagnoses in Epic Cosmos mostly occur in the under 5 year age group (**Figure S14**), suggesting infections in older populations are subclinical. Likewise, other studies observed a weak or lack of a positive correlation between rotavirus RNA concentrations in wastewater and traditional measures of disease surveillance, suggesting the presence of asymptomatic or subclinical infections in the contributing community.^19–24^ Wastewater monitoring represents a less-biased approach for inferring population-level disease occurrence compared to clinical or syndromic measures of disease surveillance.

Rotavirus wastewater event durations were generally shorter in sewersheds with markers of high rotavirus vaccination coverage, suggesting the utility of wastewater monitoring for identifying gaps in vaccination coverage. Exceptions include wastewater event durations not being associated with the proportion of Hispanic or Latino children or the pediatrician-to-child ratio and not being highest in the Northeast.^14^ However, not all indirect markers of vaccination coverage identified by Ghaswalla et al.^14^ were directly available from publicly available sources. For example, we obtained public data on pediatrician-to-child ratio which describes access to pediatricians but not whether children actually seek care from pediatricians over family physicians. We also only conducted bivariate analyses between wastewater event durations and each indirect marker of rotavirus vaccination coverage. Future work might consider combining all markers of rotavirus vaccination coverage into a multivariable model with wastewater event duration as the outcome variable. Additionally, other metrics (e.g., peak wastewater measurement) may be important to consider as the outcome variable. Herein, we were interested in the effect of vaccination on prolonged rotavirus occurrence rather than an isolated outbreak given that vaccines have been documented to shorten rotavirus season.^41^ A limitation with our selected dependent variable, however, is that the wastewater event duration may be underestimated for some WWTPs and consequently some states. The wastewater sampling start and end dates differed across WWTPs (**Table S2**), so sampling may have ended in the midst of a wastewater event for some WWTPs. Nonetheless, the national wastewater event ended before 1 July 2024, which is when a large number of WWTPs ended sampling (mainly in California); it is likely durations were underestimated for only a few WWTPs.

Prior to vaccines, approximately 2.7 million cases of rotavirus were reported each year in the United States,^3^ corresponding to about 1% of the population based on the population estimate from the 2000 census.^42^ Annual rotavirus incidence estimates in the United States are not available for the post-vaccination era but are expected to be substantially lower given the effectiveness of vaccines at preventing severe infection.^5,6^ Infected individuals may shed rotavirus RNA for consecutive days;^27^ therefore, modeled F_shed_ values are likely more representative of disease prevalence rather than incidence. Thus, it is challenging to evaluate the accuracy of our model because rotavirus is monitored in terms of incidence (i.e., reported cases) rather than prevalence. Nonetheless, we expect our model estimates higher rotavirus prevalence than could be determined from case reporting. The age-adjusted prevalence of asymptomatic rotavirus infection is 11% and rotavirus may be detected in about 20–30% of individuals without diarrhea.^43,44^ Rotavirus infection in adults also usually results in self-limiting symptoms,^8^ so adult infections are likely not captured in case reporting.

As indicated by our sensitivity analysis, the model is highly sensitive to all parameters. Fecal shedding dynamics are highly individualized,^27^ and our model demonstrates that precisely estimating disease occurrence from wastewater measurements is extremely challenging. Infected individuals who shed rotavirus RNA in feces in high concentrations or for long durations (“super shedders”) likely contribute the most rotavirus RNA to wastewater. Future work is needed to better characterize fecal shedding dynamics and refine models so disease occurrence may be estimated from wastewater measurements with less uncertainty. Additionally, rotavirus RNA in wastewater solids was measured using droplet digital reverse transcription-polymerase chain reaction (ddRT-PCR) without first subjecting samples to a “heat snap” (i.e., incubating samples at 99°C for 5 minutes).^45^ Rotavirus is a double-stranded RNA (dsRNA),^2^ and the additional denaturing step may be important for quantifying RNA of dsRNA viruses using ddRT-PCR.^46,47^ Including a heat snap step may increase measured rotavirus RNA concentrations in wastewater which could affect modeled F_shed_ reported herein.

Our study is the first to assess spatiotemporal occurrence patterns of rotavirus RNA in wastewater across the United States and compare occurrence patterns to markers of rotavirus vaccination coverage. Wastewater monitoring has been used previously to support polio eradication efforts.^48^ Wastewater monitoring of other vaccine-preventable diseases may similarly be valuable for identifying disparities in vaccination coverage and informing where vaccination campaigns should be targeted.^49,50^ Although this study focused on the United States, the global burden of rotavirus is disproportionately highest in Africa, Oceania, and South Asia and rotavirus vaccines have lower performance in countries with high child mortality.^1,6^ Therefore, wastewater monitoring for rotavirus could be especially impactful in low- and middle-income countries. Continued work on adapting wastewater monitoring methods to decentralized or non-sewered settings is needed to scale wastewater monitoring of rotavirus globally.^51–55^

## Methods

### Wastewater monitoring

Samples were collected from 185 WWTPs across 40 states and the District of Columbia (Figure 6). The number of WWTPs in states ranged from 1–54 (median: 2 WWTPs). The reported population serviced by the WWTPs ranged from 3,000–4,000,000 (median: 89,847). Wastewater samples were typically collected three times per week from WWTPs between 3 September 2023 and 12 December 2024, resulting in 33,471 total samples (**Table S2**). Following environmental molecular biology best practices,^56^ concentrations of rotavirus and pepper mild mottle virus (PMMoV) RNA in the solid fraction of wastewater samples were measured in gene copies per gram (gc/g) using ddRT-PCR. Non-detect rotavirus RNA concentrations were set to 500 gc/g (approximately half the assay detection limit). PMMoV is an indigenous wastewater virus of dietary origin.^57,58^ Full methodological details and data collected between 1 January 2022 to 30 June 2024 are available in a data descriptor by Boehm et al.^45^ Data between 1 July 2024 and 12 December 2024, which were measured using the exact methods reported by Boehm et al.^45^, are uniquely presented in this paper and are available through the Stanford Digital Repository (https://doi.org/10.25740/mv671wb2446).^59^ Based on our *in silico* evaluation described in the SI, it is improbable the rotavirus assay used herein amplifies either the RotaTeq or Rotarix human vaccine strain, animal rotavirus strains, or the bovine rota-coronavirus vaccine spiked into samples as an exogenous control due to the presence of base pair mismatches with the assay primers and/or probe (**Text S1**).

**Figure 6.**
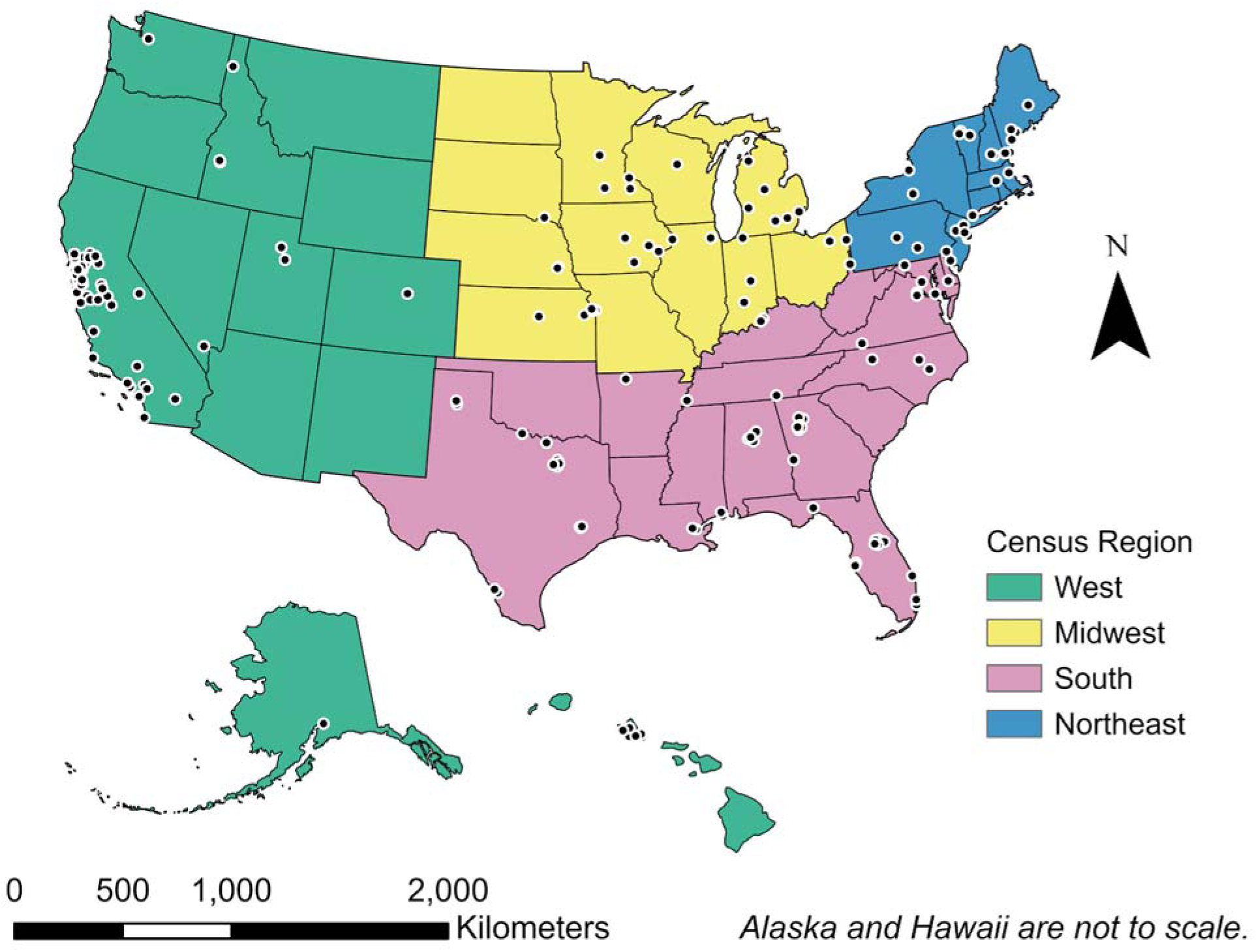
Map of national wastewater monitoring locations. Locations of WWTPs are symbolized by circles, and states are colored by census region. Created in ArcGIS Pro (version 3.1.1) using 2023 state cartographic boundaries (20-meter resolution) from the US Census Bureau (accessed 5 Aug 2024).^60^ Abbreviation: WWTP = wastewater treatment plant.

### Clinical surveillance

We obtained clinical surveillance metrics of rotavirus infections from the National Respiratory and Enteric Virus Surveillance System (NREVSS) and Epic Cosmos (Epic Systems Corporation, Verona, Wisconsin, United States). NREVSS is a laboratory-based sentinel surveillance system that monitors respiratory and enteric virus disease in the United States. Participating laboratories voluntarily report to the Centers for Disease Control (CDC) the number of fecal specimens tested and the number of those specimens that tested positive for rotavirus on a weekly basis.^4^ We obtained the publicly available, centered, three-week moving average rotavirus test positivity rate each week (Sunday–Saturday) for the United States for weeks ending 9 September 2023 through 14 December 2024 from the public-facing NREVSS dashboard.^61^ The number of specimens tested for rotavirus across the entire NREVSS network during these weeks ranged from 16–80 tests (median: 41 tests).

Epic Cosmos is an electronic health record dataset obtained from participating organizations that use Epic Systems.^39,40^ Cosmos combines data from 295 million patients across over 37,000 clinics and 1,500 hospitals; the dataset is representative of patients who seek healthcare in the United States.^39^ We used Cosmos to identify diagnoses of rotaviral enteritis over the study period. First, we geographically filtered the dataset for patients in the United States and temporally filtered the dataset to include patients alive between 1 September 2023 and 14 December 2024. From this subset, we determined the number of patients with a rotaviral enteritis encounter diagnosis each week (Sunday–Saturday). We used these data to calculate the weekly proportion of all patients alive between 1 September 2023 and 14 December 2024 with a rotavirus encounter diagnosis in the United States. Diagnoses were identified using the International Classification of Diseases (ICD-10) code A08.0 for rotaviral enteritis.^62^

### Vaccination coverage

We obtained rotavirus vaccination coverage estimates from the CDC’s National Center for Immunization and Respiratory Diseases (NCIRD) and Epic Cosmos. NCIRD conducts the National Immunization Surveys (NIS) which are phone surveys used to estimate vaccination coverage for vaccinations recommended by the Advisory Committee on Immunization Practices (ACIP).^63^ National, regional, state, and selected local area vaccination coverage estimates by birth year and 2- and 4-year birth cohorts are available for rotavirus vaccination from NCIRD.^64^ For this analysis, we used state-level rotavirus vaccination coverage estimates from the latest available 2-year birth cohort (2020–2021).

To obtain rotavirus vaccination estimates from Epic Cosmos, we first filtered the dataset for patients alive and ≤ 5 years of age in the United States between 1 January 2019 and 12 December 2024. We selected children ≤ 5 years because we aimed to estimate patterns in rotavirus vaccination coverage in the past 5 years. From this subset, we determined the number of patients with any rotavirus immunization encounter grouped by (1) state and (2) county. We used these data to calculate the proportion of all patients alive and ≤ 5 years between 1 January 2019 and 12 December 2024 with a rotavirus immunization encounter at the state and county level.

### Population characteristics

A systematic review on rotavirus vaccination coverage in the United States by Ghaswalla et al.^14^ identified social, geographical, family, and healthcare characteristics associated with vaccination coverage.^14^ Using publicly available data, we determined the value of 11 variables pertaining to these characteristics for each WWTP sewershed (i.e., the geographic area serviced by a WWTP) (**Table 2**).^65–68^ Most WWTPs (n = 113) provided a sewershed boundary. For WWTPs that did not provide a sewershed boundary (n = 72), we approximated a boundary based on the reported zip code(s) serviced by the WWTP and 2023 United States ZIP Code Boundaries published by Esri (Source: TomTom, US Postal Service, Esri).^69^ However, sewershed boundaries do not align with legal boundaries (e.g., countries or census tracts) at which publicly available data are often reported. Refer to the SI for details about how we derived sewershed-level estimates for each characteristic using data from each source (**Equations S1–S2**, **Table S3**).

**Table 2.** Characteristics associated with rotavirus vaccination coverage in the United States as reported by Ghaswalla et al.^14^.

| Sewershed Characteristic | Data Source |
| --- | --- |
| Percent of children under 5 years, Black or African American alone | 5-year American Community Survey estimates (2022)<sup>65</sup> |
| Percent of children under 5 years, Hispanic or Latino |  |
| Percent of children under 5 years, foreign born |  |
| Percent of children under 5 years, income in the past 12 months below the federal poverty level |  |
| Percent of children under 6 years, no health insurance |  |
| Percent of children under 6 years, public health insurance |  |
| Percent of women who gave birth in the past 12 months, ages 15-24 years |  |
| Average family size |  |
| Pediatricians per 100,000 children < 18 years | American Board of Pediatrics (2024)<sup>66</sup> |
| Urbanicity | US Census Bureau (2020, 1984)<sup>67,68</sup> |
| Census region |  |

### Data analysis

Unless otherwise stated, we normalized rotavirus RNA concentrations by PMMoV RNA concentrations to correct for differences in virus recovery and wastewater fecal strength and then smoothed the data to reduce effects of outliers.^25,45^ We smoothed measured PMMoV-normalized concentrations at each WWTP by calculating the five-sample, centered, trimmed, moving average.^45^ Trimming removes the highest and lowest value in the moving average window prior to calculating the average. A shrinking window was used to calculate moving average values at the start and end of a time series (e.g., a three-sample window was used to calculate the moving average value for the first sample). Herein, “wastewater measurement” refers to the smoothed, PMMoV-normalized concentration unless otherwise stated.

We spatially aggregated wastewater measurements across WWTP sewersheds using a population-weighted averaging approach which has been previously described.^70–73^ We used WWTP service populations (**Table S2**) to weight wastewater measurements across WWTPs in each state and 2023 state populations to weight wastewater measurements across states for the United States (**Equations S3–S4**).^74^ Sewershed boundaries do not align with jurisdictional boundaries at which public health data are reported, so spatially aggregating wastewater measurements to state and national scales allowed us to compare wastewater measurements to clinical or vaccination data.

For modeling (described later) and data visualization purposes only, we generated daily wastewater measurements for each WWTP prior to spatial aggregation. We used linear interpolation between adjacent wastewater measurements to generate a wastewater measurement each day that was either interpolated or based on measured values; no interpolation was performed if there was a gap in sampling > 10 days.

#### Year comparison

Rotavirus disease is known to follow a biennial occurrence pattern in the post-vaccination era,^4,17^ and wastewater monitoring data were available in both 2023 and 2024 during fall (September–December). Log_10_-transformed wastewater measurements of rotavirus RNA were not normally distributed in 2023 and 2024 upon examination of quantile-quantile plots. Therefore, we used the two-sided Wilcoxon rank sum test to evaluate the null hypothesis that log_10_-transformed wastewater measurements of rotavirus RNA collected between September–December did not differ in 2023 compared to 2024.

#### Wastewater events

We inferred the onset and offset date of rotavirus epidemics for WWTPs using wastewater rotavirus RNA concentrations as measured (i.e., prior to smoothing and PMMoV normalization). Here, a wastewater event refers to the period between onset and offset dates, and the duration of a wastewater event is the number of days between corresponding onset and offset dates. We defined onset as the first date that wastewater rotavirus RNA concentrations in all wastewater samples collected in the past 14 days were ≥ 25,000 gc/g (approximately half the median concentration detected across all samples in this study). We defined offset as the first date after onset that wastewater rotavirus RNA concentrations in ≤ 50% of wastewater samples collected in the past 14 days were ≥ 25,000 gc/g. WWTPs did not have an onset or offset designation for the first 14 days of data availability.

To identify wastewater events at the state scale, we defined onset as the first date that ≥ 50% of WWTPs in the state were in onset and offset as the first date after onset that < 50% of WWTPs in the state were in onset.^70–72^ Likewise, we determined wastewater events at the national scale by defining onset as the first date that ≥ 50% of states in the United States were in onset and offset as the first date after onset that < 50% of states in the United States were in onset.^70–72^ For all spatial scales (WWTP, state, national), we identified all occurrences of wastewater events. Sometimes more than one wastewater event occurred, so the longest wastewater event is referred to as the “wastewater event” below.

#### Correlation with clinical metrics

We evaluated the correlation between wastewater measurements of rotavirus RNA and clinical metrics of rotavirus infections. We assessed the correlation on a weekly basis at the national scale using two clinical rotavirus metrics: (1) rotavirus test positivity rate from NREVSS and (2) proportion of patients with a rotaviral enteritis encounter diagnosis from Epic Cosmos. Clinical metrics were obtained on a weekly basis; we averaged wastewater measurements across all samples for each WWTP on a weekly basis to match the reporting frequency of the clinical metrics prior to spatially aggregating measurements to the national scale. Nationally aggregated weekly wastewater measurements were not normally distributed (Shapiro-Wilk test, p < 0.0001); therefore, we used a two-sided Kendall’s tau correlation to test the null hypothesis that weekly wastewater measurements of rotavirus RNA are not correlated with weekly clinical metrics of rotavirus infections for the United States. We used the KendallTauB function in the DescTools library to determine the 95% confidence interval (CI) of Kendall’s tau estimates.^75^ Because we conducted two hypothesis tests (i.e., one test for each clinical metric), we used an alpha value of 0.05 / 2 = 0.025 to account for multiple hypothesis testing when assessing statistical significance.

#### Correlation with vaccination coverage

We evaluated the correlation between the rotavirus wastewater event duration and rotavirus vaccination coverage at the state and sewershed scales. At the state scale, rotavirus vaccination coverage estimates were available from (1) NCIRD and (2) Epic Cosmos. Rotavirus vaccination coverage is not directly estimated at the sewershed scale, so we used the vaccination coverage estimate of the county that a sewershed predominantly occupies (**Table S2**) to represent the vaccination coverage of sewersheds (**Figure S15**). County-level rotavirus vaccination coverage estimates are only available from Epic Cosmos; therefore, we only used one estimate of rotavirus vaccination coverage to evaluate this correlation at the sewershed scale. For both spatial scales, we grouped states or sewersheds into vaccination coverage tertiles (lowest, middle, and highest levels of vaccination coverage). Wastewater event durations were not normally distributed among states or sewersheds within all vaccination tertiles (Shapiro-Wilk, p < 0.05); therefore, we used Kruskal-Wallis test to assess the null hypothesis that the wastewater event duration among states or sewersheds is equal across vaccination coverage tertiles. Because we conducted two hypothesis tests at the state scale (i.e., one test for each vaccination estimate), we used an adjusted p value of 0.05 / 2 = 0.025 to account for multiple hypothesis testing when assessing statistical significance. We used a p value of 0.05 to assess statistical significance at the sewershed scale because we only conducted one hypothesis test at this scale. When the null hypothesis was rejected, we used the kruskall_effsize function in the rstatix library to determine the eta-squared effect size measure and the Conover-Inman post-hoc test with a Bonferroni correction to determine significant pairwise differences.^76,77^

#### Correlation with population characteristics

We evaluated the correlation between rotavirus wastewater event duration and each population characteristic associated with rotavirus vaccination coverage at the sewershed scale. For each population characteristic except census region, we grouped sewersheds into tertiles (lowest, middle, and highest levels of the population characteristic); census region was already a categorical variable. Wastewater event durations were not normally distributed among sewersheds within all groupings for each population characteristic (Shapiro-Wilk, p < 0.05); therefore, we used Kruskal-Wallis test to assess the null hypothesis that the wastewater event duration among sewersheds is equal across groups for each population characteristic. We used a p value of 0.05 to assess statistical significance because we did not conduct exploratory multiple hypothesis testing.^14^ When the null hypothesis was rejected, we used the kruskall_effsize function in the rstatix library to determine the eta-squared effect size measure and the Conover-Inman post-hoc test with a Bonferroni correction to determine significant pairwise differences.^76,77^

#### Estimation of shedding population

We estimated the fraction of the population shedding rotavirus RNA into wastewater from rotavirus RNA measurements in wastewater solids. We used a mass balance model established by Wolfe et al.^25^ for SAR-CoV-2 RNA. The model is not virus specific and was adopted by Boehm et al.^78^ for human norovirus GII RNA. Herein, we adopted the model for rotavirus RNA. Briefly, the model assumes fecal shedding is the predominant source of viral RNA into the waste stream and that the fraction of wastewater solids of fecal origin can be approximated from the concentration of PMMoV RNA in wastewater solids (**Equation S5**). Similar to Boehm et al.^78^, we neglect viral RNA decay given the time sewage spends in the system, including the primary clarifier, is short (< 1 day) and that the first order decay rate constant for rotavirus RNA and PMMoV RNA can reasonably be assumed to be both small and similar in magnitude (**Equation S6**).^25,79–82^ Lastly, we assume the solid-liquid partitioning coefficient for rotavirus RNA and PMMoV RNA are equivalent like Boehm et al.^78^, leading to the simplified model shown in **Equation 1**.

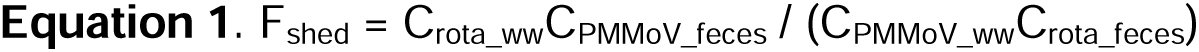

F_shed_ is the population fraction shedding rotavirus RNA in feces; C_rota_ww_ and C_PMMoV_ww_ are the concentration of rotavirus RNA and PMMoV RNA in wastewater solids, respectively; and C_PMMoV_feces_ and C_rota_feces_ are the concentration of PMMoV RNA and rotavirus RNA shed in feces, respectively. Arts et al.^26^ provide a large dataset on PMMoV RNA shedding in feces which we used to characterize the distribution of C_PMMoV_feces_. Zheng et al.^27^ compiled data on rotavirus RNA shedding in feces from infected individuals which we used to characterize the distribution of C_rota_feces_. Using these distributions in a Monte Carlo simulation, we modeled F_shed_ as a function of the PMMoV-normalized concentration of rotavirus RNA in wastewater solids (C_rota_ww_/C_PMMoV_ww_). We varied C_rota_ww_/C_PMMoV_ww_ as log_10_(C_rota_ww_/C_PMMoV_ww_) from -5 to -2 in 0.1 increments. For each C_rota_ww_/C_PMMoV_ww_, we conducted ten thousand simulations, randomly drawing C_PMMoV_feces_ and C_rota_feces_ from the distributions given by Arts et al.^26^ and Zheng et al.^27^, to estimate the median and IQR of F_shed_. We also conducted the Monte Carlo simulation to model F_shed_ each day using longitudinal measurements of C_rota_ww_/C_PMMoV_ww_ from an individual WWTP and spatially aggregated across all WWTPs to represent the United States.

We conducted a sensitivity analysis to evaluate the uncertainty of each model parameter. Following the method outlined by Xue et al.^83^, we determined the 25th, 50th, and 75th percentiles for each model input parameter (C_rota_ww_/C_PMMoV_ww_, C_PMMoV_feces_, C_rota_feces_). For C_rota_ww_/C_PMMoV_ww_, we determined percentile values based on nationally aggregated wastewater measurements. Then, holding all other parameters at their 50th percentile value, we calculated F_shed_ for each parameter twice: once at its 25th percentile value (F_shed,25_) and once at its 75th percentile value (F_shed,75_). We calculated the F_shed,75_:F_shed,25_ ratio for each input parameter. A ratio less than 1 indicates F_shed_ decreases as the input parameter increases, a ratio equal to 1 indicates F_shed_ is not affected by input parameter increases, and a ratio greater than 1 indicates F_shed_ increases as the input parameter increases (add other refs - see Sarah’s QMRA paper)

All calculations, analyses, and visualizations were conducted in R (version 4.1.3). This study was reviewed by the Stanford University Committee for the Protection of Human Subjects and determined to be exempt from oversight.

## Acknowledgments

We thank the participating wastewater treatment plant staff for collecting samples for this project. Data used in this study came from Epic Cosmos, a dataset created in collaboration with a community of Epic health systems representing more than 284 million patient records from over 1,500 hospitals and 36,000 clinics from all 50 states, D.C., Lebanon, and Saudi Arabia. We thank Dr. Rebecca Linfield for assistance with understanding the Epic Cosmos dataset. This work was supported by a gift to ABB from the Sergey Brin Family Foundation. The funder played no role in study design, data collection, analysis and interpretation of data, or the writing of this manuscript.

## Author contributions

**Elana M. G. Chan:** conceptualization, data curation, formal analysis, investigation, methodology, software, validation, visualization, writing - original draft, writing - review & editing; **Alessandro Zulli:** conceptualization, data curation, methodology, writing - review & editing; **Alexandria B Boehm:** conceptualization, data curation, funding acquisition, investigation, methodology, project administration, resources, supervision, validation, writing - original draft, writing - review & editing.

## Data availability

The datasets generated and analyzed for this study are available through the Stanford Digital Repository (https://doi.org/10.25740/mv671wb2446).

## Code availability

The underlying code for this study is available through the Stanford Digital Repository (https://doi.org/10.25740/mv671wb2446).

## Supplementary information

Rotavirus_SI.pdf

