## Supporting information for "Wastewater concentrations of rotavirus RNA are associated with infection and vaccination metrics in the USA"

---

Table of contents

PMMoV RNA concentrations: Wastewater treatment plants 2
Rotavirus RNA wastewater measurements and events: Wastewater treatment plants 10
Rotavirus RNA wastewater measurements and events: States 18
Correlation with vaccination coverage: State scale 20
Supplementary analysis: Comparing vaccination coverage estimates at the state scale 21
Distribution of population characteristics among sewersheds 22
Modeled $F_{shed}$ as a function of $C_{rota\_ww}/C_{PMMoV\_ww}$ 24
Supplementary analysis: Comparing clinical surveillance metrics at the national scale 25
Supplementary analysis: Correlation with clinical metrics at the state scale 26
Supplementary analysis: Rotavirus encounter diagnoses by age group 29
Wastewater monitoring: Further details 30
Rotavirus assay: Further details 45
Population characteristics: Further details 46
Spatial aggregation of wastewater monitoring data 49
Sewershed vaccination coverage: Further details 50
Mass balance model: Further details 51
References 52

PMMoV RNA concentrations: Wastewater treatment plants

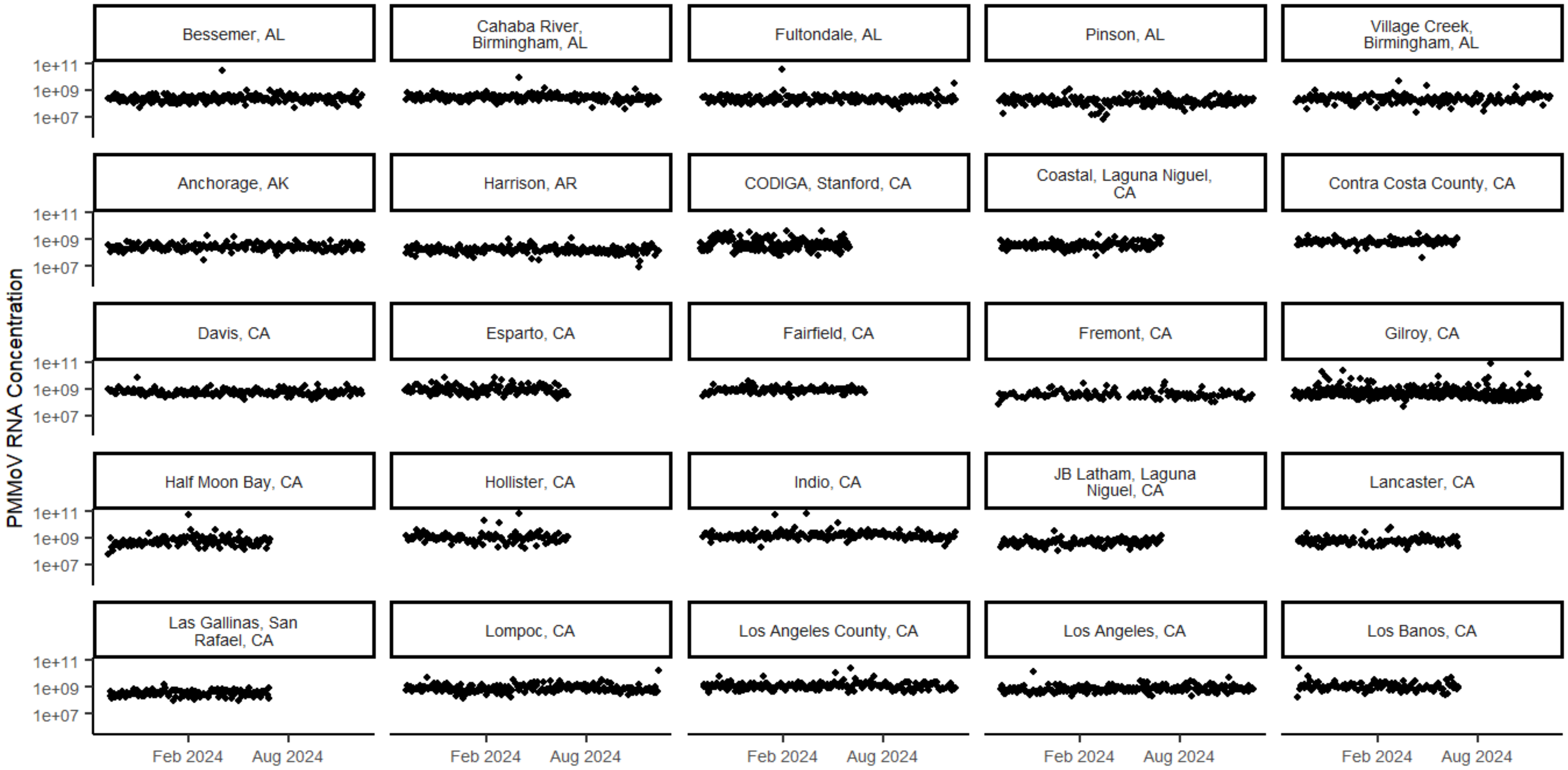

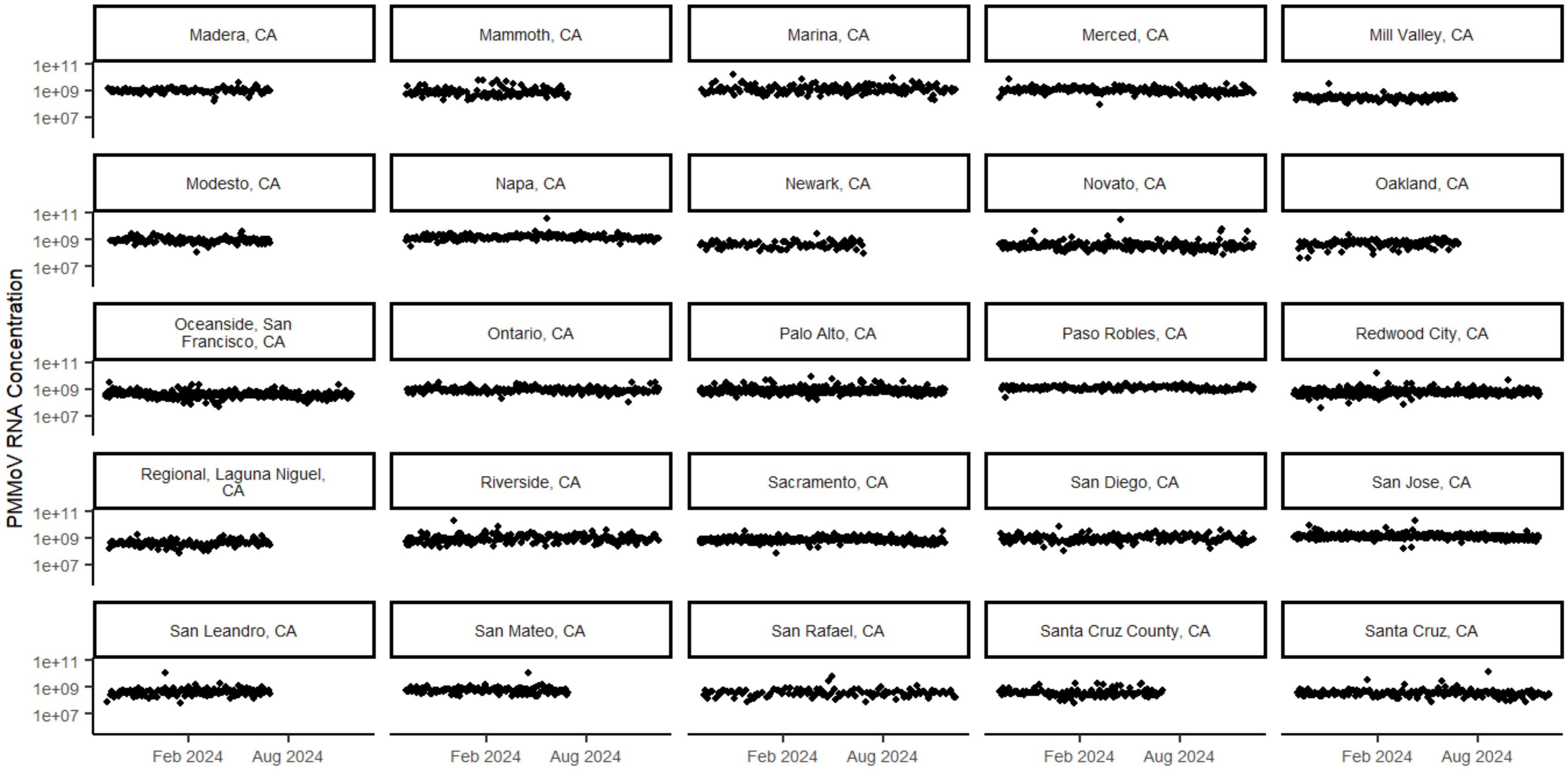

Madera, CA
Mammoth, CA
Marina, CA
Merced, CA
Mill Valley, CA
Modesto, CA
Napa, CA
Newark, CA
Novato, CA
Oakland, CA
Oceanside, San Francisco, CA
Ontario, CA
Palo Alto, CA
Paso Robles, CA
Redwood City, CA
Regional, Laguna Niguel, CA
Riverside, CA
Sacramento, CA
San Diego, CA
San Jose, CA
San Leandro, CA
San Mateo, CA
San Rafael, CA
Santa Cruz County, CA
Santa Cruz, CA
PMMoV RNA Concentration
1e+11
1e+09
1e+07
Feb 2024
Aug 2024

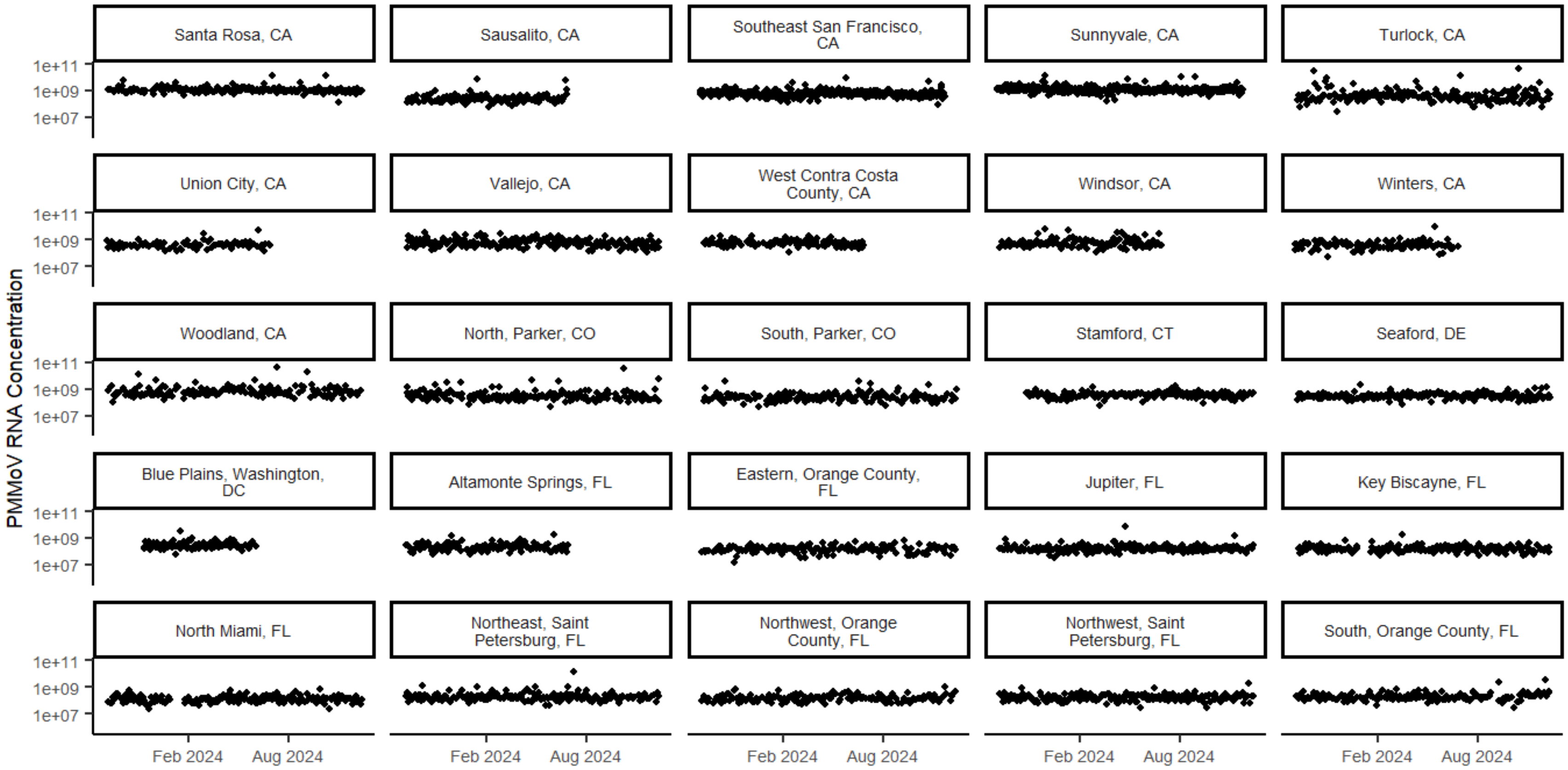

Santa Rosa, CA
Sausalito, CA
Southeast San Francisco, CA
Sunnyvale, CA
Turlock, CA
Union City, CA
Vallejo, CA
West Contra Costa County, CA
Windsor, CA
Winters, CA
Woodland, CA
North, Parker, CO
South, Parker, CO
Stamford, CT
Seaford, DE
Blue Plains, Washington, DC
Altamonte Springs, FL
Eastern, Orange County, FL
Jupiter, FL
Key Biscayne, FL
North Miami, FL
Northeast, Saint Petersburg, FL
Northwest, Orange County, FL
Northwest, Saint Petersburg, FL
South, Orange County, FL
PMMoV RNA Concentration
1e+11
1e+09
1e+07
Feb 2024
Aug 2024

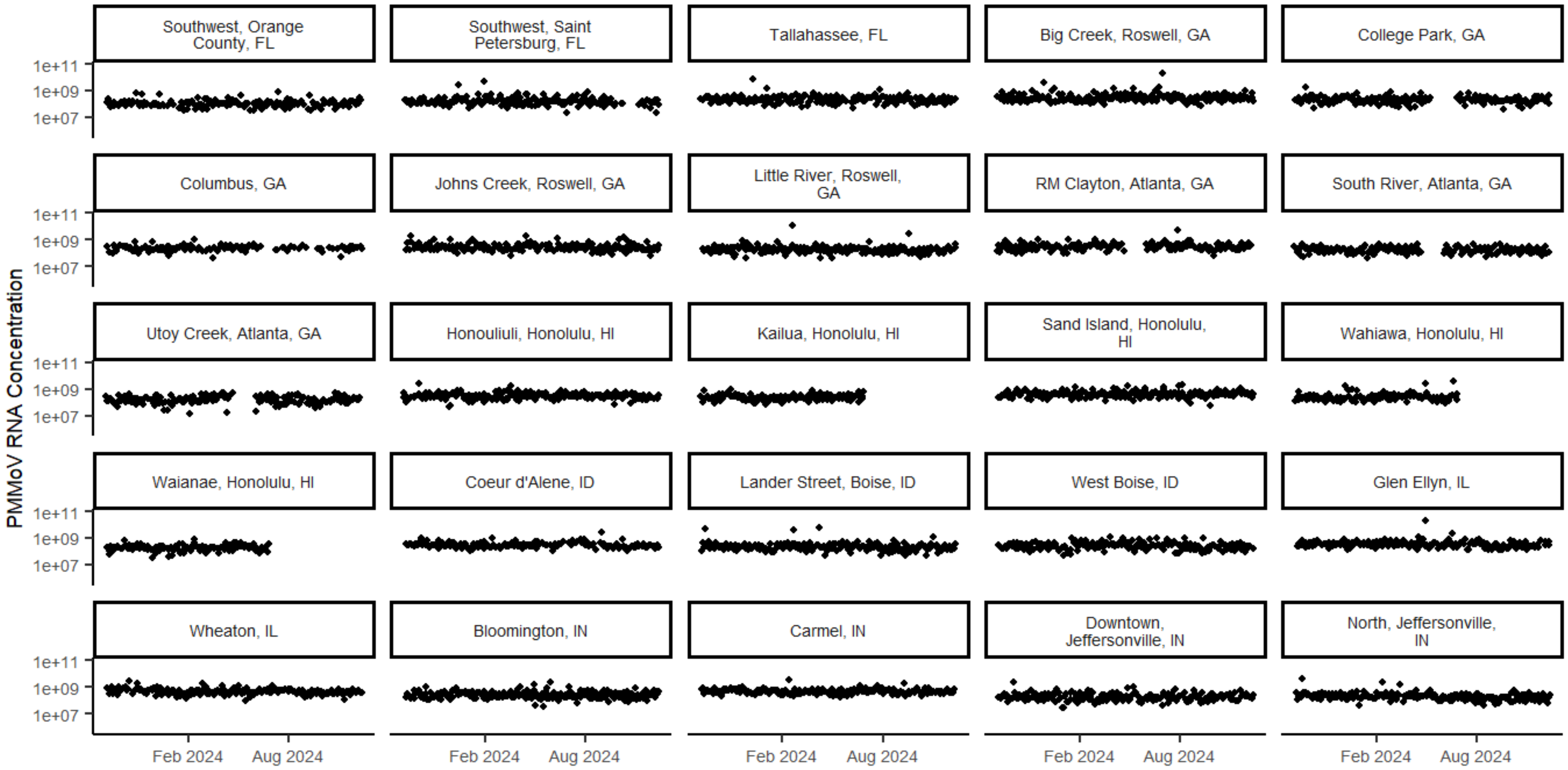

PMMoV RNA Concentration
Southwest, Orange County, FL
Southwest, Saint Petersburg, FL
Tallahassee, FL
Big Creek, Roswell, GA
College Park, GA
Columbus, GA
Johns Creek, Roswell, GA
Little River, Roswell, GA
RM Clayton, Atlanta, GA
South River, Atlanta, GA
Utoy Creek, Atlanta, GA
Honouliuli, Honolulu, HI
Kailua, Honolulu, HI
Sand Island, Honolulu, HI
Wahiawa, Honolulu, HI
Waianae, Honolulu, HI
Coeur d'Alene, ID
Lander Street, Boise, ID
West Boise, ID
Glen Ellyn, IL
Wheaton, IL
Bloomington, IN
Carmel, IN
Downtown, Jeffersonville, IN
North, Jeffersonville, IN
1e+11
1e+09
1e+07
Feb 2024
Aug 2024

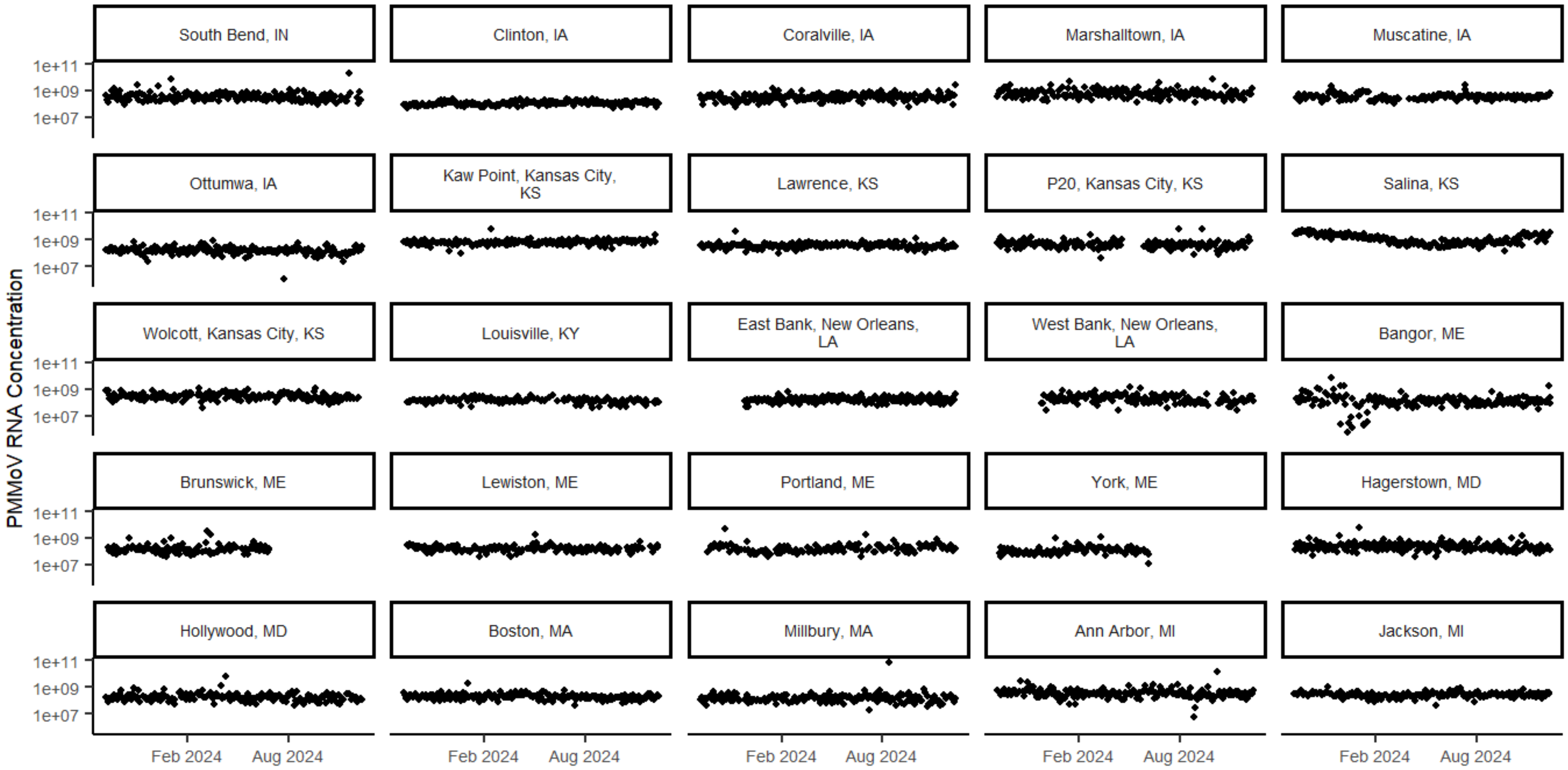

PMMoV RNA Concentration
South Bend, IN
Clinton, IA
Coralville, IA
Marshalltown, IA
Muscatine, IA
Ottumwa, IA
Kaw Point, Kansas City, KS
Lawrence, KS
P20, Kansas City, KS
Salina, KS
Wolcott, Kansas City, KS
Louisville, KY
East Bank, New Orleans, LA
West Bank, New Orleans, LA
Bangor, ME
Brunswick, ME
Lewiston, ME
Portland, ME
York, ME
Hagerstown, MD
Hollywood, MD
Boston, MA
Millbury, MA
Ann Arbor, MI
Jackson, MI
1e+11
1e+09
1e+07
Feb 2024
Aug 2024

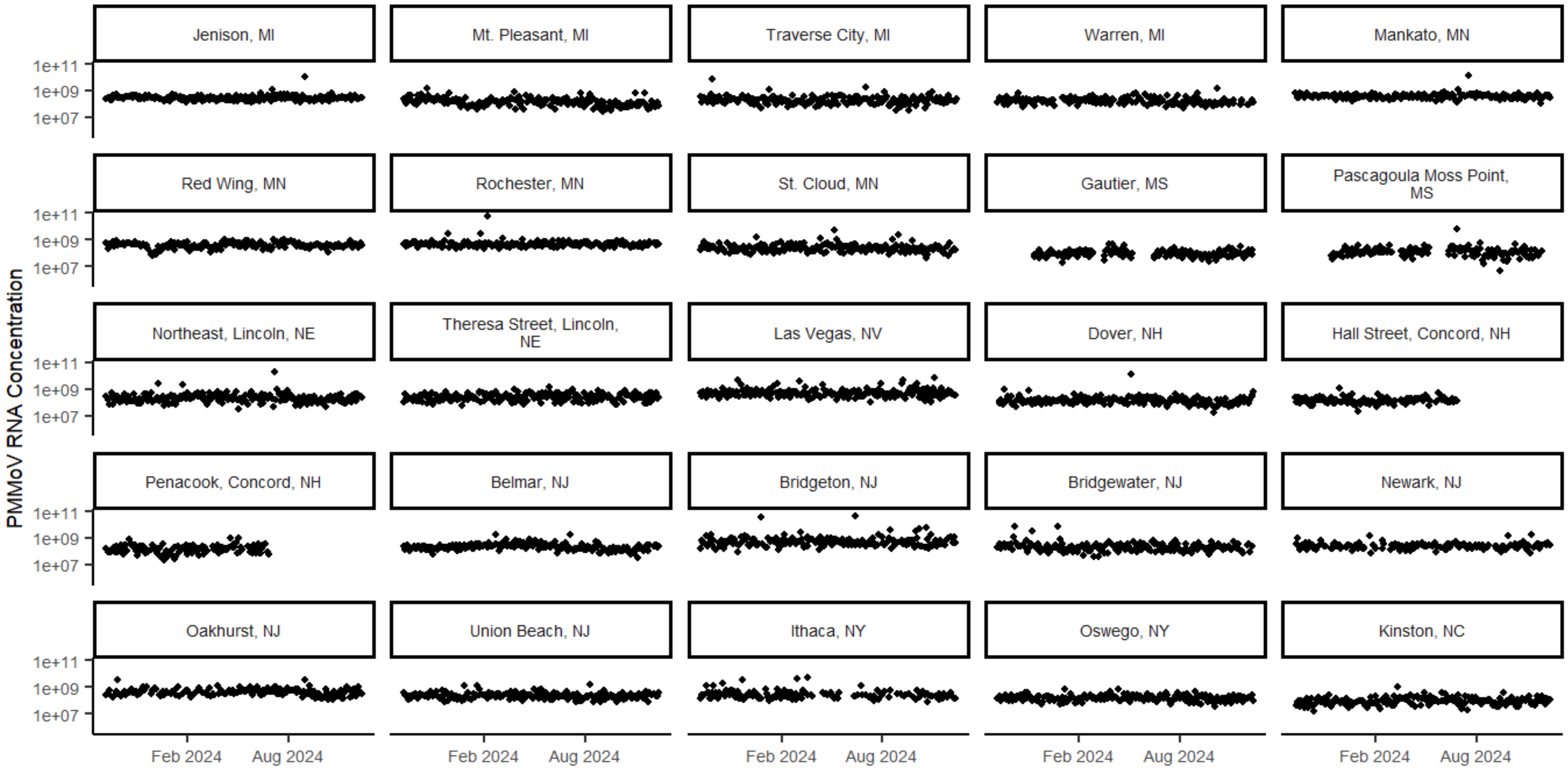

PMMoV RNA Concentration
Jenison, MI
Mt. Pleasant, MI
Traverse City, MI
Warren, MI
Mankato, MN
Red Wing, MN
Rochester, MN
St. Cloud, MN
Gautier, MS
Pascagoula Moss Point, MS
Northeast, Lincoln, NE
Theresa Street, Lincoln, NE
Las Vegas, NV
Dover, NH
Hall Street, Concord, NH
Penacook, Concord, NH
Belmar, NJ
Bridgeton, NJ
Bridgewater, NJ
Newark, NJ
Oakhurst, NJ
Union Beach, NJ
Ithaca, NY
Oswego, NY
Kinston, NC
1e+11
1e+09
1e+07
Feb 2024
Aug 2024

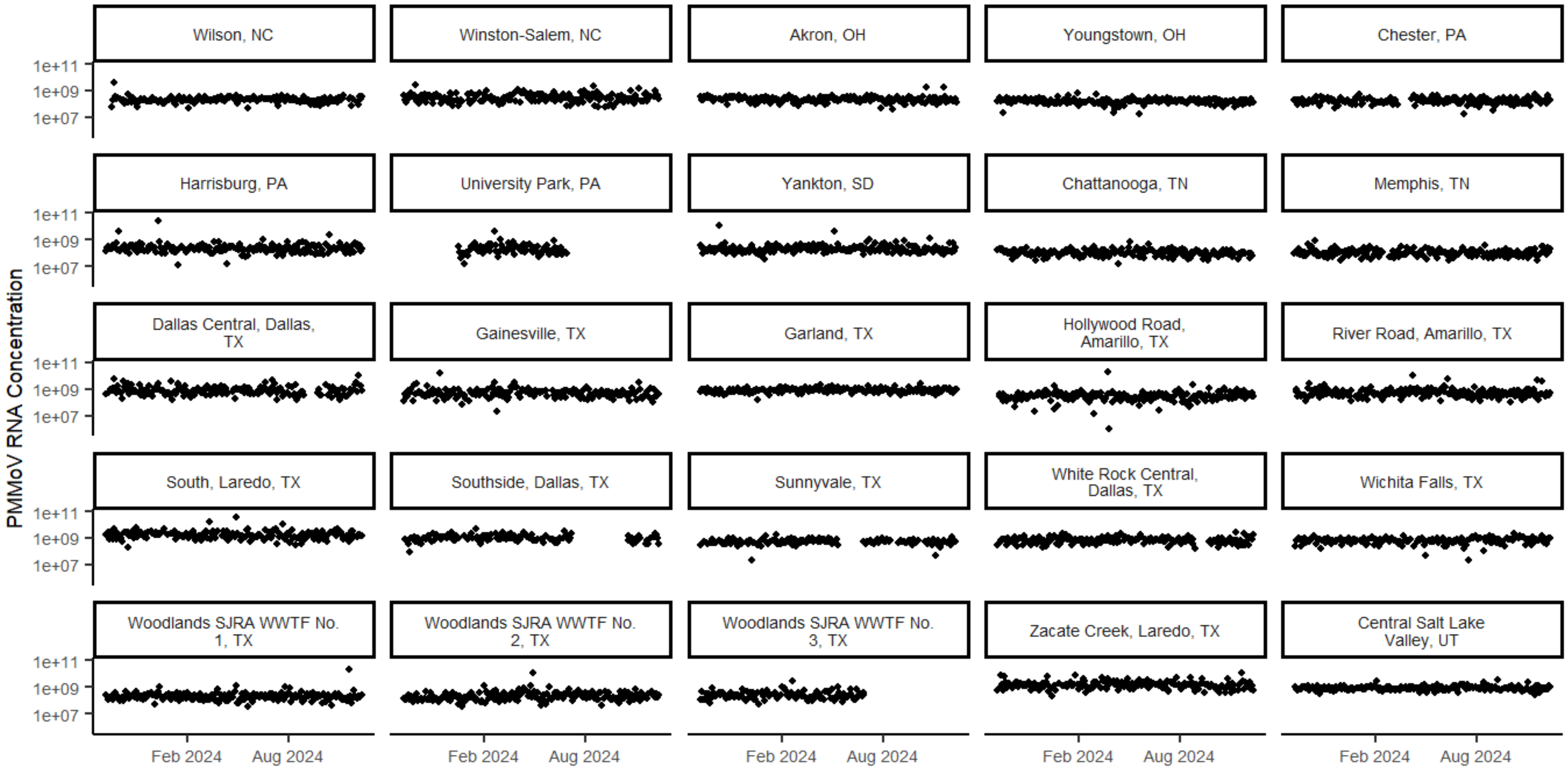

PMMoV RNA Concentration
Wilson, NC
Winston-Salem, NC
Akron, OH
Youngstown, OH
Chester, PA
Harrisburg, PA
University Park, PA
Yankton, SD
Chattanooga, TN
Memphis, TN
Dallas Central, Dallas, TX
Gainesville, TX
Garland, TX
Hollywood Road, Amarillo, TX
River Road, Amarillo, TX
South, Laredo, TX
Southside, Dallas, TX
Sunnyvale, TX
White Rock Central, Dallas, TX
Wichita Falls, TX
Woodlands SJRA WWTF No. 1, TX
Woodlands SJRA WWTF No. 2, TX
Woodlands SJRA WWTF No. 3, TX
Zacate Creek, Laredo, TX
Central Salt Lake Valley, UT
1e+11
1e+09
1e+07
Feb 2024
Aug 2024

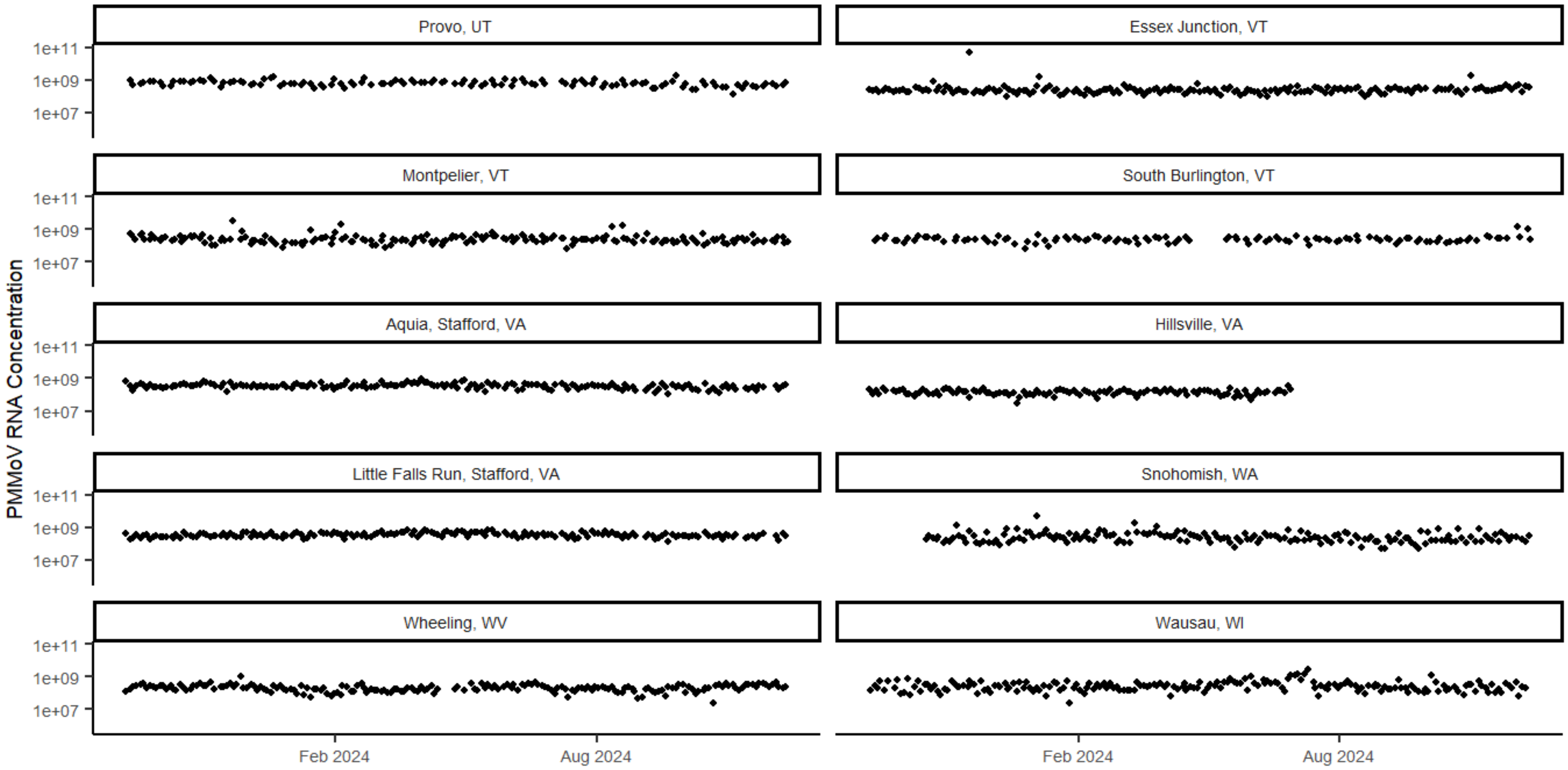


**Figure S1. PMMoV RNA concentrations in gc/g of wastewater solids measured in wastewater samples collected at WWTPs during the study period.** Concentrations are displayed on a $\log_{10}$ scale. WWTPs are ordered by state and then alphabetically within a state by WWTP location name. Abbreviations: gc/g = gene copies per gram, PMMoV = pepper mild mottle virus, WWTP = wastewater treatment plant.

### Rotavirus RNA wastewater measurements and events: Wastewater treatment plants

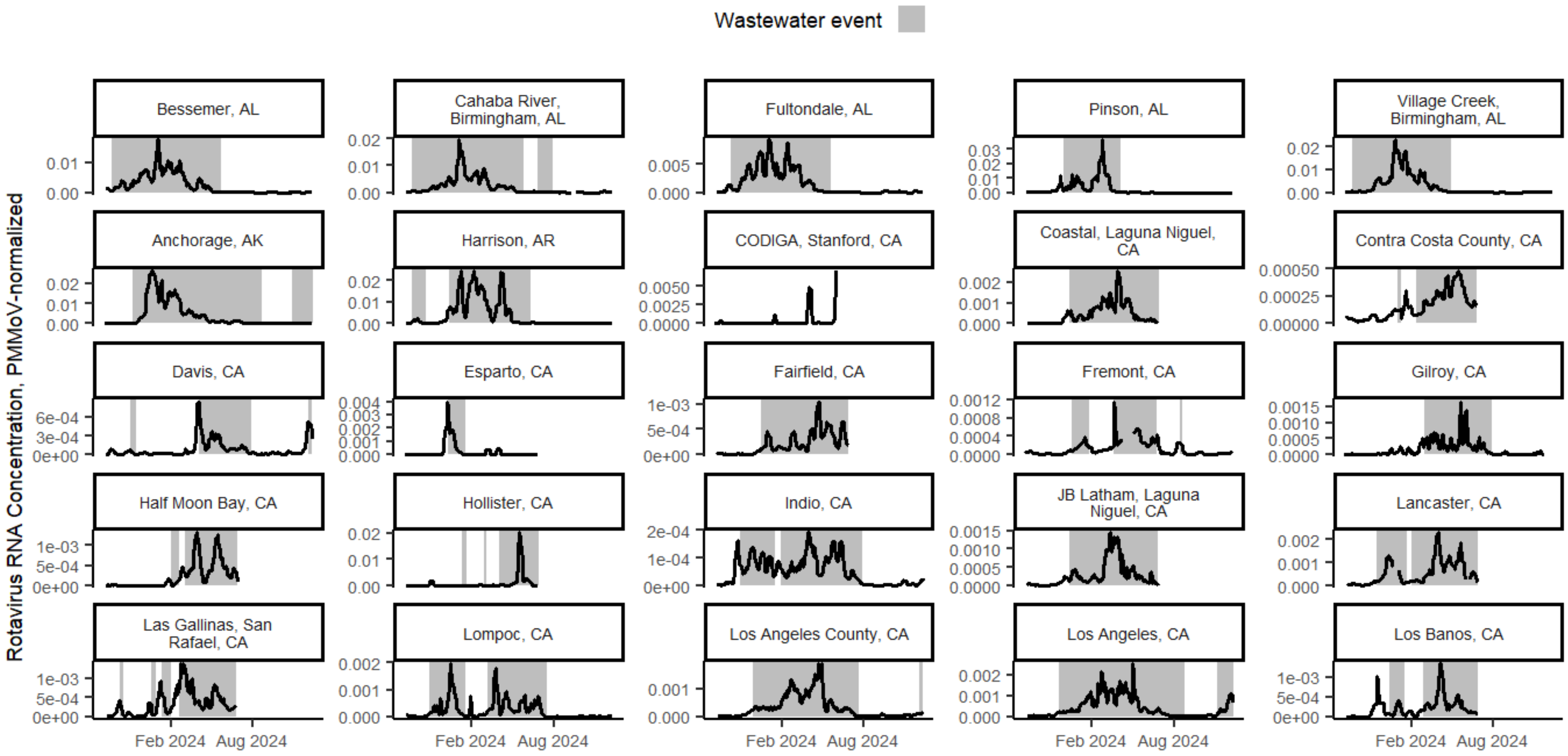

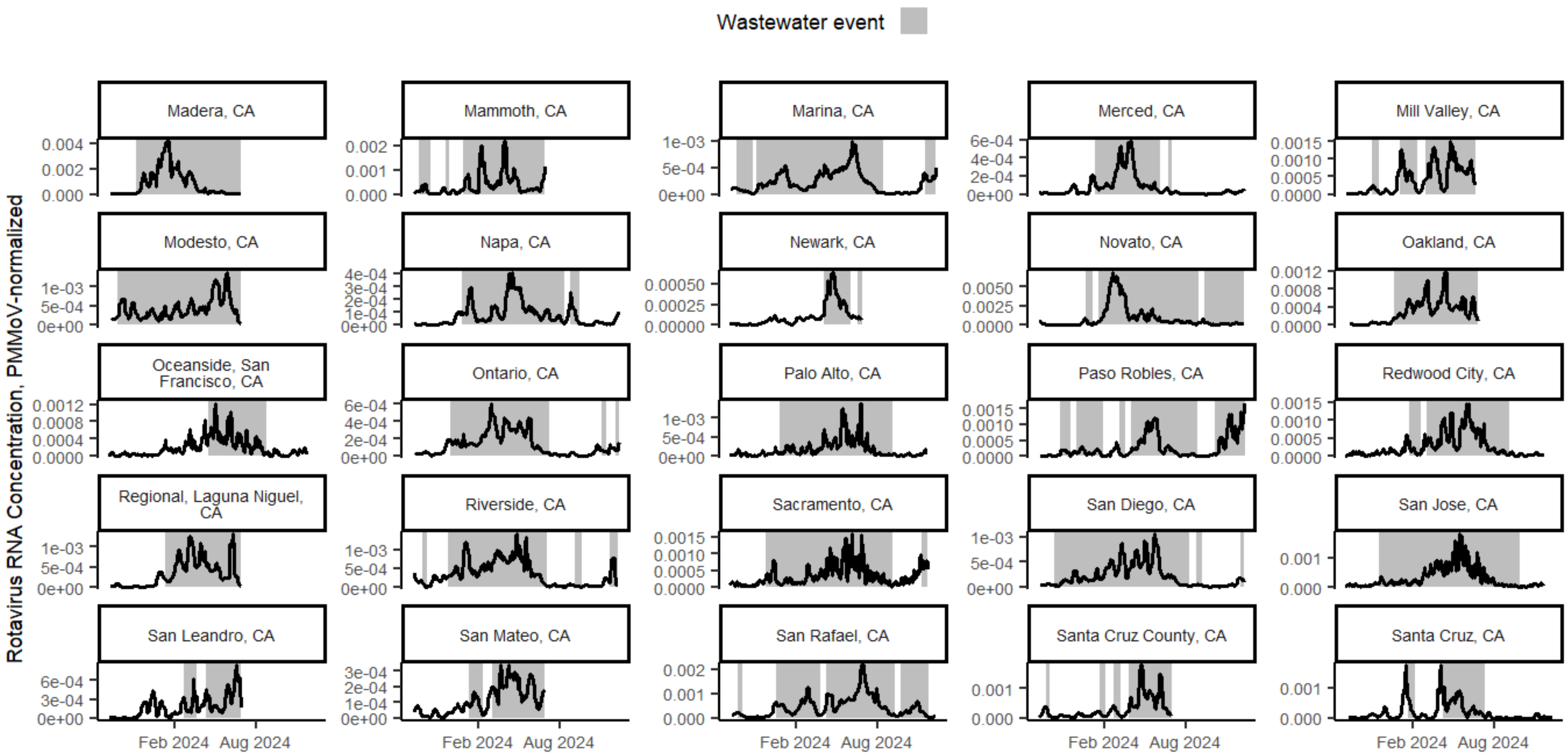

Wastewater event
Rotavirus RNA Concentration, PMMoV-normalized
Madera, CA
Mammoth, CA
Marina, CA
Merced, CA
Mill Valley, CA
Modesto, CA
Napa, CA
Newark, CA
Novato, CA
Oakland, CA
Oceanside, San Francisco, CA
Ontario, CA
Palo Alto, CA
Paso Robles, CA
Redwood City, CA
Regional, Laguna Niguel, CA
Riverside, CA
Sacramento, CA
San Diego, CA
San Jose, CA
San Leandro, CA
San Mateo, CA
San Rafael, CA
Santa Cruz County, CA
Santa Cruz, CA
Feb 2024
Aug 2024

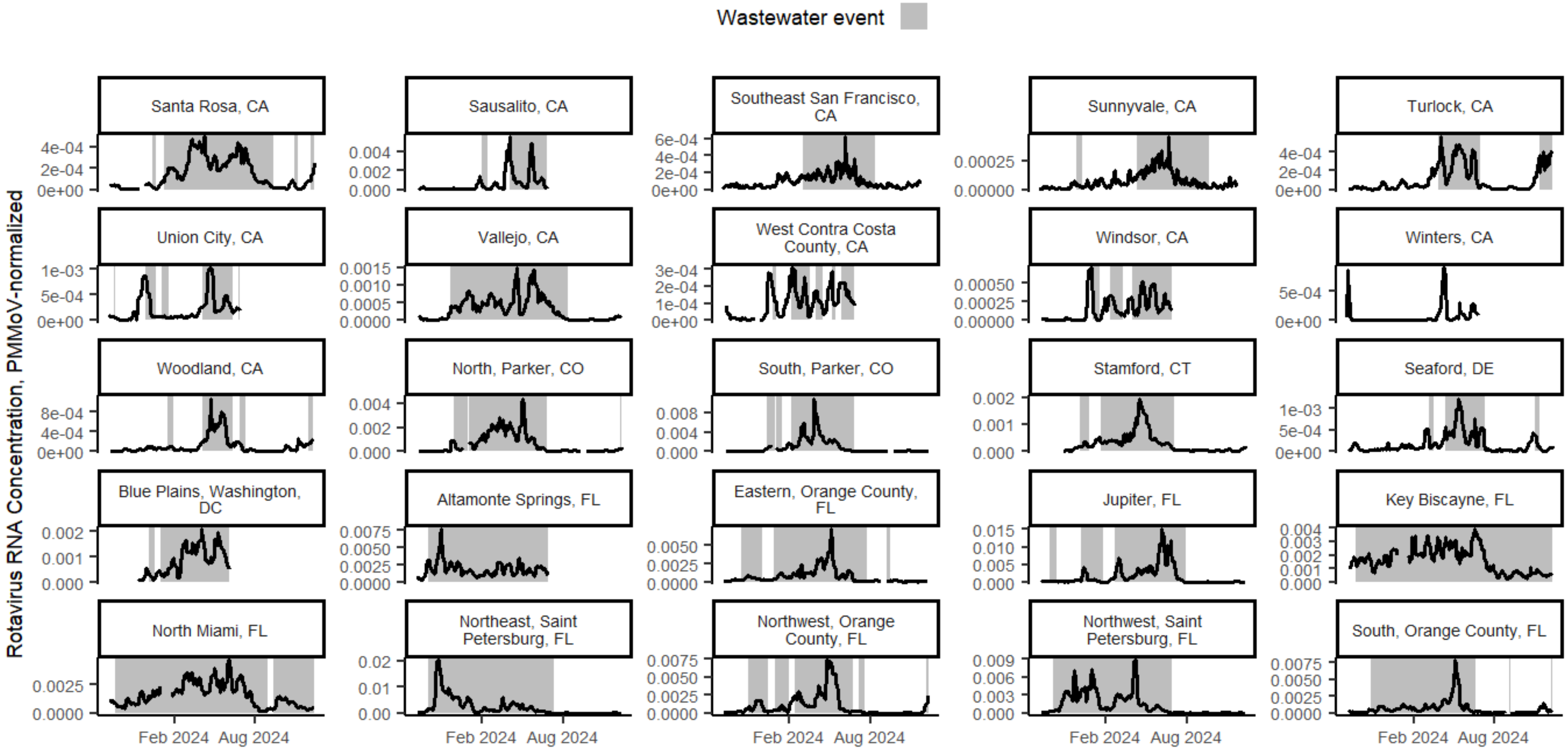
Wastewater event
Rotavirus RNA Concentration, PMMoV-normalized
Santa Rosa, CA
Sausalito, CA
Southeast San Francisco, CA
Sunnyvale, CA
Turlock, CA
Union City, CA
Vallejo, CA
West Contra Costa County, CA
Windsor, CA
Winters, CA
Woodland, CA
North, Parker, CO
South, Parker, CO
Stamford, CT
Seaford, DE
Blue Plains, Washington, DC
Altamonte Springs, FL
Eastern, Orange County, FL
Jupiter, FL
Key Biscayne, FL
North Miami, FL
Northeast, Saint Petersburg, FL
Northwest, Orange County, FL
Northwest, Saint Petersburg, FL
South, Orange County, FL
Feb 2024
Aug 2024

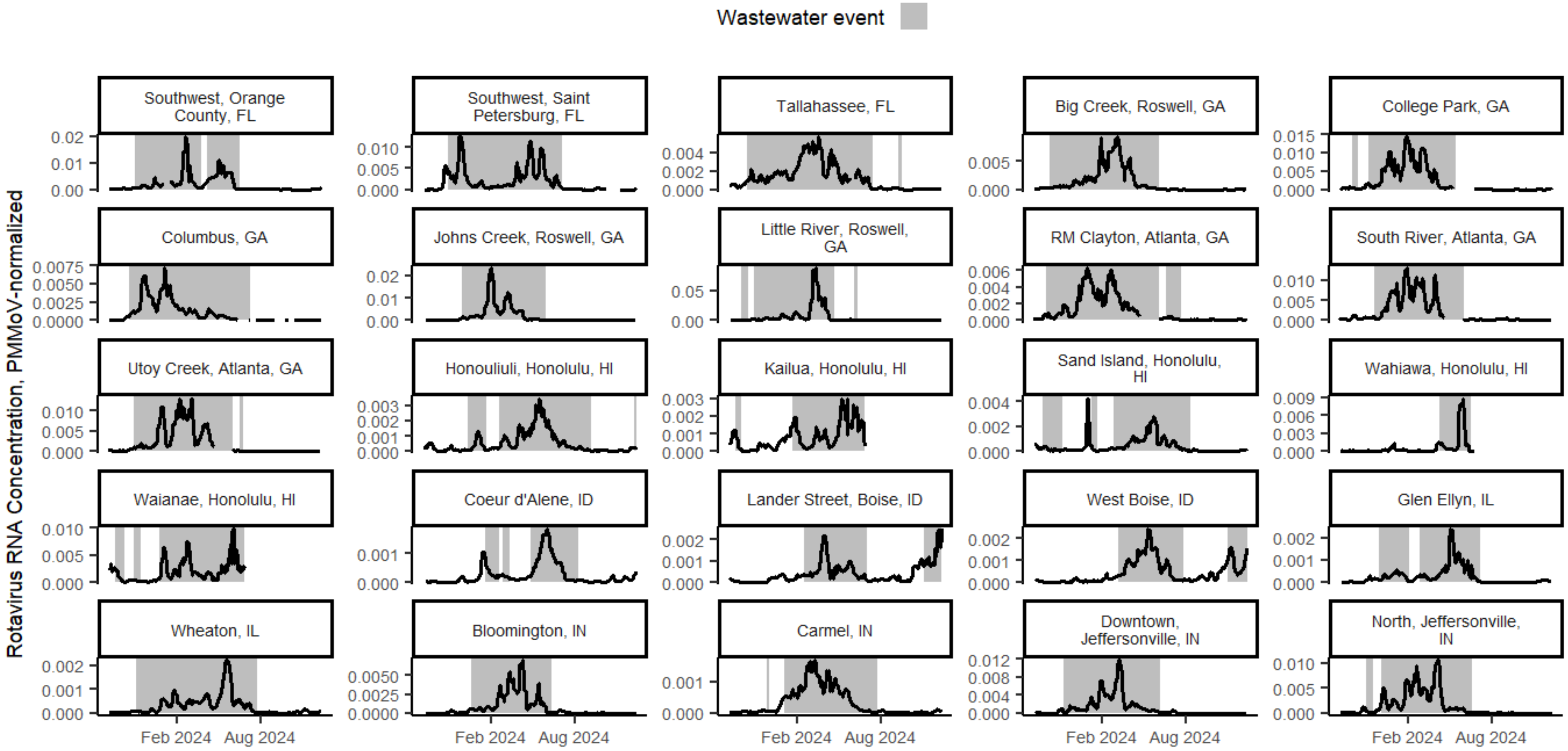
Wastewater event
Rotavirus RNA Concentration, PMMoV-normalized
Southwest, Orange County, FL
Southwest, Saint Petersburg, FL
Tallahassee, FL
Big Creek, Roswell, GA
College Park, GA
Columbus, GA
Johns Creek, Roswell, GA
Little River, Roswell, GA
RM Clayton, Atlanta, GA
South River, Atlanta, GA
Utoy Creek, Atlanta, GA
Honouliuli, Honolulu, HI
Kailua, Honolulu, HI
Sand Island, Honolulu, HI
Wahiawa, Honolulu, HI
Waianae, Honolulu, HI
Coeur d'Alene, ID
Lander Street, Boise, ID
West Boise, ID
Glen Ellyn, IL
Wheaton, IL
Bloomington, IN
Carmel, IN
Downtown, Jeffersonville, IN
North, Jeffersonville, IN
Feb 2024
Aug 2024

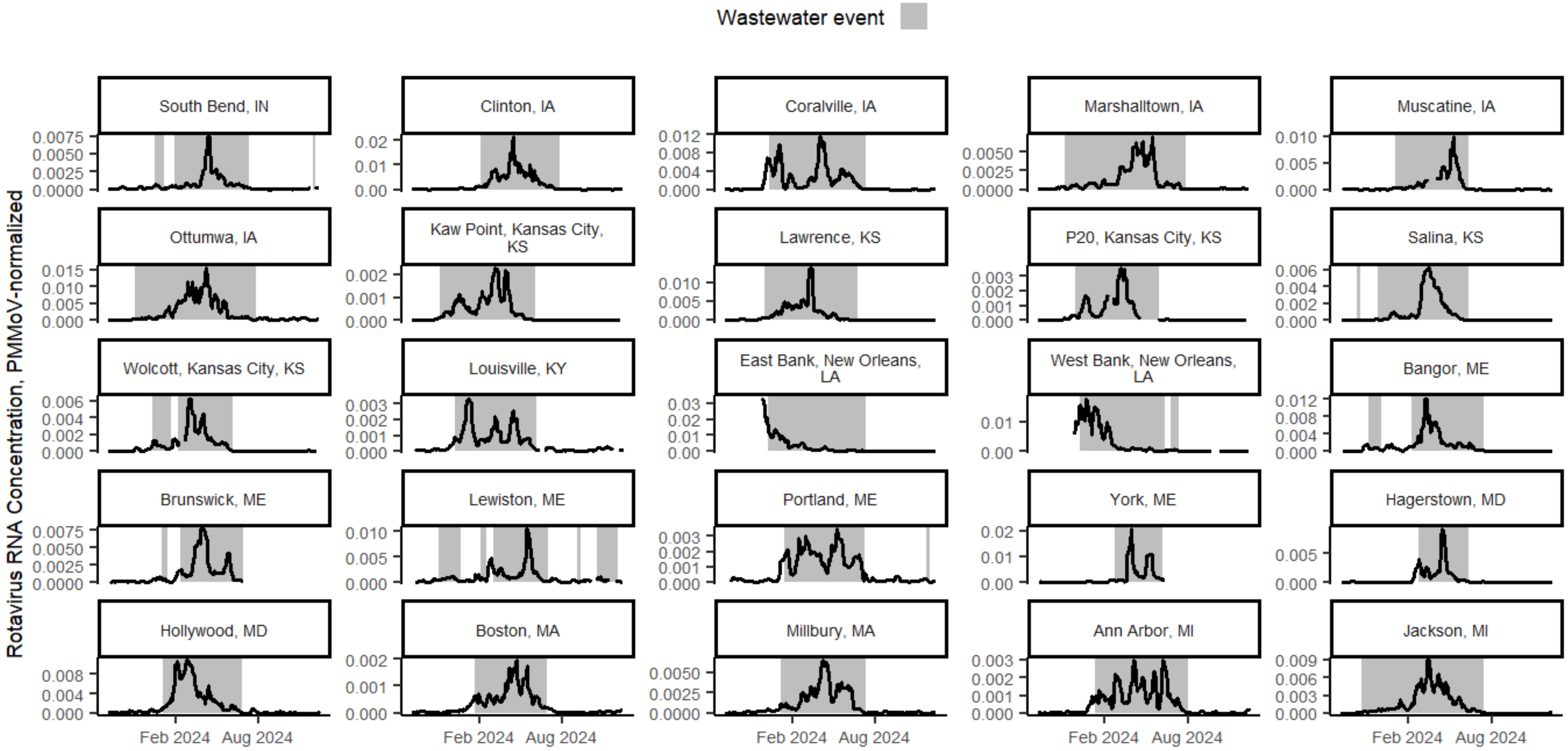
Wastewater event
Rotavirus RNA Concentration, PMMoV-normalized
South Bend, IN
Clinton, IA
Coralville, IA
Marshalltown, IA
Muscatine, IA
Ottumwa, IA
Kaw Point, Kansas City, KS
Lawrence, KS
P20, Kansas City, KS
Salina, KS
Wolcott, Kansas City, KS
Louisville, KY
East Bank, New Orleans, LA
West Bank, New Orleans, LA
Bangor, ME
Brunswick, ME
Lewiston, ME
Portland, ME
York, ME
Hagerstown, MD
Hollywood, MD
Boston, MA
Millbury, MA
Ann Arbor, MI
Jackson, MI
Feb 2024
Aug 2024

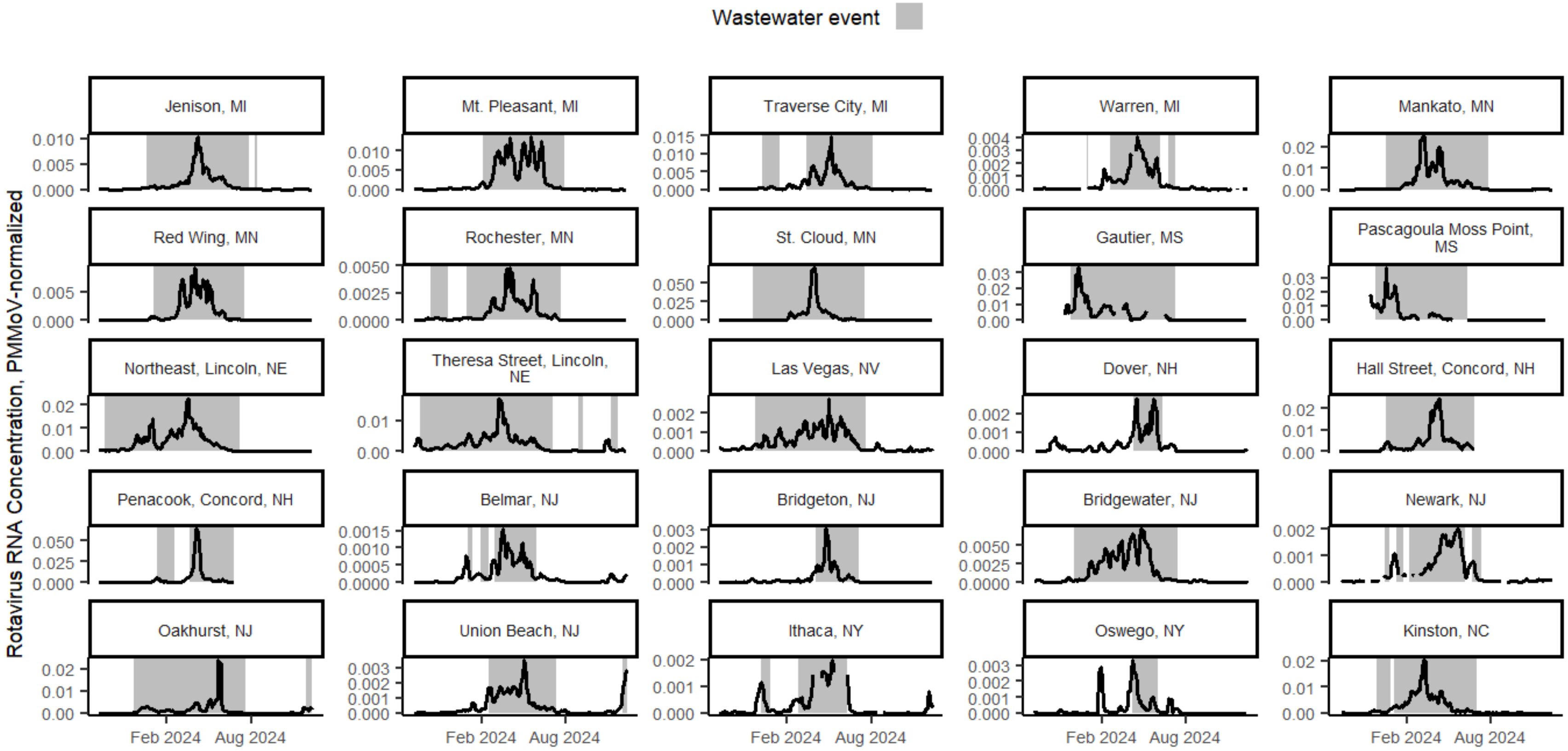
Wastewater event
Rotavirus RNA Concentration, PMMoV-normalized
Jenison, MI
Mt. Pleasant, MI
Traverse City, MI
Warren, MI
Mankato, MN
Red Wing, MN
Rochester, MN
St. Cloud, MN
Gautier, MS
Pascagoula Moss Point, MS
Northeast, Lincoln, NE
Theresa Street, Lincoln, NE
Las Vegas, NV
Dover, NH
Hall Street, Concord, NH
Penacook, Concord, NH
Belmar, NJ
Bridgeton, NJ
Bridgewater, NJ
Newark, NJ
Oakhurst, NJ
Union Beach, NJ
Ithaca, NY
Oswego, NY
Kinston, NC
Feb 2024
Aug 2024

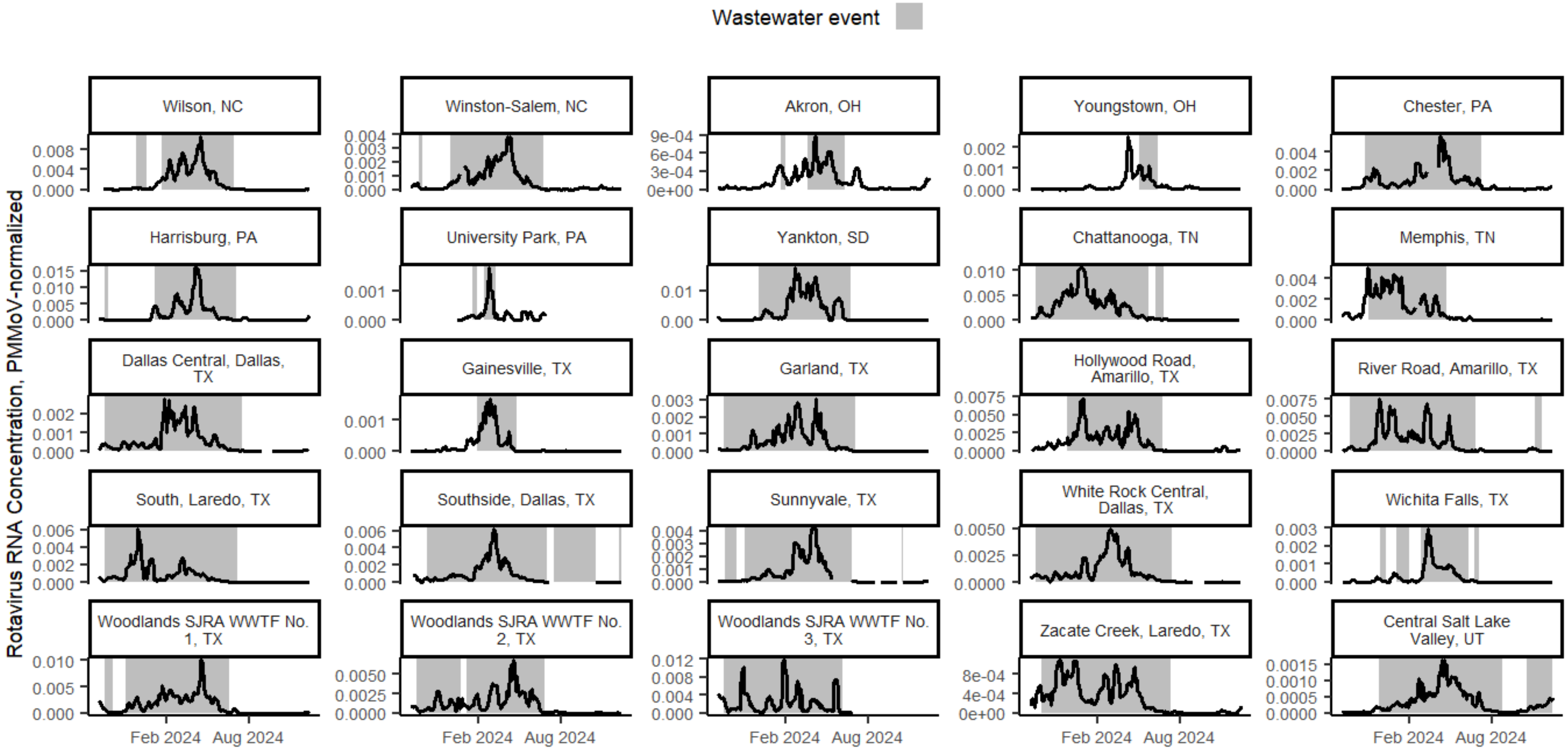

Wastewater event
Rotavirus RNA Concentration, PMMoV-normalized
Wilson, NC
Winston-Salem, NC
Akron, OH
Youngstown, OH
Chester, PA
Harrisburg, PA
University Park, PA
Yankton, SD
Chattanooga, TN
Memphis, TN
Dallas Central, Dallas, TX
Gainesville, TX
Garland, TX
Hollywood Road, Amarillo, TX
River Road, Amarillo, TX
South, Laredo, TX
Southside, Dallas, TX
Sunnyvale, TX
White Rock Central, Dallas, TX
Wichita Falls, TX
Woodlands SJRA WWTF No. 1, TX
Woodlands SJRA WWTF No. 2, TX
Woodlands SJRA WWTF No. 3, TX
Zacate Creek, Laredo, TX
Central Salt Lake Valley, UT
Feb 2024
Aug 2024

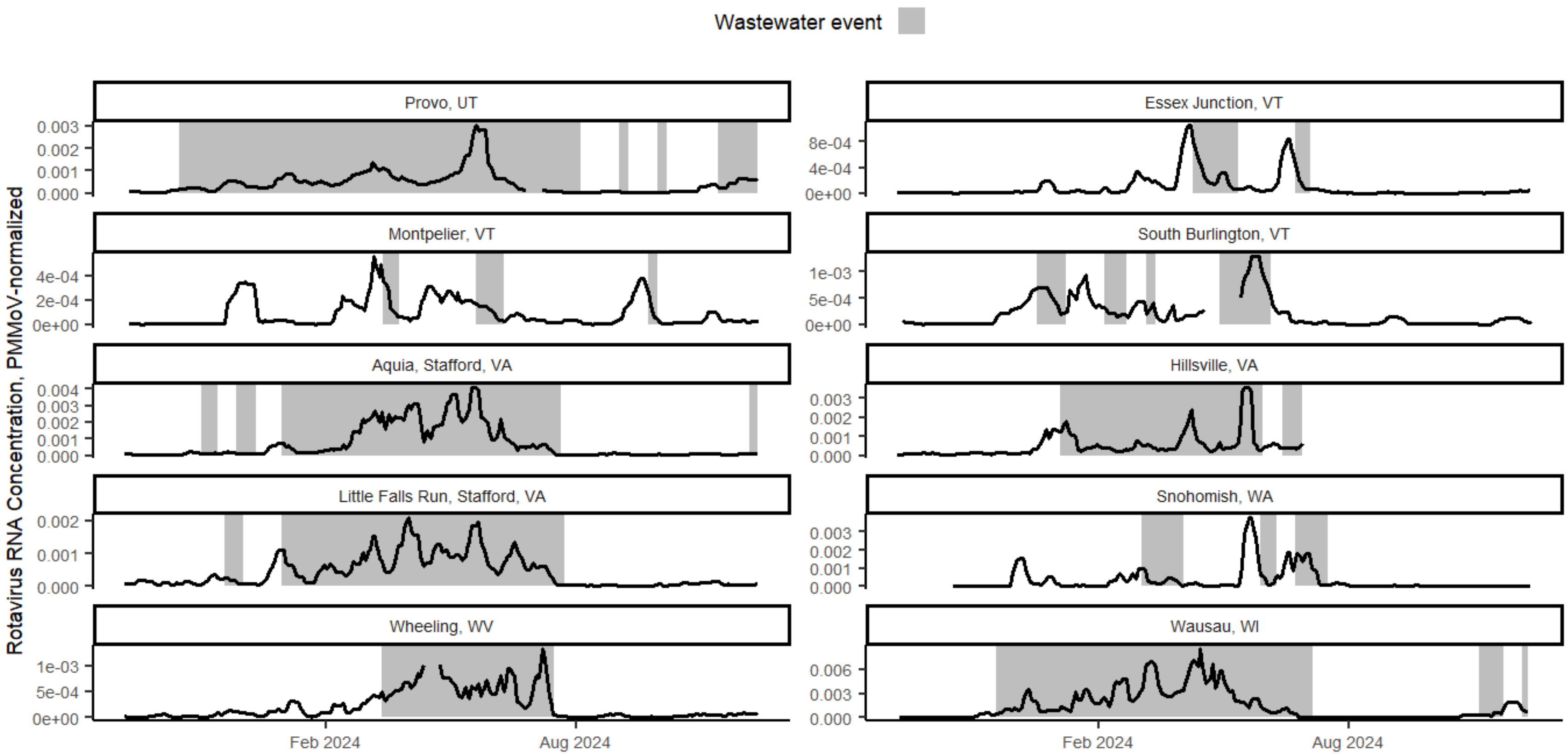


**Figure S2. Smoothed PMMoV-normalized rotavirus RNA wastewater concentrations with wastewater events shaded for each WWTP.** Linear interpolation was used between adjacent samples to generate daily time series (no interpolation if >10 day gap in sampling). WWTPs are ordered by state and then alphabetically within a state by WWTP location name. Abbreviations: PMMoV = pepper mild mottle virus, WWTP = wastewater treatment plant.

### Rotavirus RNA wastewater measurements and events: States

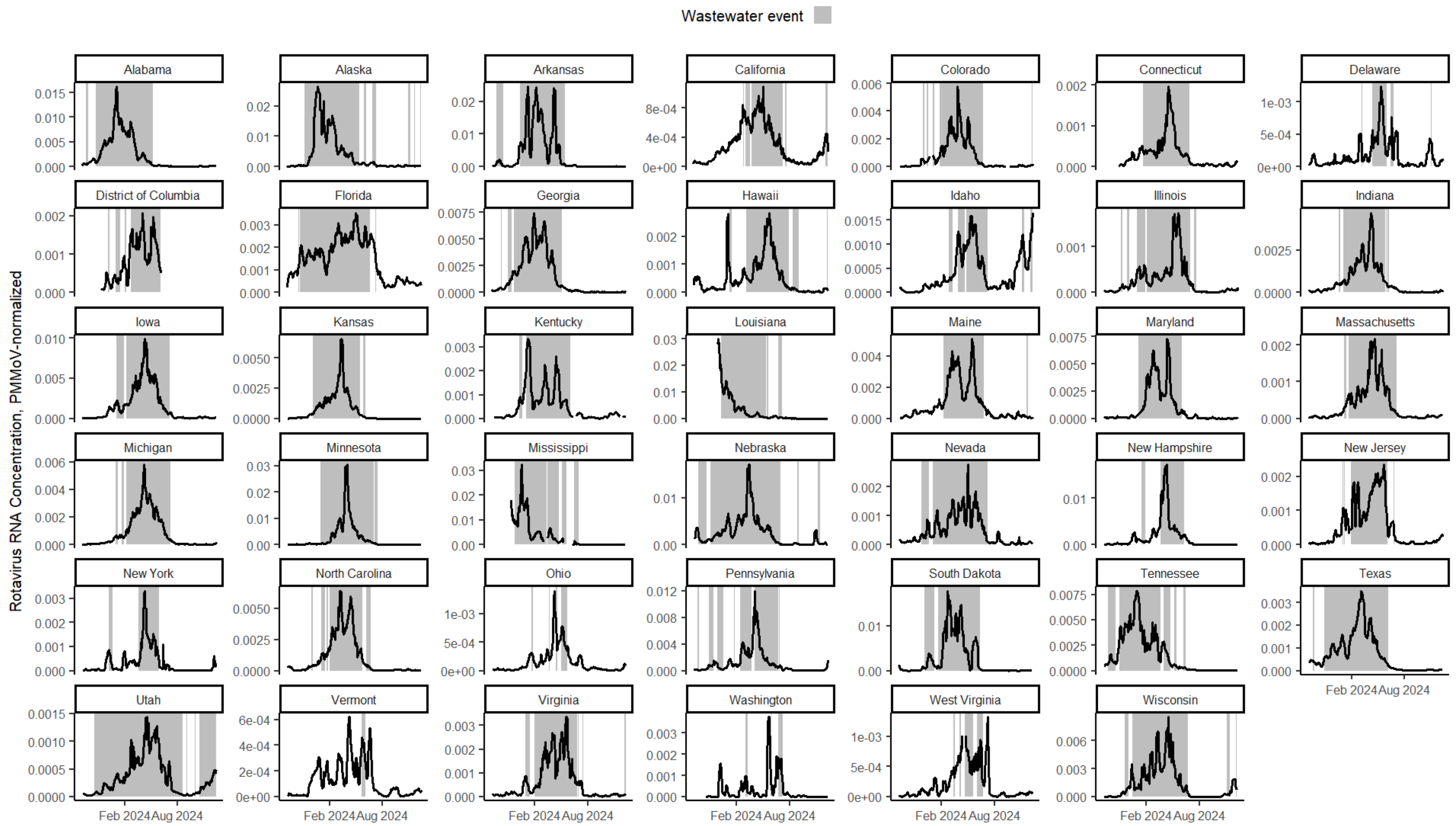


**Figure S3. State aggregated smoothed wastewater rotavirus RNA concentrations normalized by PMMoV with the rotavirus wastewater event shaded for each state.** Abbreviation: PMMoV = pepper mild mottle virus.

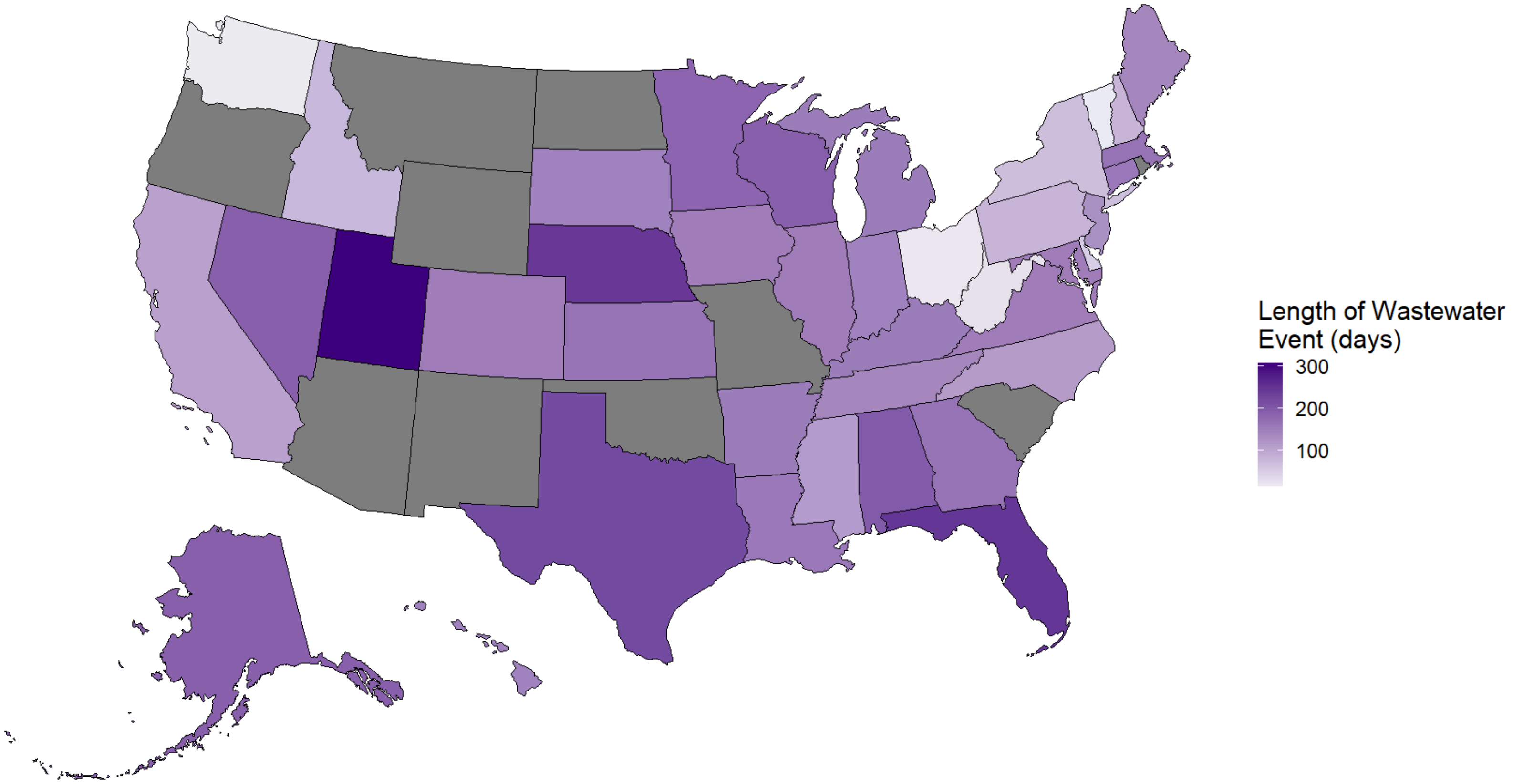


**Figure S4. Wastewater event duration in days among states.** Gray states did not have any participating wastewater treatment plants included in the study.

#### Correlation with vaccination coverage: State scale

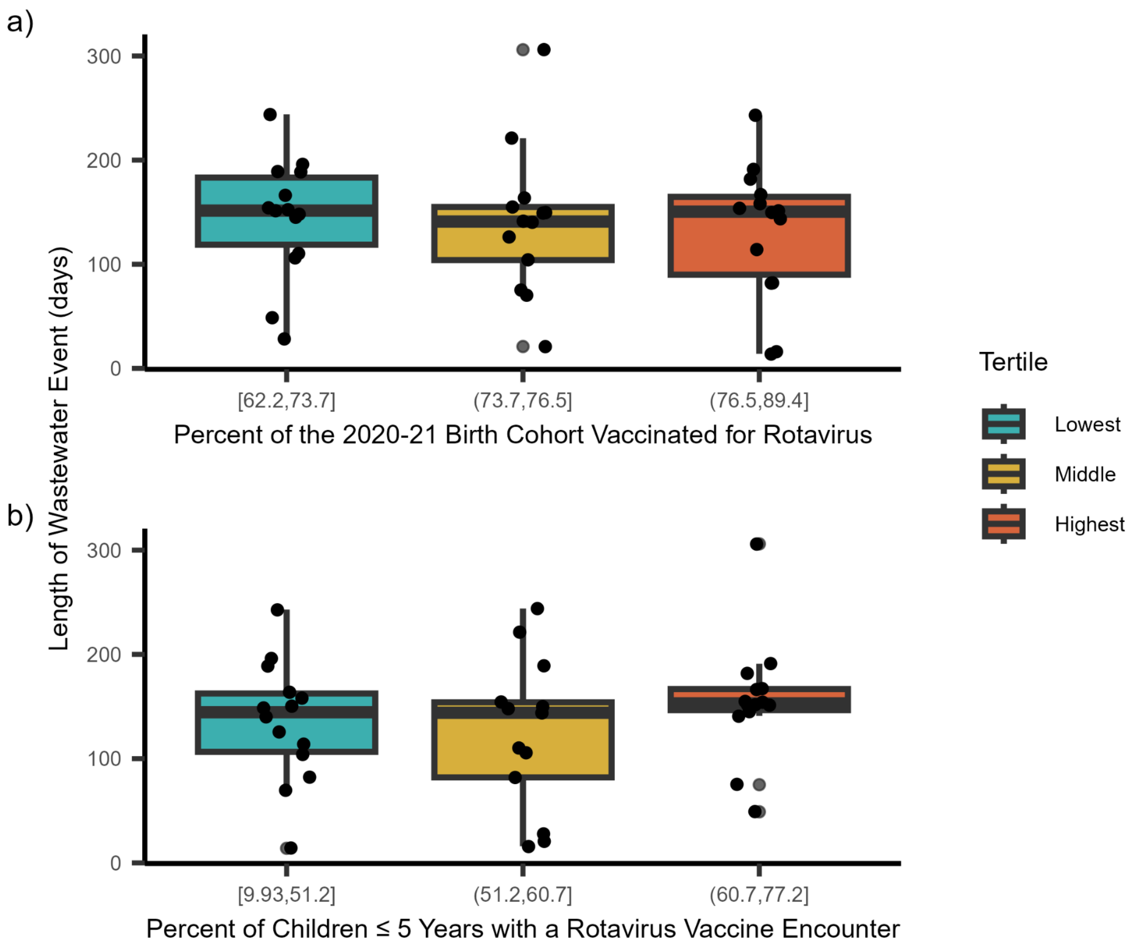


**Figure S5. Rotavirus wastewater event durations among states by tertile of rotavirus vaccination coverage.** Two estimates of rotavirus vaccination coverage were used to determine tertiles: (**a**) percent of the 2020-21 birth cohort in the state vaccinated for rotavirus from the National Center for Immunization and Respiratory Diseases and (**b**) percent of children ≤ 5 years in the state with a rotavirus vaccine encounter from Epic Cosmos. The center line represents the median, the box limits represent the 25th and 75th percentiles, and the whiskers represent 1.5 x IQR from the box limits. Jittered points represent individual data points. Abbreviations: IQR = interquartile range.

#### Supplementary analysis: Comparing vaccination coverage estimates at the state scale

Vaccination coverage estimates from both NCIRD and Epic Cosmos are normally distributed (Shapiro-Wilk test, $p > 0.05$). Rotavirus vaccination coverage from NCIRD and Epic Cosmos are not correlated (Pearson's correlation, $p = 0.95$) (**Figure S6**).

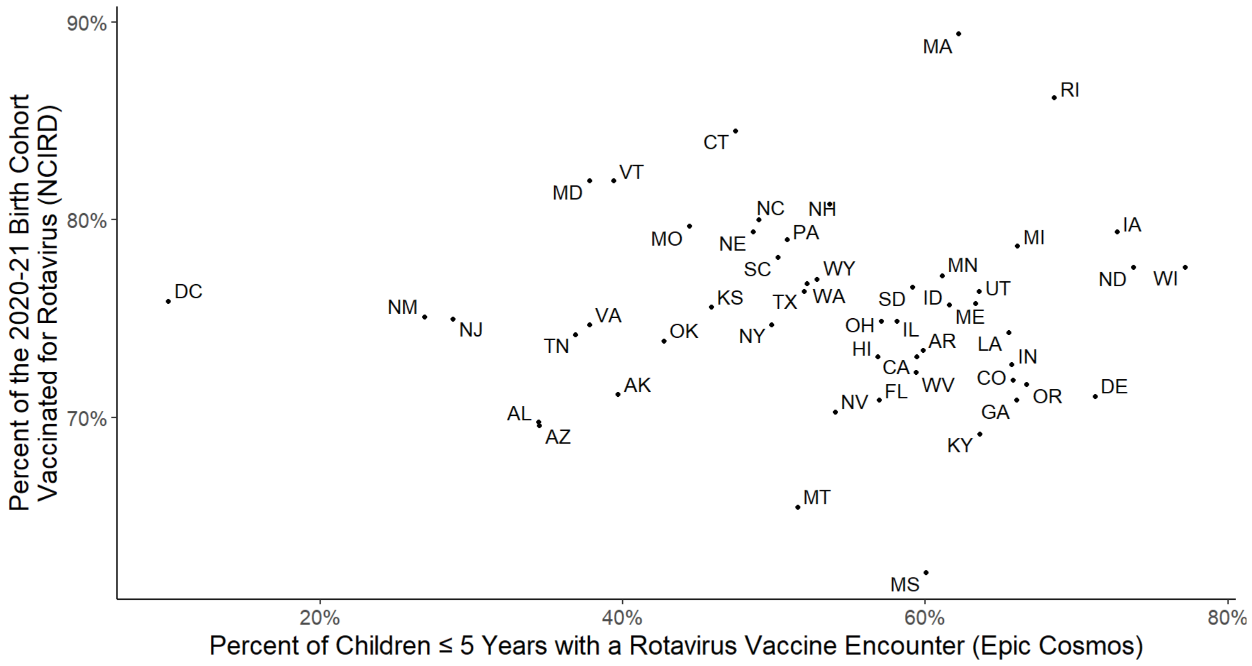


**Figure S6. State-level rotavirus vaccination coverage estimates obtained from NCIRD versus Epic Cosmos.** Abbreviation: NCIRD = National Center for Immunization and Respiratory Diseases.

#### Distribution of population characteristics among sewersheds

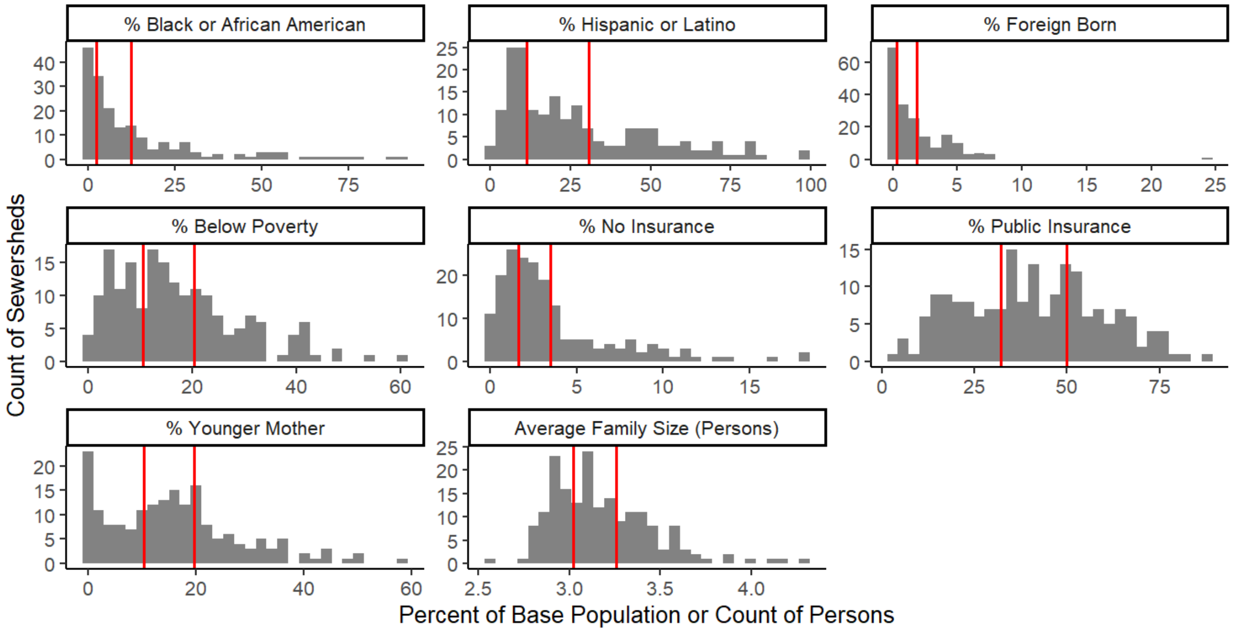


**Figure S7. Distribution of each population characteristic derived from the American Community Survey among sewersheds.** For average family size, the x-axis represents the count of persons. For all other characteristics, the x-axis represents the percent of the base population; refer to **Table S3** for a description of the base population. Vertical, red lines indicate tertile cutoffs for each characteristic.

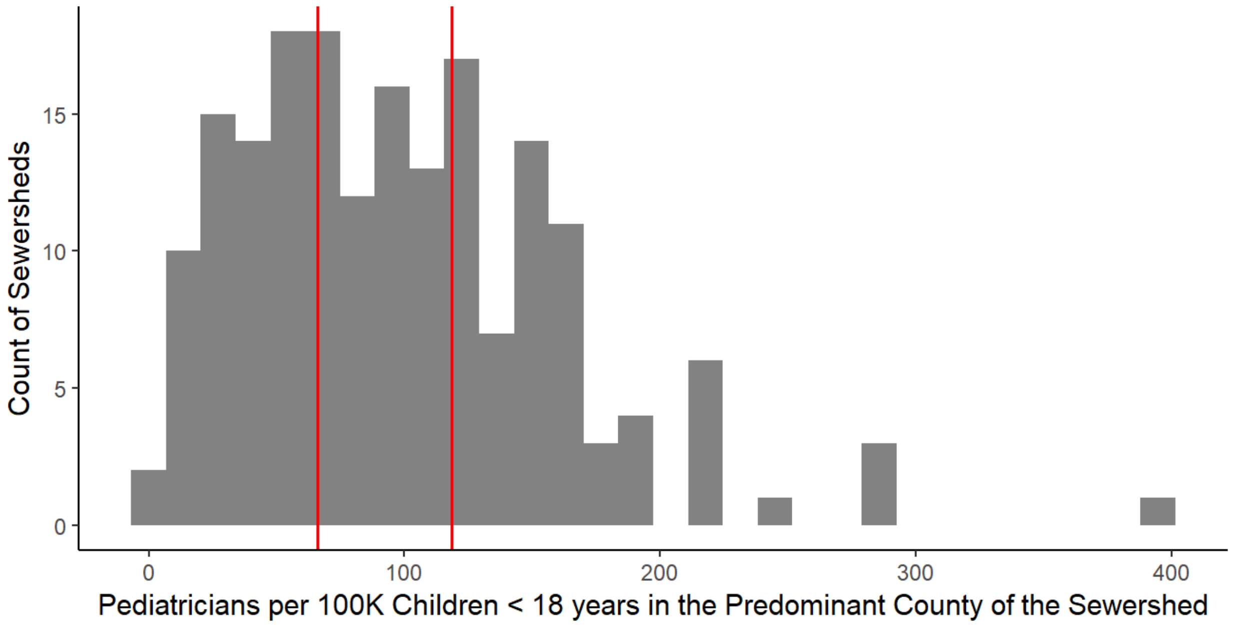


**Figure S8. Distribution of the pediatrician:child ratio (i.e., number of pediatricians per 100 thousand children < 18 years) among sewersheds.** Vertical, red lines indicate tertile cutoffs.

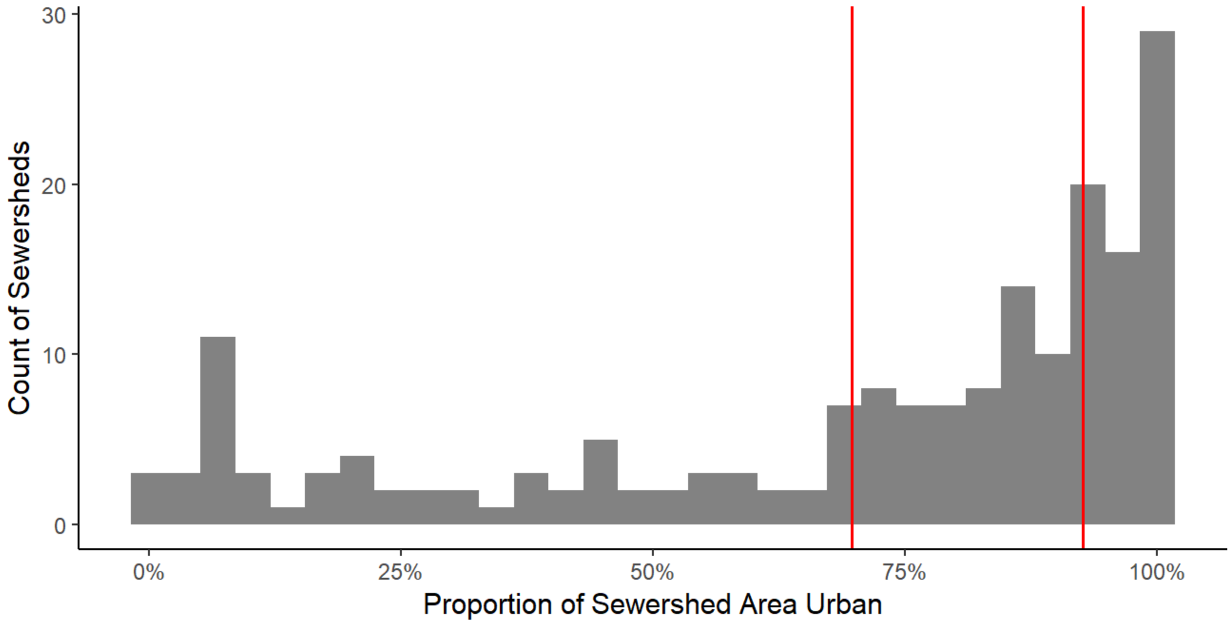


**Figure S9. Distribution of the urban area proportion among sewersheds.** Vertical, red lines indicate tertile cutoffs.

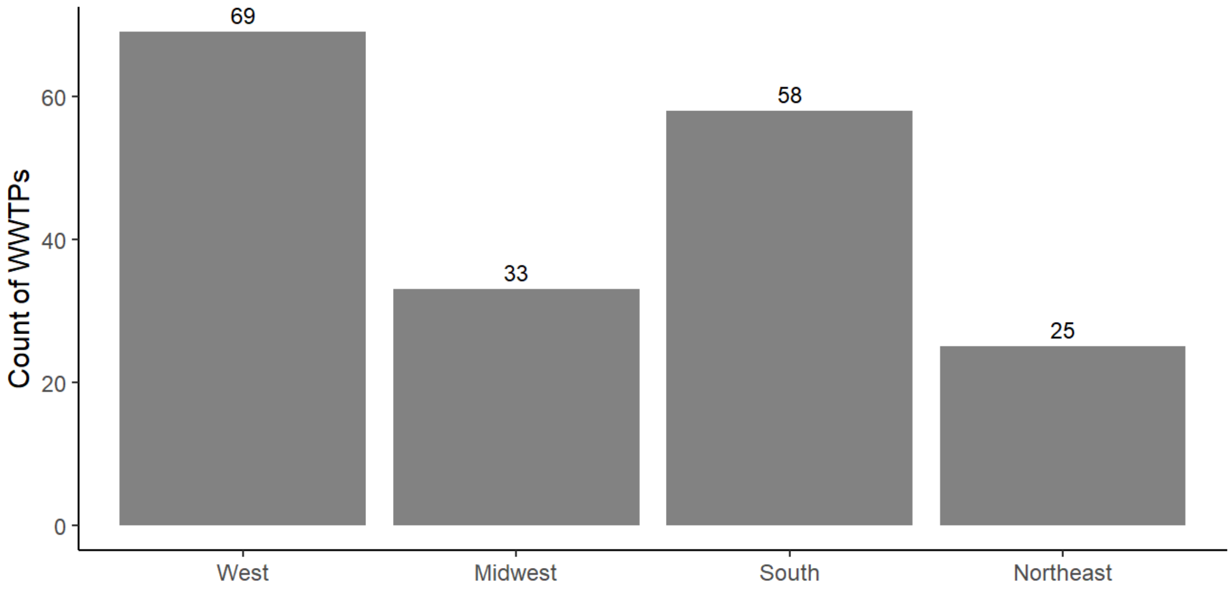


**Figure S10. Count of wastewater treatment plants (WWTPS) located in each US Census Bureau region.**

### Modeled $F_{shed}$ as a function of $C_{rota\_ww}/C_{PMMoV\_ww}$

Refer to **Equation 1** in the main text for an overview of the model and Monte Carlo simulation (**Figure S11**). We characterized the distributions of $C_{PMMoV\_feces}$ and $C_{rota\_feces}$ using data provided by Arts et al.[1] and Zheng et al.[2], respectively. Arts et al.[1] report PMMoV RNA concentrations measured in feces in units of gc/mg. We (1) filtered for concentrations above the detection limit, (2) converted concentrations to units of gc/g, and (3) $log_{10}$-transformed concentrations in gc/g. The mean and standard deviation of the $log_{10}$-transformed concentrations were 7.94 and 1.57, respectively. Zheng et al.[2] compiled rotavirus RNA concentrations measured in feces. We (1) filtered for papers with a quality score ≥ 2, (2) filtered for concentrations reported on a per mass basis, (3) converted all concentrations to units of gc/g (assuming genome copies ~ genome equivalents), (4) $log_{10}$-transformed concentrations in gc/g, and (5) weighted concentrations reported as central tendencies (median or average) by the sample number. The mean and standard deviation of the $log_{10}$-transformed concentrations were 7.01 and 2.16, respectively.

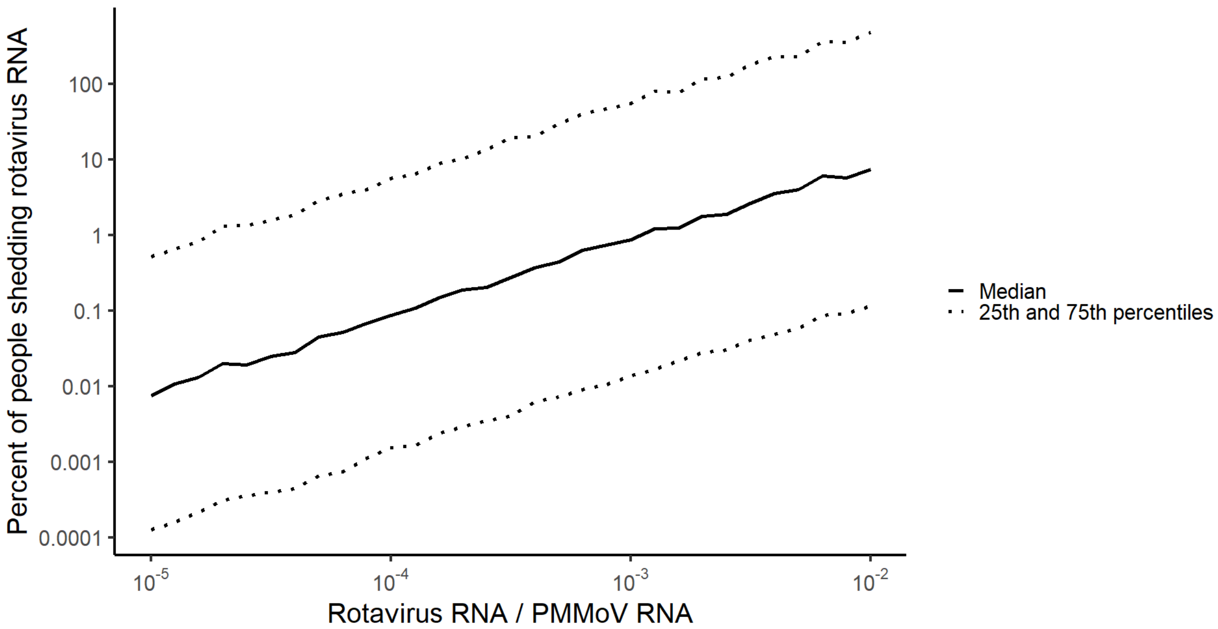


**Figure S11. Modeled percentage of people shedding rotavirus RNA as a function of PMMoV-normalized rotavirus RNA concentration in wastewater solids.** Abbreviation: PMMoV = pepper mild mottle virus.

#### Supplementary analysis: Comparing clinical surveillance metrics at the national scale

Weekly rotavirus clinical surveillance metrics from both NREVSS and Epic Cosmos are not normally distributed (Shapiro-Wilk test, $p < 0.0001$). Weekly rotavirus clinical surveillance metrics from NREVSS and Epic Cosmos are temporally correlated (Kendall's tau: 0.49, $p < 0.0001$) (**Figure S12**)

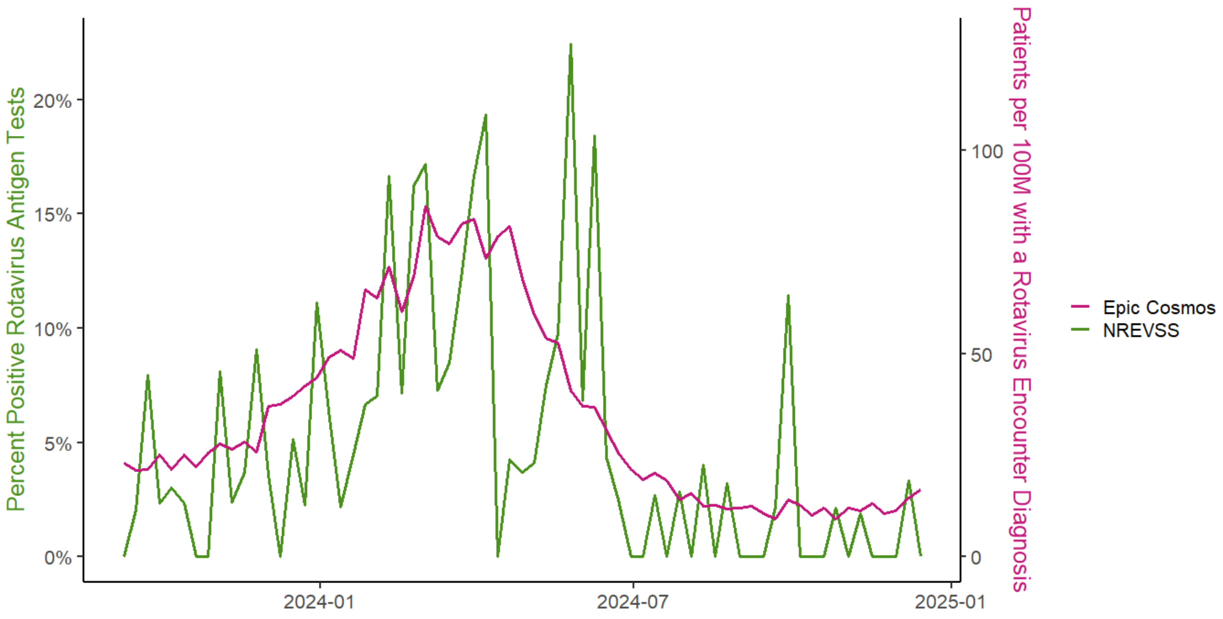


**Figure S12. Clinical rotavirus infection metrics for the United States from NREVSS versus Epic Cosmos.** Weekly rotavirus test positivity from NREVSS is shown on the left y-axis in green; weekly number of patients per 100 million with a rotaviral enteritis encounter diagnosis from Epic Cosmos is shown on the right y-axis in pink. Abbreviation: NREVSS: National Respiratory and Enteric Virus Surveillance System.

#### Supplementary analysis: Correlation with clinical metrics at the state scale

We also evaluated the temporal correlation between wastewater RNA concentrations of rotavirus and clinical metrics of rotavirus infections at the state scale. We assessed this correlation using only the proportion of patients with a rotaviral enteritis encounter diagnosis from Epic Cosmos as NREVSS test positivity data are only available at the national scale. We obtained diagnoses of rotavirus enteritis over the study period from Epic Cosmos as described in the main text—although we temporally grouped patient counts on a monthly basis and geographically grouped patient counts by state for this analysis. We assessed this correlation on a monthly rather than weekly basis because Epic Cosmos reports < 11 patients as “10 or fewer” and states had many patient counts of “10 or fewer” on a weekly basis. To match the reporting frequency of the clinical data obtained from Epic Cosmos, we temporally aggregated wastewater rotavirus RNA concentrations on a monthly basis for each WWTP by averaging wastewater rotavirus RNA concentrations across all samples collected each calendar month. We then spatially aggregated monthly average wastewater rotavirus RNA concentrations across WWTPs to the state scale using the population-weighted averaging approach. State-aggregated monthly wastewater rotavirus RNA concentrations were not always normally distributed (Shapiro-Wilk test, $p < 0.05$); therefore, we used Kendall’s tau correlation to test the null hypothesis that monthly rotavirus RNA wastewater concentrations are not temporally correlated with monthly rotavirus clinical metrics for each state. We only conducted the correlation for states with rotavirus wastewater monitoring and at least one month with > 10 patients with a rotaviral enteritis diagnoses (AL, CA, CO, CT, FL, GA, IL, IN, IA, KS, KY, LA, MI, MN, MS, NE, NJ, NY, NC, OH, PA, TN. TX, VA, WI). We imputed 0 patients for any months with “10 or fewer” patients prior to conducting the correlation tests. Results are shown in **Figure S13** and reported in **Table S1**.

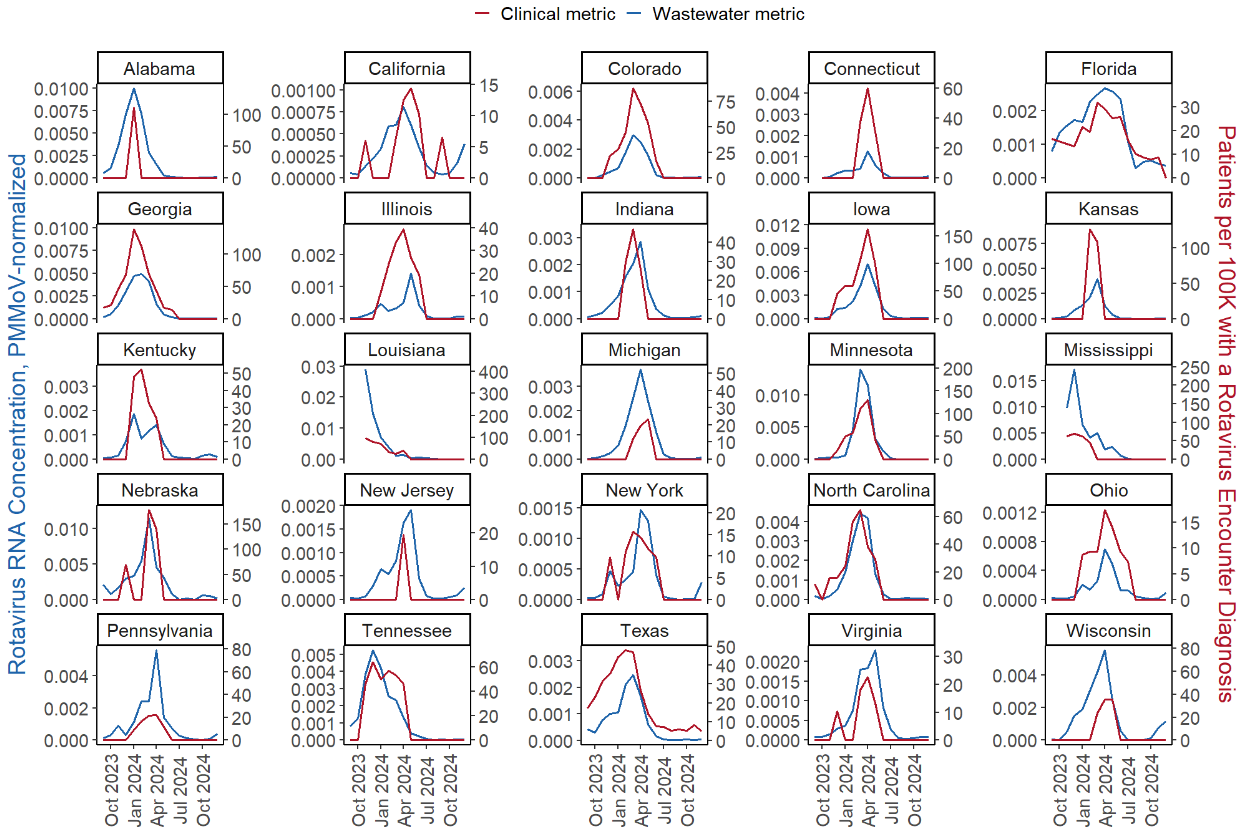


**Figure S13. Monthly correlation between wastewater rotavirus metric and clinical rotavirus metric for states.** Monthly PMMoV-normalized rotavirus RNA concentration in wastewater is shown on the left y-axis in blue); monthly clinical surveillance metric of rotavirus infections from Epic Cosmos is shown on the right y-axis in red. Abbreviation: PMMoV = pepper mild mottle virus.

**Table S1. Temporal correlation between monthly wastewater and clinical rotavirus metric for individual states**

| State | Kendall's Tau (p value) | Number of months |
|---|---|---|
| Alabama | 0.35 (0.10) | 16 |
| California | 0.34 (0.09) | 16 |
| Colorado | 0.80 (< 0.0001) | 16 |
| Connecticut | 0.58 (0.0082) | 15 |
| Florida | 0.64 (0.00052) | 16 |
| Georgia | 0.90 (< 0.0001) | 16 |
| Illinois | 0.64 (0.0015) | 16 |
| Indiana | 0.54 (0.011) | 16 |
| Iowa | 0.76 (0.00017) | 16 |
| Kansas | 0.46 (0.033) | 16 |
| Kentucky | 0.57 (0.0060) | 16 |
| Louisiana | 0.81 (0.00020) | 14 |
| Michigan | 0.54 (0.011) | 16 |
| Minnesota | 0.69 (0.00071) | 16 |
| Mississippi | 0.67 (0.0028) | 14 |
| Nebraska | 0.48 (0.023) | 16 |
| New Jersey | 0.31 (0.16) | 16 |
| New York | 0.68 (0.00084) | 16 |
| North Carolina | 0.79 (< 0.0001) | 16 |
| Ohio | 0.78 (0.00011) | 16 |
| Pennsylvania | 0.74 (0.00033) | 16 |
| Tennessee | 0.70 (0.00057) | 16 |
| Texas | 0.73 (< 0.0001) | 16 |
| Virginia | 0.55 (0.0086) | 16 |
| Wisconsin | 0.53 (0.013) | 16 |

### Supplementary analysis: Rotavirus encounter diagnoses by age group

We obtained the weekly proportion of patients with a rotaviral enteritis encounter diagnosis each week from Epic Cosmos as described in the main text, although we further disaggregated the data by the following age groups: 0-2 years, 2-5 years, 5-18 years, 18-65 years, and 65+ years (**Figure S14**). Some weekly counts were reported as "10 or fewer patients"; we imputed 0 patients in these instances.

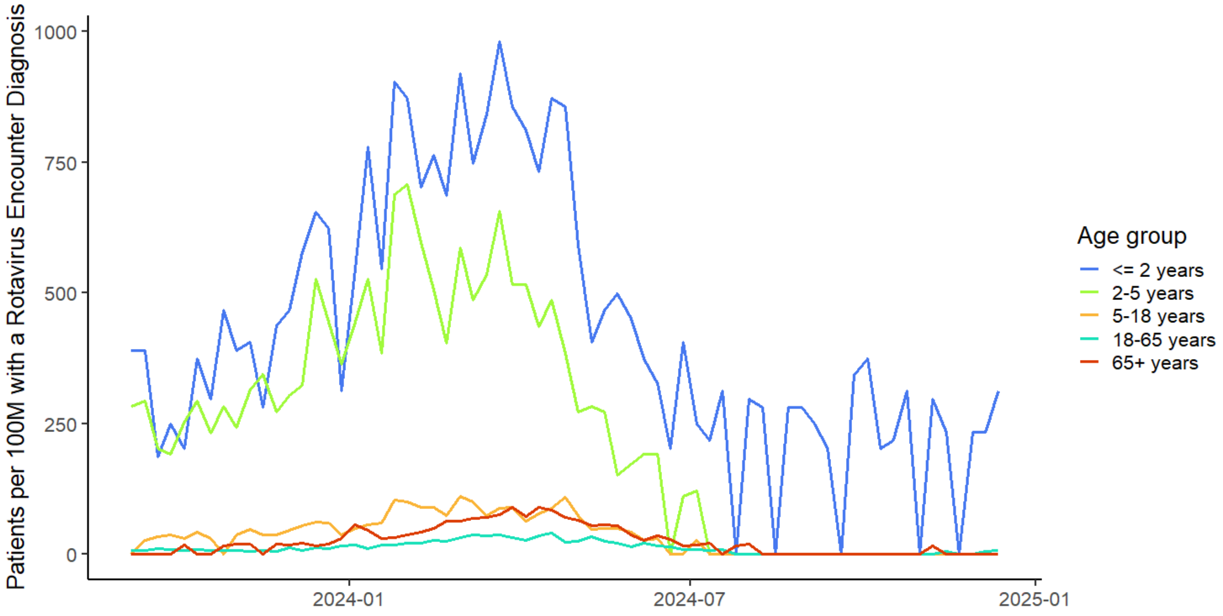


**Figure S14. Weekly number of patients per 100 million with a rotaviral enteritis encounter diagnosis from Epic Cosmos by age group.**

#### Wastewater monitoring: Further details

**Table S2. Wastewater sampling details of each wastewater treatment plant**

| State | County[1] | Location | WWTP name | Service population | Date of first/ last sample | Sample count | Median Rotavirus RNA concentration in gc/g (IQR) |
|---|---|---|---|---|---|---|---|
| Alabama | Jefferson | Bessemer, AL | Valley Creek Water Reclamation Facility | 225000 | 2023-09-08, 2024-12-11 | 198 | 109616.39 (10880.26, 827029.01) |
| Alabama | Jefferson | Cahaba River, Birmingham, AL | Cahaba River Water Reclamation Facility | 95000 | 2023-09-08, 2024-12-11 | 192 | 174061.78 (24895.9, 618139.77) |
| Alabama | Jefferson | Fultondale, AL | Five Mile Creek Water Reclamation Facility | 77000 | 2023-09-08, 2024-12-11 | 196 | 58784.2 (6834.53, 429906.53) |
| Alabama | Jefferson | Pinson, AL | Turkey Creek Water Reclamation Facility | 30000 | 2023-09-08, 2024-12-11 | 192 | 8396.72 (0, 115925.11) |
| Alabama | Jefferson | Village Creek, Birmingham, AL | Village Creek Water Reclamation Facility | 200000 | 2023-09-07, 2024-12-11 | 175 | 70868.57 (15775.01, 479416.69) |
| Alaska | Anchorage | Anchorage, AK | John M. Asplund Water Pollution Control Facility | 220000 | 2023-09-07, 2024-12-12 | 193 | 163304.06 (46382.34, 965414.18) |
| Arkansas | Boone | Harrison, AR | City of Harrison Wastewater Treatment Plant | 15000 | 2023-09-08, 2024-12-11 | 197 | 44965.68 (0, 620446.97) |
| California | Alameda | Fremont, CA | [Fremont Basin] - Raymond A. Boege Alvarado WWTP | 229476 | 2023-09-05, 2024-12-10 | 120 | 17054.24 (4570.85, 66494.19) |
| California | Alameda | Newark, CA | [Newark Basin] - Raymond A. Boege Alvarado WWTP | 47229 | 2023-09-05, 2024-06-27 | 83 | 26989.5 (8560.15, 66287.26) |
| California | Alameda | Oakland, CA | East Bay Municipal Utility District | 740000 | 2023-09-12, 2024-06-27 | 126 | 125894.65 (24557.65, 233666.72) |
| California | Alameda | San Leandro, CA | City of San Leandro Water Pollution Control Plant | 50000 | 2023-09-06, 2024-06-27 | 127 | 36307.12 (6926.9, 119319.47) |
| California | Alameda | Union City, CA | [Union City Basin] - Raymond A. Boege Alvarado WWTP | 68150 | 2023-09-05, 2024-06-27 | 83 | 45481.7 (14869.32, 88726.89) |

| State | County[1] | Location | WWTP name | Service population | Date of first/ last sample | Sample count | Median Rotavirus RNA concentration in gc/g (IQR) |
|---|---|---|---|---|---|---|---|
| California | Contra Costa | Contra Costa County, CA | Central Contra Costa Sanitary District | 484800 | 2023-09-08, 2024-06-25 | 115 | 67935.07 (25535.6, 162033.8) |
| California | Contra Costa | West Contra Costa County, CA | West County Wastewater District | 100000 | 2023-09-11, 2024-06-27 | 108 | 29688.62 (8303.21, 87298.59) |
| California | Los Angeles | Lancaster, CA | Lancaster Water Reclamation Plant | 200000 | 2023-09-11, 2024-06-27 | 112 | 170404.56 (62896.03, 464126.73) |
| California | Los Angeles | Los Angeles County, CA | A.K. Warren Water Resource Facility | 3500000 | 2023-09-10, 2024-12-11 | 198 | 137300.09 (37377.53, 568230.65) |
| California | Los Angeles | Los Angeles, CA | Hyperion Water Reclamation Plant (HWRP) | 4000000 | 2023-09-10, 2024-12-11 | 196 | 153502.78 (41932.19, 395898) |
| California | Madera | Madera, CA | City of Madera, Wastewater Treatment Plant | 67944 | 2023-09-08, 2024-06-28 | 120 | 211455.23 (58503.85, 823325.51) |
| California | Marin | Las Gallinas, San Rafael, CA | Las Gallinas Valley Sanitary District | 30000 | 2023-09-11, 2024-06-26 | 126 | 59077.95 (17412.14, 198178.48) |
| California | Marin | Mill Valley, CA | Sewerage Agency of Southern Marin Wastewater Treatment Plant | 30000 | 2023-09-06, 2024-06-20 | 124 | 70245.87 (10872.38, 173136.11) |
| California | Marin | Novato, CA | Novato Sanitary District | 53000 | 2023-09-08, 2024-12-11 | 198 | 113235.98 (30996.91, 357551.08) |
| California | Marin | San Rafael, CA | Central Marin Sanitation Agency | 104250 | 2023-09-10, 2024-12-10 | 119 | 114720.59 (33770.05, 273395.8) |
| California | Marin | Sausalito, CA | Sausalito-Marin City Sanitary District | 18000 | 2023-09-12, 2024-06-27 | 115 | 34205.23 (7402.21, 155866.9) |
| California | Merced | Los Banos, CA | Los Banos Wastewater Treatment Plant | 42000 | 2023-09-08, 2024-06-28 | 118 | 102667.92 (18796.41, 331052.79) |
| California | Merced | Merced, CA | Merced Wastewater Treatment Plant | 91000 | 2023-09-08, 2024-12-11 | 198 | 26728.49 (7248.42, 94110.73) |

| State | County[1] | Location | WWTP name | Service population | Date of first/ last sample | Sample count | Median Rotavirus RNA concentration in gc/g (IQR) |
|---|---|---|---|---|---|---|---|
| California | Mono | Mammoth, CA | Mammoth Community Water District | 35000 | 2023-09-08, 2024-06-28 | 121 | 122084.5 (42239.38, 461837.08) |
| California | Monterey | Marina, CA | Monterey One Water - Regional Treatment Plant | 262000 | 2023-09-07, 2024-12-11 | 180 | 192311.49 (57821.5, 494518.68) |
| California | Napa | Napa, CA | Soscol Water Recycling Facility | 83300 | 2023-09-08, 2024-12-11 | 196 | 61763.14 (15710.86, 196386.07) |
| California | Orange | Coastal, Laguna Niguel, CA | Coastal Treatment Plant | 48000 | 2023-09-09, 2024-06-28 | 127 | 62694.09 (10571.23, 218220.72) |
| California | Orange | JB Latham, Laguna Niguel, CA | JB Latham Treatment Plant | 120000 | 2023-09-09, 2024-06-28 | 128 | 75026.29 (18023.71, 162050.76) |
| California | Orange | Regional, Laguna Niguel, CA | Regional Treatment Plant | 129000 | 2023-09-09, 2024-06-28 | 127 | 77174.59 (9917.24, 213962.63) |
| California | Riverside | Indio, CA | Valley Sanitary District | 91765 | 2023-09-08, 2024-12-11 | 198 | 50455.79 (7390.6, 130492.07) |
| California | Riverside | Riverside, CA | Riverside Water Quality Control Plant | 350000 | 2023-09-08, 2024-12-11 | 198 | 126437.79 (43739.43, 511371.66) |
| California | Sacramento | Sacramento, CA | Sacramento Regional Wastewater Treatment Plant | 1480000 | 2023-09-03, 2024-11-22 | 446 | 141311.14 (57904.99, 355385.32) |
| California | San Benito | Hollister, CA | City of Hollister Domestic Water Recycling Facility | 42000 | 2023-09-08, 2024-06-28 | 116 | 66317.54 (17538.2, 305008.15) |
| California | San Bernardino | Ontario, CA | Regional Water Recycling Plant No.1 (RP-1) | 890000 | 2023-09-11, 2024-12-12 | 193 | 60735.55 (19046.92, 181797.51) |
| California | San Diego | San Diego, CA | E.W. Blom Point Loma Wastewater Treatment Plant | 2200000 | 2023-09-10, 2024-12-11 | 188 | 97659.36 (44827.5, 319529) |
| California | San Francisco | Oceanside, San Francisco, CA | Oceanside Water Pollution Control Plant | 250000 | 2023-09-03, 2024-11-22 | 425 | 34757.93 (10975.28, 100455.45) |

| State | County[1] | Location | WWTP name | Service population | Date of first/ last sample | Sample count | Median Rotavirus RNA concentration in gc/g (IQR) |
|---|---|---|---|---|---|---|---|
| California | San Francisco | Southeast San Francisco, CA | Southeast San Francisco | 750000 | 2023-09-03, 2024-11-21 | 435 | 38913.36 (17905.94, 87568.2) |
| California | San Luis Obispo | Paso Robles, CA | City of Paso Robles Wastewater Treatment Plant | 31037 | 2023-09-11, 2024-12-12 | 194 | 114185.98 (28372.76, 424341.66) |
| California | San Mateo | Half Moon Bay, CA | Sewer Authority Mid-Coastside | 28000 | 2023-09-08, 2024-06-28 | 123 | 34094.47 (0, 180654.81) |
| California | San Mateo | Redwood City, CA | Silicon Valley Clean Water | 199000 | 2023-09-03, 2024-11-22 | 373 | 66665.48 (22954.23, 180813.84) |
| California | San Mateo | San Mateo, CA | City of San Mateo & Estero M.I.D. Water Quality Control Plant | 150000 | 2023-09-08, 2024-06-28 | 126 | 56344.72 (17854.24, 100446.6) |
| California | Santa Barbara | Lompoc, CA | Lompoc Regional Wastewater Reclamation Plant | 69290 | 2023-09-08, 2024-12-11 | 197 | 37629.81 (5828.36, 163961.48) |
| California | Santa Clara | CODIGA, Stanford, CA | CODIGA | 10000 | 2023-09-04, 2024-05-31 | 233 | 0 (0, 11552.87) |
| California | Santa Clara | Gilroy, CA | South County Regional Wastewater Authority | 110338 | 2023-09-03, 2024-11-22 | 447 | 19992.38 (4006.4, 99770.69) |
| California | Santa Clara | Palo Alto, CA | Palo Alto Regional Water Quality Control Plant | 236000 | 2023-09-03, 2024-11-21 | 445 | 60285.32 (23959.64, 163338.7) |
| California | Santa Clara | San Jose, CA | San Jose-Santa Clara Regional Wastewater Facility | 1500000 | 2023-09-03, 2024-11-21 | 445 | 191882.76 (73041.45, 556350.75) |
| California | Santa Clara | Sunnyvale, CA | City of Sunnyvale Water Pollution Control Plant | 153000 | 2023-09-03, 2024-11-22 | 436 | 52305.03 (20749.38, 131229.09) |
| California | Santa Cruz | Santa Cruz County, CA | City of Santa Cruz WTF - County Influent | 160000 | 2023-09-07, 2024-06-30 | 125 | 63067.4 (18331.8, 185242.95) |
| California | Santa Cruz | Santa Cruz, CA | City of Santa Cruz WTF - City Influent | 160000 | 2023-09-10, 2024-12-10 | 195 | 16625.1 (0, 95415.97) |

| State | County[1] | Location | WWTP name | Service population | Date of first/ last sample | Sample count | Median Rotavirus RNA concentration in gc/g (IQR) |
|---|---|---|---|---|---|---|---|
| California | Solano | Fairfield, CA | Fairfield-Suisun Sewer District | 155000 | 2023-09-08, 2024-06-28 | 127 | 92047.6 (16516.61, 264041.33) |
| California | Solano | Vallejo, CA | Vallejo Flood and Wastewater District Wastewater Treatment Plant | 121000 | 2023-09-12, 2024-12-12 | 199 | 92425.28 (13108.59, 305417.82) |
| California | Sonoma | Santa Rosa, CA | City of Santa Rosa, Laguna Treatment Plant | 230000 | 2023-09-08, 2024-12-11 | 190 | 93550.86 (20565.97, 288268.92) |
| California | Sonoma | Windsor, CA | Windsor Wastewater Treatment, Reclamation, and Disposal Facility | 28000 | 2023-09-08, 2024-06-28 | 116 | 34684.9 (5690.99, 146141.45) |
| California | Stanislaus | Modesto, CA | Modesto's Sutter Primary Treatment Facility | 230000 | 2023-09-11, 2024-06-27 | 123 | 290840.39 (138908, 693845.7) |
| California | Stanislaus | Turlock, CA | Turlock Regional Water Quality Control Facility | 86000 | 2023-09-08, 2024-12-11 | 198 | 13415.86 (1748.99, 56640.91) |
| California | Yolo | Davis, CA | City of Davis Wastewater Treatment Plant | 68000 | 2023-09-08, 2024-12-11 | 189 | 17339.63 (6067.35, 54360.42) |
| California | Yolo | Esparto, CA | Esparto Wastewater Treatment Facility | 4006 | 2023-09-06, 2024-06-28 | 127 | 4238.31 (0, 26199.62) |
| California | Yolo | Winters, CA | Winters - East Street Pump Station | 7286 | 2023-09-04, 2024-06-27 | 113 | 0 (0, 13863.45) |
| California | Yolo | Woodland, CA | Woodland Water Pollution Control Facility | 59000 | 2023-09-08, 2024-12-09 | 160 | 21714.6 (6071.18, 78018.77) |
| Colorado | Douglas | North, Parker, CO | Parker Water and Sanitation District North Water Reclamation Facility | 35000 | 2023-09-11, 2024-12-12 | 179 | 42135.6 (4816.63, 268251.56) |
| Colorado | Douglas | South, Parker, CO | Parker Water and Sanitation District South Water Reclamation Facility | 25000 | 2023-09-11, 2024-12-12 | 176 | 21608.54 (0, 263746.43) |

| State | County[1] | Location | WWTP name | Service population | Date of first/ last sample | Sample count | Median Rotavirus RNA concentration in gc/g (IQR) |
|---|---|---|---|---|---|---|---|
| Connecticut | Western Connecticut | Stamford, CT | City of Stamford, Water Pollution Control Authority | 140000 | 2023-10-27, 2024-12-11 | 166 | 45235.95 (15046.99, 127884.3) |
| Delaware | Sussex | Seaford, DE | Seaford Wastewater Treatment Facility | 13172 | 2023-09-08, 2024-12-11 | 198 | 22417.73 (6265.83, 69190.72) |
| District of Columbia | Montgomery | Blue Plains, Washington, DC | Blue Plains Advanced Wastewater Treatment Plant | 2000000 | 2023-11-12, 2024-06-03 | 82 | 179616.39 (65490.51, 431000.35) |
| Florida | Leon | Tallahassee, FL | TPSmith Water Reclamation Facility | 212065 | 2023-09-07, 2024-12-10 | 193 | 118363.9 (18589.57, 400209.21) |
| Florida | Miami-Dade | Key Biscayne, FL | MDWASD Central District WWTP | 829725 | 2023-09-11, 2024-12-10 | 175 | 202573.42 (89385.83, 410602.86) |
| Florida | Miami-Dade | North Miami, FL | MDWASD North District WWTF | 776150 | 2023-09-08, 2024-12-11 | 187 | 146309.39 (67253.95, 280319.77) |
| Florida | Orange | Eastern, Orange County, FL | Eastern Water Reclamation Facility | 195299 | 2023-09-07, 2024-12-10 | 158 | 54092.45 (21249.65, 121168.29) |
| Florida | Orange | Northwest, Orange County, FL | Northwest Water Reclamation Facility | 66690 | 2023-09-07, 2024-12-10 | 159 | 37226.42 (10226.74, 159934.77) |
| Florida | Orange | South, Orange County, FL | South Water Reclamation Facility | 183009 | 2023-09-07, 2024-12-10 | 159 | 63947.83 (18494.3, 166043.94) |
| Florida | Orange | Southwest, Orange County, FL | Hamlin Water Reclamation Facility | 50000 | 2023-09-07, 2024-12-10 | 156 | 42958.86 (7447.27, 202934.56) |
| Florida | Palm Beach | Jupiter, FL | Loxahatchee River Environmental Control District | 90000 | 2023-09-08, 2024-12-11 | 197 | 66977.06 (14164.7, 265559.69) |
| Florida | Pinellas | Northeast, Saint Petersburg, FL | Northeast Water Reclamation Facility | 89847 | 2023-09-08, 2024-12-11 | 183 | 113514.12 (16362.61, 462383.35) |
| Florida | Pinellas | Northwest, Saint Petersburg, FL | Northwest Water Reclamation Facility | 94218 | 2023-09-08, 2024-12-11 | 186 | 79440.49 (10788.23, 292155.52) |

| State | County[1] | Location | WWTP name | Service population | Date of first/ last sample | Sample count | Median Rotavirus RNA concentration in gc/g (IQR) |
|---|---|---|---|---|---|---|---|
| Florida | Pinellas | Southwest, Saint Petersburg, FL | Southwest Water Reclamation Facility | 47790 | 2023-09-08, 2024-12-11 | 181 | 103874.24 (6485.13, 410578.98) |
| Florida | Seminole | Altamonte Springs, FL | Altamonte Springs Regional Water Reclamation Facility | 95000 | 2023-09-08, 2024-06-28 | 120 | 314252.62 (136939.2, 788970.43) |
| Georgia | DeKalb | RM Clayton, Atlanta, GA | RM Clayton Water Reclamation Center | 294660 | 2023-09-05, 2024-12-08 | 173 | 94739.19 (22608.88, 416597.82) |
| Georgia | Fulton | Big Creek, Roswell, GA | Big Creek Water Reclamation Facility | 189593 | 2023-09-06, 2024-12-11 | 198 | 60112.8 (0, 390227.81) |
| Georgia | Fulton | College Park, GA | Camp Creek Water Reclamation Facility | 73821 | 2023-09-07, 2024-12-10 | 177 | 48866.49 (3808.65, 505285.88) |
| Georgia | Fulton | Johns Creek, Roswell, GA | Johns Creek Environmental Campus | 84486 | 2023-09-11, 2024-12-11 | 196 | 53801.35 (11583.98, 310485.8) |
| Georgia | Fulton | Little River, Roswell, GA | Little River Water Reclamation Facility | 12818 | 2023-09-11, 2024-12-11 | 197 | 36013.2 (0, 437482.32) |
| Georgia | Fulton | South River, Atlanta, GA | South River Water Reclamation Center | 105160 | 2023-09-05, 2024-12-08 | 173 | 68046.46 (18560.53, 357024.78) |
| Georgia | Fulton | Utoy Creek, Atlanta, GA | Utoy Creek Water Reclamation Center | 70887 | 2023-09-05, 2024-12-08 | 173 | 37229.44 (0, 272608.91) |
| Georgia | Muscogee | Columbus, GA | South Columbus Water Resources Facility | 278000 | 2023-09-08, 2024-12-11 | 121 | 109986.68 (7965.29, 506677.88) |
| Hawaii | Honolulu | Honouliuli, Honolulu, HI | Honouliuli Wastewater Treatment Plant | 300000 | 2023-09-06, 2024-12-11 | 198 | 58743.17 (19982.19, 194668.96) |
| Hawaii | Honolulu | Kailua, Honolulu, HI | Kailua Regional Wastewater Treatment Plant | 90000 | 2023-09-07, 2024-06-26 | 127 | 102673.49 (33489.17, 279956.33) |
| Hawaii | Honolulu | Sand Island, Honolulu, HI | Sand Island Wastewater Treatment Plant | 390000 | 2023-09-07, 2024-12-11 | 199 | 61136.19 (18481.28, 192291.13) |
| Hawaii | Honolulu | Wahiawa, | Wahiawa Wastewater | 18000 | 2023-09-07, | 125 | 20506.37 (4363.5, |

| State | County[1] | Location | WWTP name | Service population | Date of first/ last sample | Sample count | Median Rotavirus RNA concentration in gc/g (IQR) |
|---|---|---|---|---|---|---|---|
| | | Honolulu, HI | Treatment Plant | | 2024-06-26 | | 115223.91) |
| Hawaii | Honolulu | Waianae, Honolulu, HI | Waianae Wastewater Treatment Plant | 44000 | 2023-09-07, 2024-06-26 | 127 | 177923.36 (54269.81, 549009.93) |
| Idaho | Ada | Lander Street, Boise, ID | Lander Street Water Renewal Facility | 108556 | 2023-09-08, 2024-12-11 | 196 | 37011.43 (13225.86, 132480.23) |
| Idaho | Ada | West Boise, ID | West Boise Water Renewal Facility | 186901 | 2023-09-08, 2024-12-11 | 195 | 47601.72 (11944.48, 202447.71) |
| Idaho | Kootenai | Coeur d'Alene, ID | City of Coeur d'Alene Water Resource Recovery Facility | 50540 | 2023-09-11, 2024-12-11 | 173 | 22761.87 (7668.24, 81502.27) |
| Illinois | DuPage | Glen Ellyn, IL | Glenbard Wastewater Authority | 86000 | 2023-09-11, 2024-12-10 | 184 | 24487.89 (3386.83, 94318.78) |
| Illinois | DuPage | Wheaton, IL | Wheaton Sanitary District | 63000 | 2023-09-08, 2024-12-11 | 191 | 54565.59 (18944.31, 172163.52) |
| Indiana | Clark | Downtown, Jeffersonville, IN | Jeffersonville Downtown WWTP | 25000 | 2023-09-08, 2024-12-11 | 197 | 33042.99 (2539.05, 276419.92) |
| Indiana | Clark | North, Jeffersonville, IN | North Water Reclamation Facility | 25000 | 2023-09-08, 2024-12-11 | 197 | 42697.51 (7421.22, 290132.59) |
| Indiana | Hamilton | Carmel, IN | City of Carmel WWTP | 86000 | 2023-09-08, 2024-12-09 | 197 | 51146.77 (14650.34, 217277.84) |
| Indiana | Monroe | Bloomington, IN | Dillman Road WWTP | 56090 | 2023-09-11, 2024-12-12 | 222 | 36997.46 (2417.3, 214345.4) |
| Indiana | St. Joseph | South Bend, IN | City of South Bend Wastewater Treatment Plant | 130000 | 2023-09-07, 2024-12-10 | 190 | 61554.73 (14435.56, 192493.93) |
| Iowa | Clinton | Clinton, IA | City of Clinton | 29300 | 2023-09-08, 2024-12-11 | 198 | 25130.78 (6809.82, 233515.47) |
| Iowa | Johnson | Coralville, IA | Coralville Wastewater Treatment Facility | 23000 | 2023-09-08, 2024-12-11 | 195 | 44086.1 (4992.75, 544882.44) |

| State | County[1] | Location | WWTP name | Service population | Date of first/ last sample | Sample count | Median Rotavirus RNA concentration in gc/g (IQR) |
|---|---|---|---|---|---|---|---|
| Iowa | Marshall | Marshalltown, IA | City of Marshalltown Water Pollution Control Plant | 27400 | 2023-09-07, 2024-12-10 | 191 | 149067.47 (39211.78, 585751.49) |
| Iowa | Muscatine | Muscatine, IA | Muscatine STP | 24400 | 2023-09-11, 2024-12-11 | 183 | 8861.82 (0, 82029.93) |
| Iowa | Wapello | Ottumwa, IA | Ottumwa WPCF | 25529 | 2023-09-08, 2024-12-11 | 198 | 96982.76 (34644.2, 364719.03) |
| Kansas | Douglas | Lawrence, KS | Lawrence Kansas River Wastewater Treatment Facility | 80000 | 2023-09-08, 2024-12-11 | 193 | 61901.54 (4081.77, 469259.13) |
| Kansas | Saline | Salina, KS | Salina Wastewater Treatment Plant | 47000 | 2023-09-08, 2024-12-11 | 198 | 55542.72 (8271.2, 447643.92) |
| Kansas | Wyandotte | Kaw Point, Kansas City, KS | Municipal Wastewater Treatment Plant No. 1 (Kaw Point) | 90000 | 2023-09-08, 2024-12-05 | 189 | 18117.24 (2921.12, 220761.55) |
| Kansas | Wyandotte | P20, Kansas City, KS | Kansas City Treatment Plant #20 | 35000 | 2023-09-08, 2024-12-05 | 170 | 8494.45 (0, 175993.87) |
| Kansas | Wyandotte | Wolcott, Kansas City, KS | Wolcott Wastewater Treatment Facility | 15000 | 2023-09-08, 2024-12-05 | 179 | 12344.91 (0, 258956.44) |
| Kentucky | Jefferson | Louisville, KY | Morris Forman Water Quality Treatment Center | 423913 | 2023-09-14, 2024-12-12 | 166 | 22592.18 (7218.51, 92919.27) |
| Louisiana | Orleans | East Bank, New Orleans, LA | SWBNO East Bank Wastewater Treatment Plant | 333406 | 2023-11-27, 2024-12-11 | 164 | 73793.18 (11298.74, 379041.58) |
| Louisiana | Orleans | West Bank, New Orleans, LA | SWBNO West Bank Wastewater Treatment Plant | 50591 | 2023-11-27, 2024-12-11 | 155 | 52139.13 (3348.69, 391030.89) |
| Maine | Androscoggin | Lewiston, ME | Lewiston Auburn Clean Water Authority | 60000 | 2023-09-14, 2024-12-10 | 162 | 73595.54 (20801.19, 247438.47) |
| Maine | Cumberland | Brunswick, ME | Brunswick Sewer District | 10000 | 2023-09-12, 2024-06-27 | 114 | 67122.1 (16301.89, 194339.27) |

| State | County[1] | Location | WWTP name | Service population | Date of first/ last sample | Sample count | Median Rotavirus RNA concentration in gc/g (IQR) |
|---|---|---|---|---|---|---|---|
| Maine | Cumberland | Portland, ME | Portland Water District (East End Wastewater Treatment Facility) | 65000 | 2023-09-20, 2024-12-11 | 141 | 42823.36 (9924.96, 171758.95) |
| Maine | Penobscot | Bangor, ME | City of Bangor Wastewater Treatment Plant | 40000 | 2023-09-11, 2024-12-12 | 187 | 14565.82 (0, 154833.35) |
| Maine | York | York, ME | York Sewer District | 10000 | 2023-09-11, 2024-06-06 | 103 | 14674.88 (0, 159852.49) |
| Maryland | St. Mary's | Hollywood, MD | Marlay Taylor Water Reclamation Facility | 55000 | 2023-09-08, 2024-12-11 | 173 | 51138.75 (9675.77, 319810.13) |
| Maryland | Washington | Hagerstown, MD | Hagerstown Wastewater Treatment Plant | 90000 | 2023-09-08, 2024-12-11 | 196 | 6823.29 (0, 50730.95) |
| Massachusetts | Middlesex | Boston, MA | Deer Island Treatment Plant | 2400000 | 2023-09-08, 2024-12-11 | 195 | 17421.09 (6772.91, 80138.26) |
| Massachusetts | Worcester | Millbury, MA | Upper Blackstone Clean Water | 250000 | 2023-09-07, 2024-12-10 | 181 | 34761.18 (7179.48, 186172.06) |
| Michigan | Grand Traverse | Traverse City, MI | Traverse City Regional Waste Water Treatment Plant | 30623 | 2023-09-07, 2024-12-12 | 192 | 33953.88 (5561.24, 172195.47) |
| Michigan | Isabella | Mt. Pleasant, MI | Mt. Pleasant WRRF | 21690 | 2023-09-08, 2024-12-11 | 189 | 23100.29 (3729.24, 255671.31) |
| Michigan | Jackson | Jackson, MI | Jackson Wastewater Treatment Plant | 90000 | 2023-09-07, 2024-12-10 | 197 | 74088.83 (3864.06, 372908.68) |
| Michigan | Macomb | Warren, MI | City of Warren Wastewater Treatment Plant | 140000 | 2023-09-07, 2024-12-12 | 166 | 14421.86 (0, 78643.4) |
| Michigan | Ottawa | Jenison, MI | Grandville Clean Water Plant | 75000 | 2023-09-08, 2024-12-11 | 198 | 35055.08 (4217.61, 226034.56) |
| Michigan | Washtenaw | Ann Arbor, MI | City of Ann Arbor Wastewater Treatment Plant | 125000 | 2023-09-08, 2024-12-11 | 197 | 43341.83 (5394.06, 262194.38) |

| State | County[1] | Location | WWTP name | Service population | Date of first/ last sample | Sample count | Median Rotavirus RNA concentration in gc/g (IQR) |
|---|---|---|---|---|---|---|---|
| Minnesota | Blue Earth | Mankato, MN | City of Mankato Water Resource Recovery Facility (WRRF) | 70000 | 2023-09-08, 2024-12-12 | 197 | 65630.84 (7514.64, 700458.02) |
| Minnesota | Goodhue | Red Wing, MN | Red Wing Wastewater Treatment Facility | 16000 | 2023-09-08, 2024-12-11 | 197 | 24236.91 (4350.97, 249443.67) |
| Minnesota | Olmsted | Rochester, MN | City Of Rochester MN Water Reclamation Plant | 120000 | 2023-09-08, 2024-12-11 | 192 | 54396.33 (8077.58, 286397.72) |
| Minnesota | Stearns | St. Cloud, MN | St. Cloud Nutrient, Energy and Water Recovery Facility | 120000 | 2023-09-08, 2024-12-11 | 197 | 69300.19 (7512.92, 726853.62) |
| Mississippi | Jackson | Gautier, MS | 2C-Gautier POTW | 19008 | 2023-11-13, 2024-12-10 | 150 | 68486.62 (0, 391964.24) |
| Mississippi | Jackson | Pascagoula Moss Point, MS | 7 C- Pascagoula Moss Point POTW | 34333 | 2023-11-12, 2024-11-26 | 146 | 31007.55 (0, 584376.28) |
| Nebraska | Lancaster | Northeast, Lincoln, NE | Northeast Water Resource Recovery Facility | 60000 | 2023-09-08, 2024-12-11 | 197 | 130352.98 (7053.31, 854510.97) |
| Nebraska | Lancaster | Theresa Street, Lincoln, NE | Theresa Street Water Resource Recovery Facility | 240000 | 2023-09-08, 2024-12-11 | 197 | 224139.72 (32009.16, 822075.91) |
| Nevada | Clark | Las Vegas, NV | Clark County Water Reclamation District (CCWRD) Flamingo Water Resource Center (FWRC) | 990000 | 2023-09-08, 2024-12-11 | 212 | 88641.91 (24409.07, 275844.35) |
| New Hampshire | Merrimack | Hall Street, Concord, NH | Hall Street Wastewater Treatment Plant | 45000 | 2023-09-08, 2024-06-26 | 117 | 179785.72 (30792.38, 590616.75) |
| New Hampshire | Merrimack | Penacook, Concord, NH | Penacook Wastewater Treatment Facility | 4000 | 2023-09-08, 2024-06-26 | 115 | 41156.07 (3304.73, 312651.21) |
| New Hampshire | Strafford | Dover, NH | City of Dover Wastewater Treatment Facility | 30000 | 2023-09-11, 2024-12-12 | 192 | 7884.17 (0, 42219.34) |
| New Jersey | Cumberland | Bridgeton, NJ | Cumberland County Utilities | 50000 | 2023-09-08, | 179 | 13406.82 (3721.85, 68575.3) |

| State | County[1] | Location | WWTP name | Service population | Date of first/ last sample | Sample count | Median Rotavirus RNA concentration in gc/g (IQR) |
|---|---|---|---|---|---|---|---|
| | | | Authority | | 2024-12-11 | | |
| New Jersey | Essex | Newark, NJ | Passaic Valley Sewerage Commission | 1500000 | 2023-09-11, 2024-12-11 | 156 | 23363.12 (8528.58, 103847.99) |
| New Jersey | Monmouth | Belmar, NJ | South Monmouth Regional Sewerage Authority | 52672 | 2023-09-08, 2024-12-12 | 197 | 13995.4 (0, 71061.5) |
| New Jersey | Monmouth | Oakhurst, NJ | Township of Ocean Sewerage Authority | 50000 | 2023-09-07, 2024-12-11 | 162 | 101660.19 (5029.58, 569298.53) |
| New Jersey | Monmouth | Union Beach, NJ | Bayshore Regional Sewerage Authority | 100000 | 2023-09-08, 2024-12-11 | 198 | 27334.75 (5167.72, 139539.36) |
| New Jersey | Somerset | Bridgewater, NJ | The Somerset Raritan Valley Sewerage Authority | 130000 | 2023-09-08, 2024-12-10 | 185 | 45406.28 (8447.37, 332321.32) |
| New York | Oswego | Oswego, NY | City of Oswego Wastewater Treatment Plant | 30000 | 2023-09-08, 2024-12-11 | 193 | 3354.39 (0, 23830.68) |
| New York | Tompkins | Ithaca, NY | Ithaca Area Wastewater Treatment Facility | 90000 | 2023-09-08, 2024-12-11 | 139 | 12254.55 (0, 94008.39) |
| North Carolina | Forsyth | Winston-Salem, NC | Archie Elledge WWTP | 92000 | 2023-09-08, 2024-12-11 | 182 | 42711.94 (6112.22, 230598.08) |
| North Carolina | Lenoir | Kinston, NC | Johnnie Mosley Regional Water Reclamation Facility | 25000 | 2023-09-11, 2024-12-11 | 184 | 28597.61 (5793.52, 223052.62) |
| North Carolina | Wilson | Wilson, NC | City of Wilson - Hominy Creek Water Reclamation Facility | 50000 | 2023-09-18, 2024-12-11 | 182 | 13311.91 (0, 295221.06) |
| Ohio | Mahoning | Youngstown, OH | City of Youngstown Wastewater Treatment Plant | 174000 | 2023-09-08, 2024-12-11 | 196 | 4644.96 (0, 27497.41) |
| Ohio | Summit | Akron, OH | Akron Water Reclamation Facility | 365000 | 2023-09-08, 2024-12-11 | 190 | 12238.08 (3745.55, 46014.11) |
| Pennsylvania | Centre | University Park, PA | University Park Water Reclamation Plant | 16000 | 2023-12-17, 2024-06-27 | 84 | 15001.65 (0, 56716.74) |

| State | County[1] | Location | WWTP name | Service population | Date of first/ last sample | Sample count | Median Rotavirus RNA concentration in gc/g (IQR) |
|---|---|---|---|---|---|---|---|
| Pennsylvania | Dauphin | Harrisburg, PA | Capital Region Water AWTF | 125000 | 2023-09-07, 2024-12-11 | 199 | 49767.51 (6972.62, 329857.91) |
| Pennsylvania | Delaware | Chester, PA | DELCORA Western Regional Treatment Plant | 220000 | 2023-09-07, 2024-12-10 | 174 | 60814.21 (15616.49, 195244.26) |
| South Dakota | Yankton | Yankton, SD | City of Yankton Wastewater Treatment Facility | 20000 | 2023-09-07, 2024-12-10 | 198 | 28213.17 (3758.48, 460476.23) |
| Tennessee | Hamilton | Chattanooga, TN | Moccasin Bend WWTP | 400000 | 2023-09-08, 2024-12-09 | 185 | 50880.79 (7666.38, 269669.36) |
| Tennessee | Shelby | Memphis, TN | M.C. Stiles Wastewater Treatment Facility | 300000 | 2023-09-07, 2024-12-10 | 187 | 23129.96 (2491.12, 125669.87) |
| Texas | Clay | Wichita Falls, TX | Wichita Falls Resource Recovery Facility | 90000 | 2023-09-08, 2024-12-11 | 188 | 19734.27 (3662.99, 118766.39) |
| Texas | Cooke | Gainesville, TX | City of Gainesville Wastewater Treatment Plant | 17300 | 2023-09-08, 2024-12-11 | 188 | 5789.12 (0, 56290.52) |
| Texas | Dallas | Dallas Central, Dallas, TX | DCWT Dallas | 270000 | 2023-09-08, 2024-12-10 | 173 | 126051 (21925.84, 436255.57) |
| Texas | Dallas | Garland, TX | City of Garland Rowlett Creek WWTP | 200000 | 2023-09-08, 2024-12-11 | 188 | 77215.91 (14468.42, 457975.68) |
| Texas | Dallas | Southside, Dallas, TX | Southside Wastewater Treatment Plant (City of Dallas) | 421700 | 2023-09-11, 2024-12-10 | 125 | 317067.68 (69346.06, 1330688.08) |
| Texas | Dallas | Sunnyvale, TX | Duck Creek Wastewater Treatment Plant | 186000 | 2023-09-08, 2024-12-10 | 148 | 58293.62 (9440.43, 279968.26) |
| Texas | Dallas | White Rock Central, Dallas, TX | DCWT White Rock | 630000 | 2023-09-08, 2024-12-10 | 176 | 218150.48 (28727.53, 793097.6) |
| Texas | Montgomery | Woodlands SJRA WWTF No. 1, TX | SJRA WWTF No.1 | 65000 | 2023-09-08, 2024-12-11 | 197 | 78292.06 (6182.06, 374958.54) |

| State | County[1] | Location | WWTP name | Service population | Date of first/ last sample | Sample count | Median Rotavirus RNA concentration in gc/g (IQR) |
|---|---|---|---|---|---|---|---|
| Texas | Montgomery | Woodlands SJRA WWTF No. 2, TX | SJRA WWTF No.2 | 70000 | 2023-09-08, 2024-12-11 | 197 | 78763.81 (11500.86, 310456.28) |
| Texas | Montgomery | Woodlands SJRA WWTF No. 3, TX | SJRA WWTF No.3 | 15000 | 2023-09-08, 2024-06-28 | 126 | 224177.59 (50666.66, 871799.69) |
| Texas | Potter | River Road, Amarillo, TX | River Road WWTP | 140000 | 2023-09-10, 2024-12-10 | 202 | 175698.27 (15982.42, 737955.03) |
| Texas | Randall | Hollywood Road, Amarillo, TX | Hollywood Road WWTP | 60000 | 2023-09-11, 2024-12-10 | 197 | 82299.83 (7718.15, 510108.86) |
| Texas | Webb | South, Laredo, TX | South Laredo WWTP | 120000 | 2023-09-08, 2024-12-11 | 181 | 182401.22 (5226.93, 1360291.32) |
| Texas | Webb | Zacate Creek, Laredo, TX | Zacate Creek WWTP | 140000 | 2023-09-08, 2024-12-11 | 181 | 182972.82 (12780.96, 723322.6) |
| Utah | Salt Lake | Central Salt Lake Valley, UT | Central Valley Water Reclamation Facility | 600000 | 2023-09-08, 2024-12-11 | 195 | 159016.72 (54820.88, 380930.02) |
| Utah | Utah | Provo, UT | Provo City Water Reclamation Facility | 115000 | 2023-09-11, 2024-12-11 | 128 | 169600.35 (40592.89, 410510.34) |
| Vermont | Chittenden | Essex Junction, VT | City of Essex Junction Wastewater Treatment Facility | 30000 | 2023-09-08, 2024-12-11 | 194 | 6742.71 (2014.13, 20213.36) |
| Vermont | Chittenden | South Burlington, VT | South Burlington-Airport Parkway WWTF | 16000 | 2023-09-12, 2024-12-12 | 119 | 9980.9 (0, 55359) |
| Vermont | Washington | Montpelier, VT | Montpelier Water Resource Recovery Facility | 10100 | 2023-09-11, 2024-12-12 | 189 | 6545.78 (0, 34992.92) |
| Virginia | Carroll | Hillsville, VA | Town of Hillsville Wastewater Treatment Plant | 3000 | 2023-09-08, 2024-06-28 | 123 | 36412.34 (13413.28, 92543.11) |
| Virginia | Stafford | Aquia, Stafford, VA | Aquia Wastewater Treatment Facility | 100000 | 2023-09-08, 2024-12-11 | 187 | 62204.43 (16944.56, 309252.88) |
| Virginia | Stafford | Little Falls Run, | Little Falls Run Wastewater | 50000 | 2023-09-08, | 187 | 68651.12 (15475.19, |

| State | County[1] | Location | WWTP name | Service population | Date of first/ last sample | Sample count | Median Rotavirus RNA concentration in gc/g (IQR) |
|---|---|---|---|---|---|---|---|
| | | Stafford, VA | Treatment Facility | | 2024-12-11 | | 212143.89) |
| Washington | Snohomish | Snohomish, WA | City of Snohomish Wastewater Treatment Plant | 10150 | 2023-10-18, 2024-12-11 | 178 | 9404.2 (0, 51977.35) |
| West Virginia | Ohio | Wheeling, WV | City of Wheeling, Water Pollution Control Division | 100000 | 2023-09-08, 2024-12-11 | 190 | 12339.89 (2691.68, 47594.17) |
| Wisconsin | Marathon | Wausau, WI | Wausau Waterworks Wastewater Treatment Facility | 44000 | 2023-09-09, 2024-12-09 | 196 | 85824.87 (5817.24, 500574.17) |

[1] For WWTPs servicing multiple counties, the county predominantly serviced by the WWTP is listed. We determined the predominant county intersecting sewersheds using the Tabulate Intersection geoprocessing tool in ArcGIS Pro (version 3.1.1) and 2022 county boundaries at the 5-m resolution from the US Census Bureau (accessed 22 Feb 2024).[3]

Abbreviations: WWTP = wastewater treatment plant, gc/g = gene copies per gram, IQR = interquartile range

### Rotavirus assay: Further details

**Text S1. *In silico* evaluation**

The rotavirus genome consists of 11 linear double-stranded RNA segments.[4] The primers and probes used in our study to detect rotavirus RNA in wastewater are based on a previously published assay that targets the gene encoding the nonstructural protein 3 (NSP3),[5] which is located on segment 7.[6] We evaluated whether the assay may amplify human rotavirus vaccine strains, animal rotavirus strains, or the bovine rota-coronavirus vaccine used as an exogenous control.

***Amplification of vaccine strains.*** There are two human rotavirus vaccines licensed in the United States. RotaTeq is a pentavalent reassortant vaccine based on a live-attenuated human-bovine rotavirus strain (4 reassortants with the VP7 gene from the bovine strain and 1 reassortant with the VP4 P1A[8] gene from the human strain).[4,7] Rotarix is a monovalent vaccine based on a live-attenuated human rotavirus strain G1P[8].[4,7] The NSP3 gene sequence is available for five RotaTeq vaccine strains (GenBank accession numbers GU565049.1, GU565060.1, GU565071.1, GU565082.1, GU565093.1) and one Rotarix vaccine strain (GenBank accession number KX954622.1). We checked for binding (defined as matching bases with no more than 3 total mismatches) with the assay primers and probe. For all the RotaTeq NSP3 gene sequences, the forward primer binds with no mismatches and the probe and reverse primer bind with two mismatches each. For the Rotarix NSP3 gene sequence, the forward primer does not bind and the probe and reverse primer bind with no mismatches each. Overall, it is improbable the rotavirus assay used herein amplifies either rotavirus vaccine strain.

***Amplification of animal strains.*** We obtained sequences of segment 7 RNA from several animal rotaviruses (avian, bovine, chiropteran, equine, porcine). We checked for binding (defined as matching bases with no more than 3 total mismatches) with the assay primers and probe. Overall, the primers and probe entirely or partially (e.g., just the forward primer) did not bind to the segment 7 sequences. When both primers and the probe binded to the segment 7 sequence (most common for equine and porcine strains), there was often at least 1 mismatch still present with one of the primers or the probe. Amplification of some animal strains may be possible but is likely negligible.

***Amplification of exogenous control.*** A bovine rota-coronavirus vaccine (https://www.pbsanimalhealth.com/calf-guard-cattle-vaccine/p/11712/?v=2), which contains modified live virus, is spiked into wastewater samples as an exogenous control.[8] No amplification has been observed in the ddRT-PCR process control, confirming the rotavirus assay used herein does not amplify the exogenous control.

#### Population characteristics: Further details

**Data source: American Community Survey (ACS)**

We used estimates from the 2022 5-year ACS to determine characteristics of sewersheds outlined in **Table S4**.[9] We obtained ACS variables at the census tract resolution; however, census tract and sewershed boundaries do not align, so we aggregated data across census tracts to obtain sewershed-level estimates. Specifically, we used the Tabulate Intersection geoprocessing tool in ArcGIS Pro (version 3.1.1) to determine the area proportion $p$ of each census tract $n$ intersecting each sewershed; we obtained 2022 census tract boundaries at the 5-m resolution from the US Census Bureau (accessed 15 Feb 2024).[3] For count ACS variables (i.e., all variables in **Table S4** except S1101_C01_004E), we then adjusted the census tract-level count based on $p$ (**Equation S1**). Next, we summed adjusted counts across all $N$ census tracts intersecting a sewershed to determine the sewershed-level ACS variable count (**Equation S2**). We then calculated the sewershed-level characteristics described in **Table S4** as proportions using sewershed-level counts of each ACS variable. For S1101_C01_004E (average family size) which is already reported as a proportion at the census tract level, we approximated the sewershed-level average family size as the median average family size across all census tracts intersecting a sewershed. The distribution of each population characteristic among sewersheds is shown in **Figure S7**.

**Equation S1.** $count\_adjusted_n = count_n \times p_n$

**Equation S2.** $count\_sewershed = \sum_{n=1}^{N} count\_adjusted_n$

**Table S3. Sewershed characteristics derived from the American Community Survey**

| Characteristic | Calculation Using ACS variables[1] |
|---|---|
| Percent of children under 5 years, Black or African American alone | (B01001B_003E + B01001B_018E) / S0101_C01_002E |
| Percent of children under 5 years, Hispanic or Latino | (B01001I_003E + B01001I_018E) / S0101_C01_002E |
| Percent of children under 5 years, foreign born | B06001_050E / S0101_C01_002E |
| Percent of children under 5 years, income in the past 12 months below the federal poverty level | (B17001_004E + B17001_018E) / S0101_C01_002E |
| Percent of children under 6 years, no health insurance | (B27001_005E + B27001_033E) / (B27001_003E + B27001_031E) |
| Percent of children under 6 years, public health insurance | (B27003_004E + B27003_032E) / (B27001_003E + B27001_031E) |
| Percent of women who gave birth in the past 12 months, ages 15-24 years | (B13016_003E + B13016_004E) / B13016_002E |
| Average family size | S1101_C01_004E |

[1] To calculate sewershed population proportions, population counts of census tracts were first adjusted based on the proportion of the tract intersecting the sewershed and then summed by sewershed to obtain sewershed-level population counts. The sewershed average family size is the median average family size across all census tracts intersecting the sewershed.

Abbreviation: ACS = American Community Survey

#### Data source: American Board of Pediatrics (ABP)

We used 2024 counts of pediatricians by county from the ABP to estimate the number of pediatricians per 100 thousand children < 18 years in each sewershed.[10] General pediatricians aged 70 and under who are currently certified by the ABP are categorized as being certified in general pediatrics alone or in general pediatrics alongside another specialty. We used the sum of both categorizations to determine all currently certified general pediatricians, regardless of certification in another speciality, by county. The ABP dataset also includes the count of children < 18 years from the 2023 ACS 5-year estimates in each county. Using these county-level counts, we calculated the number of pediatricians per 100 thousand children < 18 years in each county as follows: $all\ pediatricians\ /\ children\ under\ 18\ *\ 100,000$. We approximated the pediatrician:child ratio of a sewershed as the pediatrician:child ratio of the predominant county intersecting the sewershed (see **Table S2**). The distribution of pediatrician:child ratios among sewersheds is shown in **Figure S8**.

**Data source: US Census Bureau**

We used 2020 urban-rural designations from the US Census Bureau to determine the proportion of sewersheds classified as urban.[11] We used the Tabulate Intersection geoprocessing tool in ArcGIS Pro (version 3.1.1) to determine the the proportion of each sewershed area intersecting an urban-designated area. The distribution of urbanicity among sewersheds is shown in **Figure S9**.

We used census regions from the US Census Bureau to determine the census region of sewersheds.[12] We assigned sewersheds to a census region based on the state that the WWTP servicing the sewershed is located in. Refer to **Figure 6** for the classification of states by census region. The number of WWTPs in each census region is shown in **Figure S10**.

#### Spatial aggregation of wastewater monitoring data

We used a population-weighted averaging approach to spatially aggregate wastewater measurements across WWTP sewersheds to the state (**Equation S3**) and national (**Equation S4**) spatial scales.

**Equation S3.** $avg\_rota\_state = \frac{\sum_{n=1}^{N} pop_n \times rota_n}{\sum_{n=1}^{N} pop_n}$

$avg\_rota\_state$ represents the population-weighted average wastewater rotavirus RNA measurement for a given state from $N$ WWTPs in the state, where $pop_n$ is the population serviced by WWTP $n$ and $rota_n$ is the wastewater rotavirus RNA measurement from WWTP $n$. WWTP service populations ($pop_n$) are listed in **Table S2**.

**Equation S4.** $avg\_rota\_USA = \frac{\sum_{s=1}^{S} pop_s \times avg\_rota\_state_s}{\sum_{s=1}^{S} pop_s}$

$avg\_rota\_USA$ represents the population-weighted average wastewater rotavirus RNA measurement from $S$ states in the United States, where $pop_s$ is the population of state $s$ and $avg\_rota\_state_s$ is the population-weighted average wastewater rotavirus RNA measurement of state $s$ (see **Equation S3**). We obtained 2023 state populations ($pop_s$) from the US Census.[13]

#### Sewershed vaccination coverage: Further details

As described in the main text, rotavirus vaccination coverage is not directly estimated at the sewershed scale, so we used the vaccination coverage estimate of the county that a sewershed predominantly occupies (see **Table S2**) to represent the vaccination coverage of sewersheds. County-level rotavirus vaccination coverage estimates are only available from Epic Cosmos. We used the Epic Cosmos county "District of Columbia, DC" to represent the predominant county of the Blue Plains, Washington, DC sewershed and the Epic Cosmos county "Fairfield, CT" to represent the predominant county of the Stamford, CT sewershed; the Epic Cosmos county used for all other sewersheds remained the same as **Table S2**. **Figure S15** shows the distribution of rotavirus vaccination coverage among sewersheds.

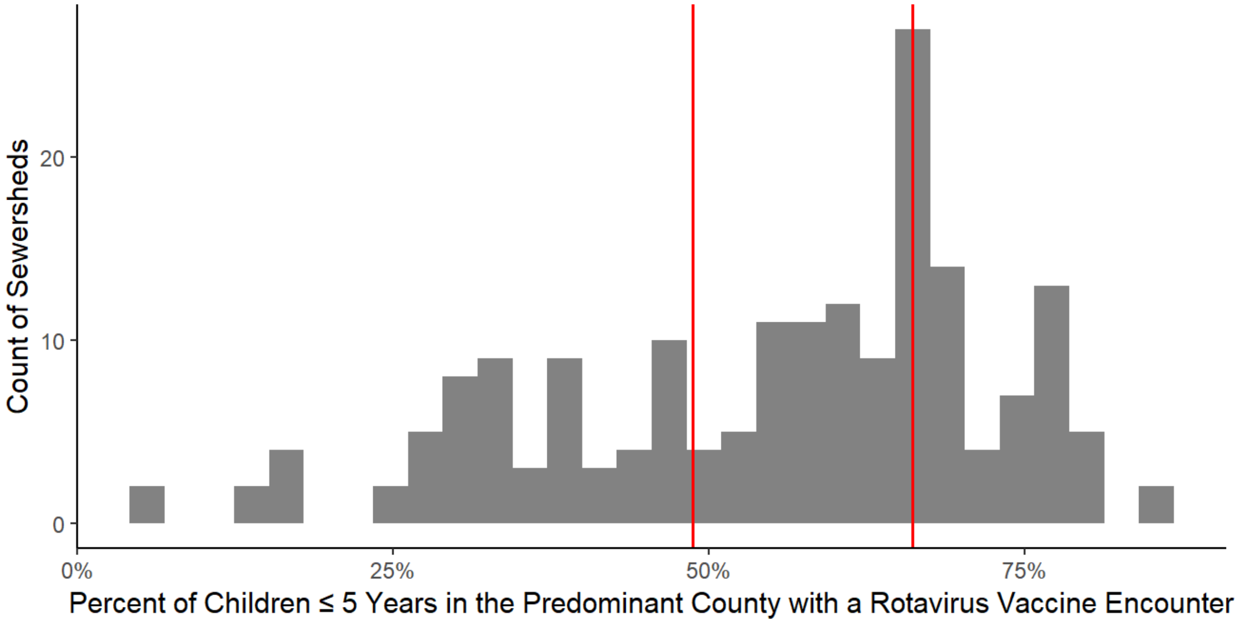


**Figure S15. Distribution of rotavirus vaccination coverage estimates from Epic Cosmos among sewersheds.** Vertical, red lines indicate tertile cutoffs.

### Mass balance model: Further details

Wolfe et al.[14] established a mass balance model relating the fraction of the population shedding SARS-CoV-2 RNA into wastewater from SARS-CoV-2 RNA measurements in wastewater solids. The model is not virus specific. Boehm et al.[15] adopted the model for norovirus GII RNA; herein we adopted the model for rotavirus RNA (**Equation S5**).

**Equation S5**. $$F_{shed} = \frac{C_{rota\_ww} C_{PMMoV\_feces} K_{dp} (1 + K_d TSS) exp(t(k_r - k_p))}{C_{PMMoV\_ww} K_d (1 + K_{dp} TSS) C_{rota\_feces}}$$

$F_{shed}$ = population fraction shedding rotavirus RNA in feces
$C_{rota\_ww}$ = concentration of rotavirus RNA in wastewater solids
$C_{PMMoV\_ww}$ = concentration of PMMoV RNA in wastewater solids
$C_{rota\_feces}$ = concentration of rotavirus RNA shed in feces
$C_{PMMoV\_feces}$ = concentration of PMMoV RNA shed in feces
$K_d$ = rotavirus RNA solid-liquid partitioning coefficient
$K_{dp}$ = PMMoV RNA solid-liquid partitioning coefficient
TSS = total suspended solids in the influent
t = time sewage spends in the system, including primary clarifier, prior to sampling
$k_r$ = rotavirus RNA first order decay rate constant
$k_p$ = PMMoV RNA first order decay rate constant

Similar to Boehm et al.,[15] we neglect the decay term given t is short (< 1 day) and $k_r$ and $k_p$ can reasonably be assumed to be both small and similar in magnitude (**Equation S6**).[14,16–19]

**Equation S6.** $$F_{shed} = \frac{C_{rota\_ww} C_{PMMoV\_feces} K_{dp} (1 + K_d TSS)}{C_{PMMoV\_ww} K_d (1 + K_{dp} TSS) C_{rota\_feces}}$$

Lastly, we assume $K_d = K_{dp}$ like Boehm et al.[15], simplifying the model to **Equation 1** in the main text.
